# Epidemiologic and economic modelling of optimal COVID-19 policy: public health and social measures, masks and vaccines in Victoria, Australia

**DOI:** 10.1101/2022.08.01.22278262

**Authors:** Joshua Szanyi, Tim Wilson, Samantha Howe, Jessie Zeng, Hassan Andrabi, Shania Rossiter, Tony Blakely

**Affiliations:** Population Interventions Unit, Centre for Epidemiology and Biostatistics, Melbourne School of Population and Global Health, The University of Melbourne, Melbourne, Australia

## Abstract

**Background:** Identifying optimal COVID-19 policies is challenging. For Victoria, Australia (6.6 million people), we evaluated 104 policy packages (two levels of stringency of public health and social measures [PHSMs], by two levels each of mask-wearing and respirator provision during large outbreaks, by 13 vaccination schedules) for nine future SARS-CoV-2 variant scenarios.

**Methods:** We used an agent-based model to estimate morbidity, mortality, and costs over 12 months from October 2022 for each scenario. The 104 policies (each averaged over the nine future variant scenarios) were ranked based on four evenly weighted criteria: cost-effectiveness from (a) health system only and (b) health system plus GDP perspectives, (c) deaths and (d) days exceeding hospital occupancy thresholds.

**Findings:** More compared to less stringent PHSMs reduced cumulative infections, hospitalisations and deaths but also increased time in stage ≥3 PHSMs. Any further vaccination from October 2022 decreased hospitalisations and deaths by 12% and 27% respectively compared to no further vaccination and was usually a cost-saving intervention from a health expenditure plus GDP perspective. High versus low vaccine coverage decreased deaths by 15% and reduced time in stage ≥3 PHSMs by 20%. The modelled mask policies had modest impacts on morbidity, mortality, and health system pressure. The highest-ranking policy combination was more stringent PHSMs, two further vaccine doses (an Omicron-targeted vaccine followed by a multivalent vaccine) for ≥30-year-olds with high uptake, and promotion of increased mask wearing (but not Government provision of respirators).

**Interpretation:** Ongoing vaccination and PHSMs continue to be key components of the COVID-19 pandemic response. Integrated epidemiologic and economic modelling, as exemplified in this paper, can be rapidly updated and used in pandemic decision making.

**Funding:** Anonymous donation, University of Melbourne funding.

**Abstract:** 

**Background:** Identifying optimal COVID-19 policies is challenging. For Victoria, Australia (6.6 million people), we evaluated 104 policy packages: (a) two levels of stringency of public health and social measures (PHSMs; lower, higher), by (b) two levels each of mask wearing (low, high) and Government respirator provision (nil, yes) during large outbreaks (defined as when the projected number of people in hospital reached >270 or >130 per million population for lower and higher stringency PHSM settings respectively), by (c) 13 vaccination schedules (nil, and four combinations of low/high coverage for ≥30/60-year-olds, each with an Omicron-targeted (OT) booster in the last quarter of 2022 followed by one of: nil, another OT booster in the second quarter of 2023, or a multivalent booster in the second quarter of 2023). These policies were modelled in the setting of nine future SARS-CoV-2 variant scenarios (no major new variant of concern and one of eight variants arriving in November 2022 with different virulence, antigenic, and immune escape profiles).

**Methods:** We used an agent-based model to estimate morbidity, mortality, and costs over 12 months from October 2022 for each scenario. The 104 policies (each averaged over the nine future variant scenarios) were ranked based on four evenly weighted criteria: cost-effectiveness from (a) health system only and (b) health system plus GDP perspectives (HALYs valued at AUD 70,000; discount rate 3%), (c) deaths and (d) days exceeding hospital occupancy thresholds.

**Findings:** More compared to less stringent PHSMs reduced cumulative infections, hospitalisations and deaths by an average of 25%, 24% and 24% respectively across 468 policy comparisons (other policy and variant scenarios held constant), but also increased time in stage ≥3 (out of 5) PHSMs by an average of 42 days (23 days for low virulence and 70 days for high virulence variants).

Any further vaccination from October 2022 decreased hospitalisations and deaths by 12% and 27% respectively compared to no further vaccination, however the cumulative number of infections increased by 10% due to vaccination preferentially decreasing hospitalisation rates that were used to dynamically set PHSM stages. Any further vaccination was of marginal cost-effectiveness from a health system perspective (an average of AUD 77,500 per HALY gained for vaccinating ≥60-year-olds, and AUD 41,600 for 30- to 59-year-olds incremental to ≥60-year-olds), but vaccination also resulted in 36% fewer days in Stage ≥3 PHSMs usually making it a cost-saving intervention from a health expenditure plus GDP perspective. High versus low vaccine coverage reduced deaths by 15% and reduced time in Stage ≥3 PHSMs by 20%.

Promotion to increase mask wearing or government provision of respirators during large outbreaks reduced cumulative infections, hospitalisations and deaths over the 12 months by 1% to 2%, and reduced days with hospital occupancy exceeding 750 COVID-19 patients by 2% (4% to 5% in the context of highly virulent variants).

The highest-ranking policy combination was more stringent PHSMs, two further vaccine doses (an Omicron-targeted vaccine followed by a multivalent vaccine) for ≥30-year-olds with high uptake, and promotion of increased mask wearing (but not Government provision of respirators).

**Funding:** Anonymous donation, University of Melbourne funding.

**Research in context:** 

**Evidence before this study:** We searched Ovid MEDLINE to 28 July 2022 for studies using the terms (economic evaluation.mp. OR cost effectiveness.mp. OR health economic*.mp.) AND (simulation.mp. OR model*.mp.) AND pandemic*.mp. to identify existing simulation modelling analyses of pandemic preparedness and response that incorporated cost effectiveness considerations. All identified literature examined pandemic influenza and COVID-19 and was highly heterogeneous in terms of modelled interventions (which included school closures, masks, hand hygiene, vaccination, testing strategies, antiviral medication, physical distancing measures, indoor ventilation, and personal protective equipment), quality, context, model structure, and economic evaluation approach.

Systematic reviews of COVID-19 modelling studies that include a health economic component generally indicate that SARS-CoV-2 testing, personal protective equipment, masks, and physical distancing measures are cost-effective. However, few prior studies consider optimal packages of interventions (as opposed to standalone interventions), and none explicitly account for ongoing viral evolution or accurately capture the complexities of vaccine- or natural infection-derived immunity to SARS-CoV-2.

For example, a previous study integrating a dynamic SARS-CoV-2 transmission model with an economic analysis using a net monetary benefit approach published in early 2021 emphasized the combined public health and economic advantages of COVID-19 vaccination combined with physical distancing measures in the UK. However, considering current knowledge regarding the substantial waning of vaccine effectiveness and relatively low protection against infection conferred by vaccination (compared to more severe clinical outcomes), this model likely over-estimated the impact of COVID-19 vaccination on viral transmission. Scenarios that considered the emergence of SARS-CoV-2 variants of concern and thus associated changes in viral transmissibility, immune escape capacity (which has, in the case of the Omicron variant, greatly reduced protection following vaccination and prior infection) or virulence were also not modelled.

**Added value of this study:** To our knowledge, our study is the first that utilises a dynamic disease transmission model combined with an integrated economic evaluation framework to systematically compare COVID-19 policy intervention packages while accounting for ongoing SARS-CoV-2 evolution and waning population immunity. At a high-level, we found that a considerable degree of COVID-19 disease burden should be expected in the future, with modelled interventions only able to partly mitigate pandemic-associated morbidity and mortality in the medium-term.

Across nine plausible future SARS-CoV-2 variant scenarios, higher stringency PHSMs notably reduced cumulative infections, hospitalisations and deaths in the 12-month period modelled but had the tradeoff of higher expected societal economic losses. Increasing community mask-wearing and substituting cloth and surgical masks for government supplied respirators during periods of high SARS-CoV-2 morbidity both reduced the number of days with hospital occupancy exceeding 750 COVID-19 patients by 2% on average across scenarios, and minimally reduced the cumulative infection, hospitalization and death burden. Compared to no further vaccines, the modelled vaccination schedules (with next-generation vaccines; one or two further doses) reduced hospitalisations by an average of 12%, and deaths by 27%. Vaccinating ≥30-year-olds was modestly superior to just vaccinating ≥60-year-olds (reducing cumulative deaths, for example, by 3.1%).

Considering all policy options together, and ranking by optimality on cost-effectiveness, health system pressure and deaths, the highest ranking policy combinations tended to be a mix of higher stringency PHSMs, promotion to increase mask wearing but no Government-funded respirator provision during large outbreaks, and the administration of two booster vaccine doses within the 12-month period to ≥30-year-olds with associated high coverage (noting gains from vaccinating ≥30-year-olds compared to ≥60-year-olds were modest).

**Implications of all the available evidence:** The policy implications of this study are three-fold. Firstly, it reinforces the cost-effectiveness of ongoing vaccination of the public to mitigate morbidity and mortality associated with COVID-19. Secondly, the characteristics of emerging SARS-CoV-2 variants, outside the control of policy makers, will likely substantially influence public health outcomes associated with the pandemic in the future. Finally, at a phase of the pandemic characterised by growing intervention options urgently requiring prioritisation by decision makers alongside a large degree of ongoing uncertainty about future variants, this study provides a framework within which to systematically compare the health and economic benefits and burdens of packages of interventions that can be rapidly updated with new information (such as estimated effectiveness and waning kinetics of newly-developed vaccines) to support policy making.

## Introduction

The COVID-19 pandemic is well into its third year, with ongoing high levels of SARS-CoV-2 transmission driving significant morbidity and mortality globally. This is due in part to the emergence of variants, such as Omicron and its sub-variants, that possess enhanced capacity to evade pre-existing immunity. Continued SARS-CoV-2 evolution is likely,^1^ now occurring against a backdrop of dynamic population immunity (from vaccination, natural infection, or both) and an expanding array of public health and clinical intervention options to respond to the pandemic. As such, COVID-19-related policy decisions must be made in the context of substantial uncertainty, a significant challenge for policy makers. In this complex environment it is increasingly important that the benefits and drawbacks of interventions are rigorously and systematically compared – including from a cost effectiveness perspective.^2^

In response to these needs we developed an integrated epidemiologic and economic simulation model to determine the optimal of 104 illustrative policy packages (two stringency levels of public health and social measures [PHSMs], two respirator provision policies, two levels of baseline mask compliance during large outbreaks, and 13 vaccination schedules) for the state of Victoria, Australia. Each policy was modelled in the context of nine future SARS-CoV-2 variant scenarios (eight combinations of low or high virulence, low or high antigenic similarity to the Omicron variant, and low or high immune escape capacity, in addition to a scenario of no new variant), emerging in November 2022 following successive periods of Omicron BA.1/2 and Omicron BA.4/5 dominance from April 2022. Policies were then ranked based on cumulative deaths, hospital system pressure, and cost effectiveness from both health system and health system plus GDP perspectives in the 12 months from October 2022, providing a framework for assessing optimal pandemic policy in the face of a rapidly evolving and uncertain future.

## Methods

### Agent-based model

We used an agent-based model (ABM)^3,4^ with a daily cycle length and 5,000 agents scaled up to represent the Victorian population. Each agent moves in a two-dimensional space, creating opportunities for infection informed by parameters that influence viral transmission (Table 1, Appendix). The model was initially calibrated to the first COVID-19 waves in Australia and New Zealand and has previously been used to inform policy in Victoria. For this study, the model was initiated with a virus reflecting Omicron BA.1/2 on 1 April 2022, then a variant with immune escape representing Omicron BA.4/5 was introduced to the model on 1 May 2022. The model was then calibrated to match SARS-CoV-2 transmission data from Victoria over the 60 days from 1 April 2022 and validated against case report (assuming 50% case ascertainment) and hospitalisation data from Victoria between April and September 2022 (Appendix). The former involved the addition of a “carefulness” parameter that interacted with biological susceptibility to infection to reflect varying infection-avoidance behaviour by age and achieve the age distribution of infections occurring in Victoria during the calibration period. For the 12 months from 1 October 2022, 936 scenarios (104 policy packages combined with nine viral variant scenarios) were run 500 times each (500 separate draws of input parameters) to generate estimates of COVID-19-related morbidity, mortality and costs over this period.

**Table 1:**
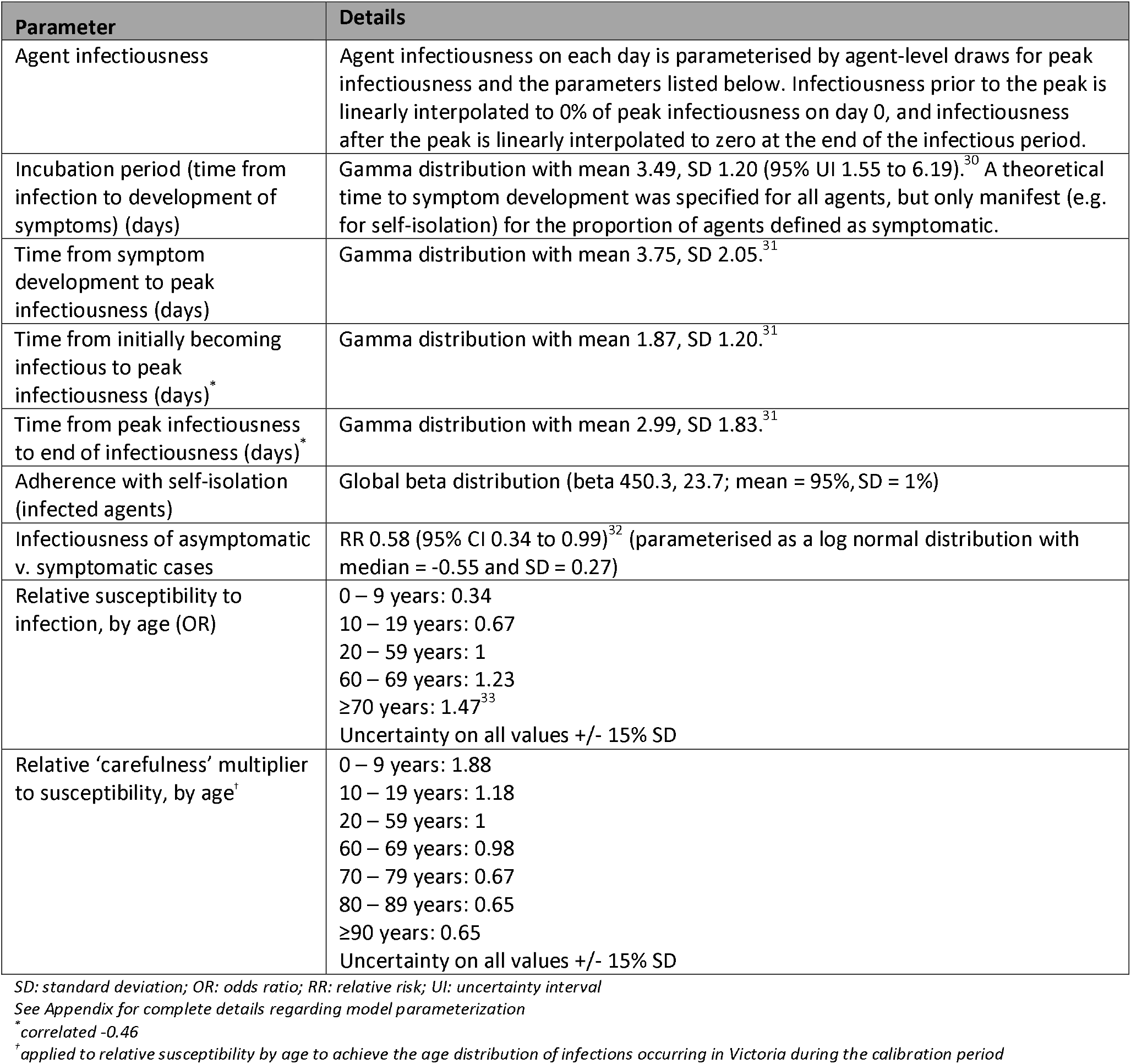
Key model input parameters.

| Parameter | Details |
| --- | --- |
| Agent infectiousness | Agent infectiousness on each day is parameterised by agent-level draws for peak infectiousness and the parameters listed below. Infectiousness prior to the peak is linearly interpolated to 0% of peak infectiousness on day 0, and infectiousness after the peak is linearly interpolated to zero at the end of the infectious period. |
| Incubation period (time from infection to development of symptoms) (days) | Gamma distribution with mean 3.49, SD 1.20 (95% UI 1.55 to 6.19).<sup>30</sup> A theoretical time to symptom development was specified for all agents, but only manifest (e.g. for self-isolation) for the proportion of agents defined as symptomatic. |
| Time from symptom development to peak infectiousness (days) | Gamma distribution with mean 3.75, SD 2.05.<sup>31</sup> |
| Time from initially becoming infectious to peak infectiousness (days)* | Gamma distribution with mean 1.87, SD 1.20.<sup>31</sup> |
| Time from peak infectiousness to end of infectiousness (days)* | Gamma distribution with mean 2.99, SD 1.83.<sup>31</sup> |
| Adherence with self-isolation (infected agents) | Global beta distribution (beta 450.3, 23.7; mean = 95%, SD = 1%) |
| Infectiousness of asymptomatic v. symptomatic cases | RR 0.58 (95% CI 0.34 to 0.99)<sup>32</sup> (parameterised as a log normal distribution with median = -0.55 and SD = 0.27) |
| Relative susceptibility to infection, by age (OR) | 0 – 9 years: 0.34<br>10 – 19 years: 0.67<br>20 – 59 years: 1<br>60 – 69 years: 1.23<br>≥70 years: 1.47<sup>33</sup><br>Uncertainty on all values +/- 15% SD |
| Relative 'carefulness' multiplier to susceptibility, by age<sup>†</sup> | 0 – 9 years: 1.88<br>10 – 19 years: 1.18<br>20 – 59 years: 1<br>60 – 69 years: 0.98<br>70 – 79 years: 0.67<br>80 – 89 years: 0.65<br>≥90 years: 0.65<br>Uncertainty on all values +/- 15% SD |

SD: standard deviation; OR: odds ratio; RR: relative risk; UI: uncertainty interval
See Appendix for complete details regarding model parameterization
\*correlated -0.46
<sup>†</sup>applied to relative susceptibility by age to achieve the age distribution of infections occurring in Victoria during the calibration period

### Modelled scenarios

#### Policy options

Five stages of PHSMs were specified (Appendix Table 4). Stages incrementally impose more restrictions up to stage 5, which approximates a lockdown. The ABM (de)escalates through these stages based on hospital occupancy thresholds and two PHSM policy options (higher and lower stringency; Table 2).

**Table 2:** Modelled policy options.

| Parameter | Details |  |  |  |  |  |
| --- | --- | --- | --- | --- | --- | --- |
| *PHSM strategy* |  |  |  |  |  |  |
| Lower stringency | Escalation: if the average expected number of people in hospital due to COVID-19 10-14 days into the future is estimated to be >600 per million → Stage 5; >400 per million → Stage 4; >270 per million → Stage 3; >180 per million → Stage 2.<br>De-escalation: if no de-escalation in last 7 days, and average expected number of people in hospital 10-14 days into the future is estimated to be <450 → Stage 4 if in Stage 5; <300 → Stage 3 if in stage 4 or 5; <200 → Stage 2 if in Stage 3, 4 or 5; <140 → Stage 1. |  |  |  |  |  |
| Higher stringency | Thresholds approximately half those for lower stringency |  |  |  |  |  |
| *Mask compliance strategy* |  |  |  |  |  |  |
| Baseline | Baseline mask wearing at locations other than the home: |  |  |  |  |  |
|  | *Age* | *Stage 1* | *Stage 2* | *Stage 3* | *Stage 4* | *Stage 5* |
|  | ≥20-year-olds | 6% | 16.3% | 45% | 66% | 80% |
|  | 10- to 19-year-olds | 4% | 10.9% | 30% | 44% | 53.3% |
|  | <10-year-olds | 2.7% | 7.2% | 20% | 29.3% | 35.6% |
|  | Of mask wearers, 20% assumed to be using respirators, 80% cloth or surgical masks (unless respirator supply policy active). Odds ratios of 2, 4, 6 and 8 are applied to the proportion of people that wear a mask for those aged 50-59, 60-69, 70-79 and ≥80 years respectively, compared to 20- to 49-year-olds. |  |  |  |  |  |
| Higher compliance | In stages 3 to 5 the proportions of mask wearing above are increased by an odds ratio of 2. Respirator use remains 20% of the resultant overall mask use unless the respirator supply policy (below) is active. |  |  |  |  |  |
| *Respirator strategy* |  |  |  |  |  |  |
| Baseline | As outlined above. |  |  |  |  |  |
| Government supply of respirators | Same population use of masks as above, but in stages 3 to 5 the proportion of mask use that is with respirators increases from 20% to 80% (for those aged ≥10 years only). 10 respirators are stockpiled and provided per person aged ≥10 years every four weeks spent in stages 3 and above. |  |  |  |  |  |
| *Vaccination strategy* |  |  |  |  |  |  |
| Vaccine schedules | Three generic vaccines were modelled: first-generation mRNA vaccines (FG); next-generation Omicron-targeted vaccines (OT) with vaccine effectiveness (VE) on an odds scale of 2 against variants that are antigenically like Omicron compared to FG; and next-generation multivalent vaccines (MV) with VE on an odds scale of 2 against all variants compared to FG.<br>Low uptake: 50% for ≥60-year-olds, 25% for 30- to 60-year-olds<br>High uptake: 75% for ≥60-year-olds, 50% for 30- to 60-year-olds<br>13 vaccine schedules were specified (following FG vaccine administration until end of Q2 2022):<br>1. Nil further vaccination beyond Q2 2022<br>2. OT vaccine in Q4 2022, ≥60-year-olds, low uptake<br>3. OT vaccine in Q4 2022, ≥60-year-olds, high uptake<br>4. OT vaccine in Q4 2022, ≥30-year-olds, low uptake<br>5. OT vaccine in Q4 2022, ≥30-year-olds, high uptake<br>6. OT vaccine in Q4 2022 & OT vaccine in Q2 2023, ≥60-year-olds, low uptake<br>7. OT vaccine in Q4 2022 & OT vaccine in Q2 2023, ≥60-year-olds, high uptake<br>8. OT vaccine in Q4 2022 & OT vaccine in Q2 2023, ≥30-year-olds, low uptake |  |  |  |  |  |

| Parameter | Details |
| --- | --- |
|  | 9. OT vaccine in Q4 2022 & OT vaccine in Q2 2023, ≥30-year-olds, high uptake<br>10. OT vaccine in Q4 2022 & MV vaccine in Q2 2023, ≥60-year-olds, low uptake<br>11. OT vaccine in Q4 2022 & MV vaccine in Q2 2023, ≥60-year-olds, high uptake<br>12. OT vaccine in Q4 2022 & MV vaccine in Q2 2023, ≥30-year-olds, low uptake<br>13. OT vaccine in Q4 2022 & MV vaccine in Q2 2023, ≥30-year-olds, high uptake |

We modelled two mask policies – (a) a respirator (e.g., N95 mask) substitution policy with options of no respirator provision or a respirator stockpile that is distributed to the population for use in place of cloth or surgical masks during large outbreaks (defined as when the model was in stage ≥3 of PHSMs) and (b) promotion of mask wearing during large outbreaks. The respirator policy did not change the overall percentage of people wearing masks but shifted respirator (compared to cloth or surgical) use from 20% to 80% among mask users. The mask promotion policy doubled overall mask use on the odds scale, e.g., 40% use becomes 57% use (odds of use doubles from 40%/60% = 0.67 to 57%/43% = 1.33). These policies were only active after the six-month lead-in period (i.e., the final quarter of 2022 onwards). Cloth and surgical masks were parameterised as reducing the odds of infection by approximately 50% compared to no mask, and respirators by approximately 80% (Appendix).^5^

Thirteen future vaccine schedules were modelled, incorporating next-generation COVID-19 vaccines specifically targeting the Omicron variant or targeting several variants (multivalent vaccines). The 13 vaccine schedules were nil further vaccination from October 2022, or 12 combinations of high or low uptake (high = 75% and 50%, low = 50% and 25% for ≥60-year-olds and 30-to 60-year-olds respectively), for ≥30-or ≥60-year-olds, with an Omicron-targeted (OT) vaccine in October to December 2022 plus one of no further doses, another OT vaccine in April to June 2023, or a multivalent vaccine in April to June 2023. The multivalent vaccine was specified to have twice the vaccine effectiveness (VE) on the odds scale of current mRNA vaccines against all variants, and the Omicron-targeted vaccine twice the VE on the odds scale for variants antigenically like Omicron (Appendix). A first-generation mRNA vaccine (i.e., a vaccine developed from the ancestral variant of SARS-CoV-2) was administered by default in all scenarios to reach the number of people vaccinated with three or four doses in Victoria as of the end of June 2022. Subsequent vaccine schedules were then applied to individuals who had received at least three vaccine doses by the time each dose was rolled out. All modelled policy scenarios are summarized in Table 2.

#### Variant scenarios

The dominant variant on 1 April 2022 was parameterised to approximate the BA.1/2 Omicron sub-variants (R_0_ 8-10, lower virulence), and gradually (over approximately two months from 1 May 2022) replaced by a variant with additional immune escape capacity (approximating Omicron BA.4/5). The emergence of eight potential new variants (plus a scenario of no new variant emergence, i.e., continued Omicron BA.4/5 dominance) from November 2022 was then modelled.

New variants were characterized as either low virulence (approximating Omicron) or high virulence. To set the low virulence infection fatality risk (IFR), we scaled age-specific IFRs associated with the ancestral variant^6^ to match deaths observed in Victoria in April and May 2022 (an Omicron BA.2-dominant period), taking into account previous infection and vaccination. This process was repeated using hospital and ICU admission risks.^7^ We parameterised hypothetical high virulence future variants by assuming 4, 4^0.75^, 4^0.5^ and 4^0.25^ ratio differences (on an odds scale) in IFR, ICU admission risk, hospital admission risk and probability of being symptomatic given infection, respectively, between low and high virulence variants. This parameterisation aims to capture a shift in severity across the spectrum of clinical disease, meaning that a high compared to low virulence variant increases symptomatic infections by a ratio of 1.41, hospitalisation if already symptomatic by 1.41, ICU admission if hospitalised by 1.41, and death as a ratio to ICU admissions by 1.41. These increases loosely approximate Delta versus Omicron differences within strata of previous vaccination and infection status.^8^

The immune escape capacity of new variants over that already possessed by Omicron BA.4/5 was set as low or high using odds ratios (ORs) applied to VE estimates (Appendix). Antigenic similarity to Omicron influenced the effectiveness of Omicron-targeted vaccines against new variants with immune escape.

### Vaccine effectiveness and protection against reinfection

Protection following vaccination or previous infection was a function of age, time since last vaccine or infection (i.e., waning), the number and type of vaccine doses received, and the variant responsible for primary infection, based on a previously published model of VE (Appendix).^9^

### Morbidity and mortality

We quantified acute COVID-19 morbidity using disability rates (DRs) from the Global Burden of Disease (GBD) study.^10^ Morbidity was calculated separately for high and low virulence variant infections by altering the duration of illness and length of hospital stay.^11,12^ Morbidity from long COVID was estimated based on reported symptoms and their prevalence and duration (by age, severity of infection, vaccination status, and viral variant), each assigned a disability weight from the GBD study (Appendix).^10,13-16^ For each COVID-19 death we estimated future HALY loss (discounted at 3%), assuming people dying of COVID-19 have twice the mortality and 1.5 times the morbidity of the average person of the same sex and age (Appendix).

### Economic analyses

We used a net monetary benefit (NMB) approach where total net health expenditure was subtracted from monetized HALYs at a given willingness to pay (WTP; Australian GDP per capita [AUD 70,000] per HALY unless otherwise specified) in each model iteration, with two perspectives considered – health system only, and health system plus GDP loss (Appendix). For acute illness we applied unit costs to each agent depending on their infection and clinical outcome status, including for testing, medication, ambulatory care, and hospital costs. We also determined healthcare utilisation costs for those experiencing long COVID based on international data and Australian clinical guidelines, stratified by acute disease severity, variant virulence, and vaccination status (Appendix).

Interventions were costed using the unit costs of vaccines and respirators, in addition to transportation, storage, vaccine administration, respirator distribution and health promotion costs where applicable (Appendix). Net health expenditure was the sum of intervention costs (e.g., warehousing masks, purchasing vaccines), the immediate costs of treating acute and long-COVID, plus the difference between scenarios in future health expenditure. Costs to society due to PHSMs were assumed to be nil for stages 1 and 2, and 10% to 50% of the Australian Government-estimated GDP losses per week from 2020^17^ for approximately equivalent levels of restrictions in stages 3 to 5. Health expenditure and GDP losses were all discounted at 3% per annum. All costs are reported in 2021 Australian dollars (1 AUD = 0.695 USD in 2021 using OECD purchasing power parities).

### Ranking

We ranked each policy option, averaged over all variant scenarios, by: (a) the cumulative number of SARS-CoV-2 deaths over 12 months, (b) the number of days hospital occupancy by COVID-19 patients was >750 (114 per million) or >1500 (227 per million), (c) NMB from a health system perspective, and (d) NMB from a health system plus GDP perspective. We then generated an average ranking across these four dimensions. Ranking was sequential, i.e., to determine the n^th^ ranked policy option all policy options already ranked 1 to n-1 were removed from the comparison. Note that whilst this provides a fair ‘head-to-head’ comparison among the remaining policy options, caution is required in interpreting the *incremental* cost-effectiveness for the next top-ranked policy compared to policies already ranked and removed. For example, policies including masks often followed the same policy without masks in ranking, yet the incremental cost effectiveness of adding masks was poor. For key policy comparisons, we also calculated incremental cost-effectiveness ratios (ICERs).

### Uncertainty and sensitivity analyses

We generated tornado plots showing the variation in model outputs when comparing the lowest and highest quintiles of key input parameters. Additionally, we ran sensitivity analyses using an alternative discounting approach recommended by the UK Treasury^18^ (1.5% for HALYs and 3.5% for costs), adjusting the WTP per HALY to $140,000 and $35,000, and assuming people dying of COVID-19 have 1.5 times the mortality and 1.25 times the morbidity of the average person of the same sex and age.

## Results

### Model validation

The mean number of infections generated by the model from 1 April to 30 September 2022 was 3.2 million (median 3.4m, interquartile range [IQR] 2.5 to 4.1m). The number of reported cases in Victoria in the same period was 1.3 million,^19^ the same as the mean number of symptomatic infections output by the model. The mean number of COVID-19-related deaths was 2,300 (median 2,200, IQR 1,400 to 3,000) compared to 2,900 recorded in Victoria in the same period.^20^

### Health and cost impacts of modelled policies

Across the 936 policy-by-variant scenarios the mean number of infections, hospitalisations and deaths over the 12 months from October 2022 output by the model were 4.2 million, 34,900, and 8,100 respectively.

Figure 1 shows cumulative infections, hospitalisations and deaths over the 12-month period for the modelled policy options across the nine SARS-CoV-2 variant scenarios. For ease of interpretation, each outcome is averaged over the four mask policies given their modest impact on health outcomes; results for these and other key model outputs across all 936 scenarios are shown as heatmaps in Supplementary Figures 1 to 10. Mean total infections ranged from 1.8 million (an antigenically Omicron-like variant with low immune escape and high virulence, with higher stringency PHSMs) to 6.1 million (an antigenically novel variant with low virulence and high immune escape, with lower stringency PHSMs).

**Figure 1:**
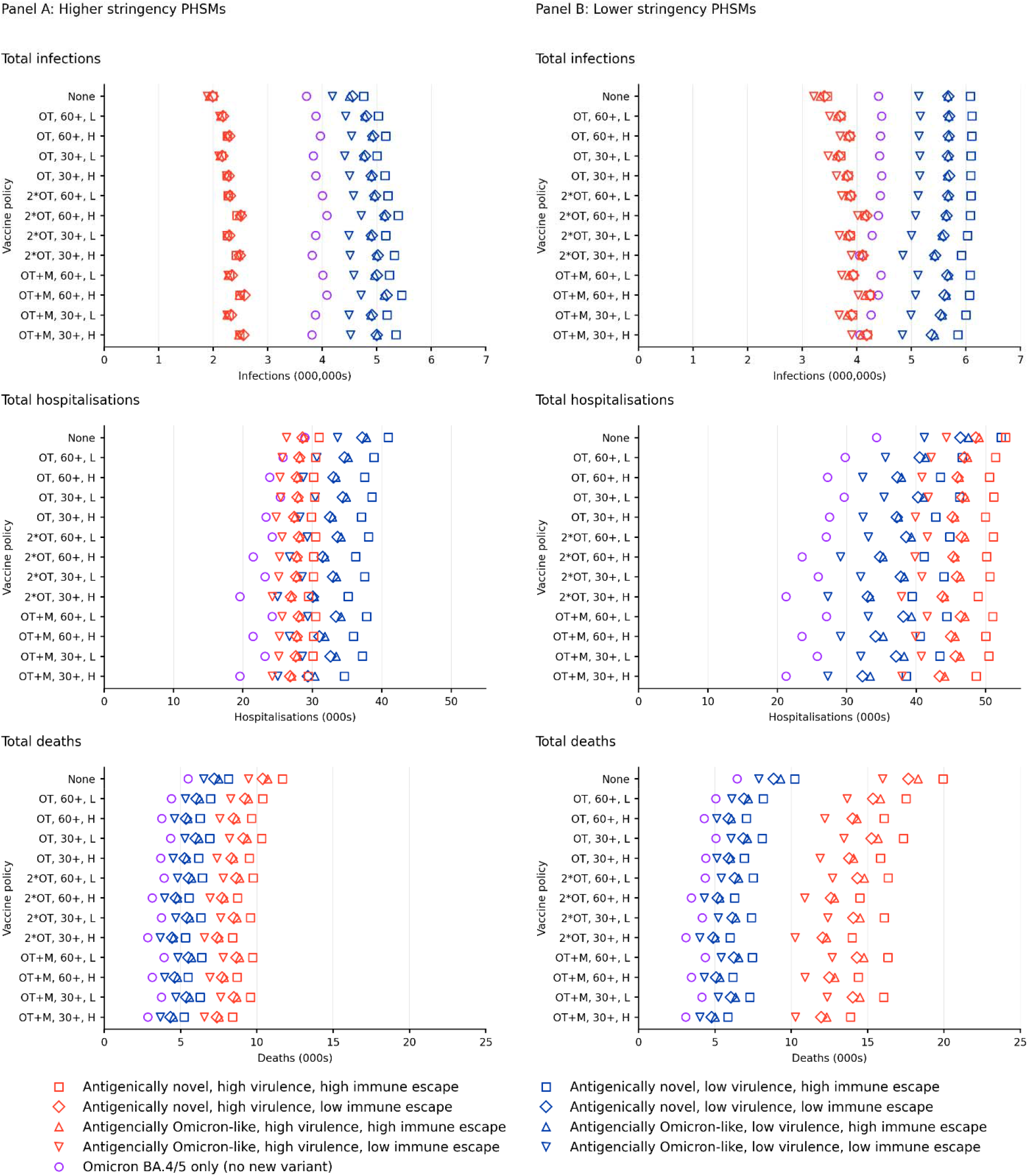
Mean cumulative infections, hospitalisations and deaths over 12 months for packages of policy options and nine future SARS-CoV-2 variant scenarios, averaged across mask policies. OT: Omicron-targeted vaccine in Q4 2022; 2*OT: Omicron-targeted vaccines in Q4 2022 and Q2 2023; OT+M: Omicron-targeted vaccine in Q4 2022 and multivalent vaccine in Q2 2023; 30+: administered to people aged ≥30 years; 60+ administered to people aged ≥60 years; H: high coverage; L: low coverage

More stringent PHSMs reduced cumulative infections, hospitalisations and deaths by an average of 25%, 24% and 24% across 468 comparisons with policies containing less stringent PHSMs (other policies and variant scenarios held constant). Any vaccination schedule from October 2022, compared to no further vaccination, reduced hospitalisations by an average of 12% and deaths by 27% (but resulted in a 10% increase in infections as more transmission is tolerated within the hospitalisation thresholds used to set PHSM stages). Additional vaccination of 30-to 59-year-olds, compared to just vaccinating ≥60-year-olds, reduced hospitalisations and deaths by 3%, and resulted in a 2% reduction in infections. High versus low vaccination coverage reduced deaths and hospitalisations on average by 15% and 6%, but also resulted in a 3% increase in average infections. The mask promotion policy (leading to increased mask wearing) and respirator substitution policy (leading to 80% compared to 20% of mask wearers wearing respirators, but no overall increase in mask wearing) both applied during large outbreaks led to 1% to 2% decreases in cumulative infections, hospitalisations and deaths.

Figure 2 similarly shows HALYs lost, net health expenditure and GDP loss for the 12 months following October 2022. More stringent PHSMs gained 13,400 HALYs on average compared to less stringent PHSMs, across comparisons of scenarios varying only by PHSM policy (3,390 HALYs gained on average in the setting of low virulence new variants, 26,200 for high virulence variants). Any vaccination schedule, compared with no further vaccination, gained 6,370 HALYs on average. The mask promotion and respirator substitution policies led to an average of 840 and 650 HALYs gained, respectively. Net health expenditure predictably increased when moving from nil further vaccination to vaccination with increasing levels of population vaccination coverage. Net health expenditure varied substantially by emergent SARS-CoV-2 variant when PHSMs were less stringent. GDP losses also varied widely across variant scenarios and were 482% greater on average for the higher stringency PHSM strategy.

**Figure 2:**
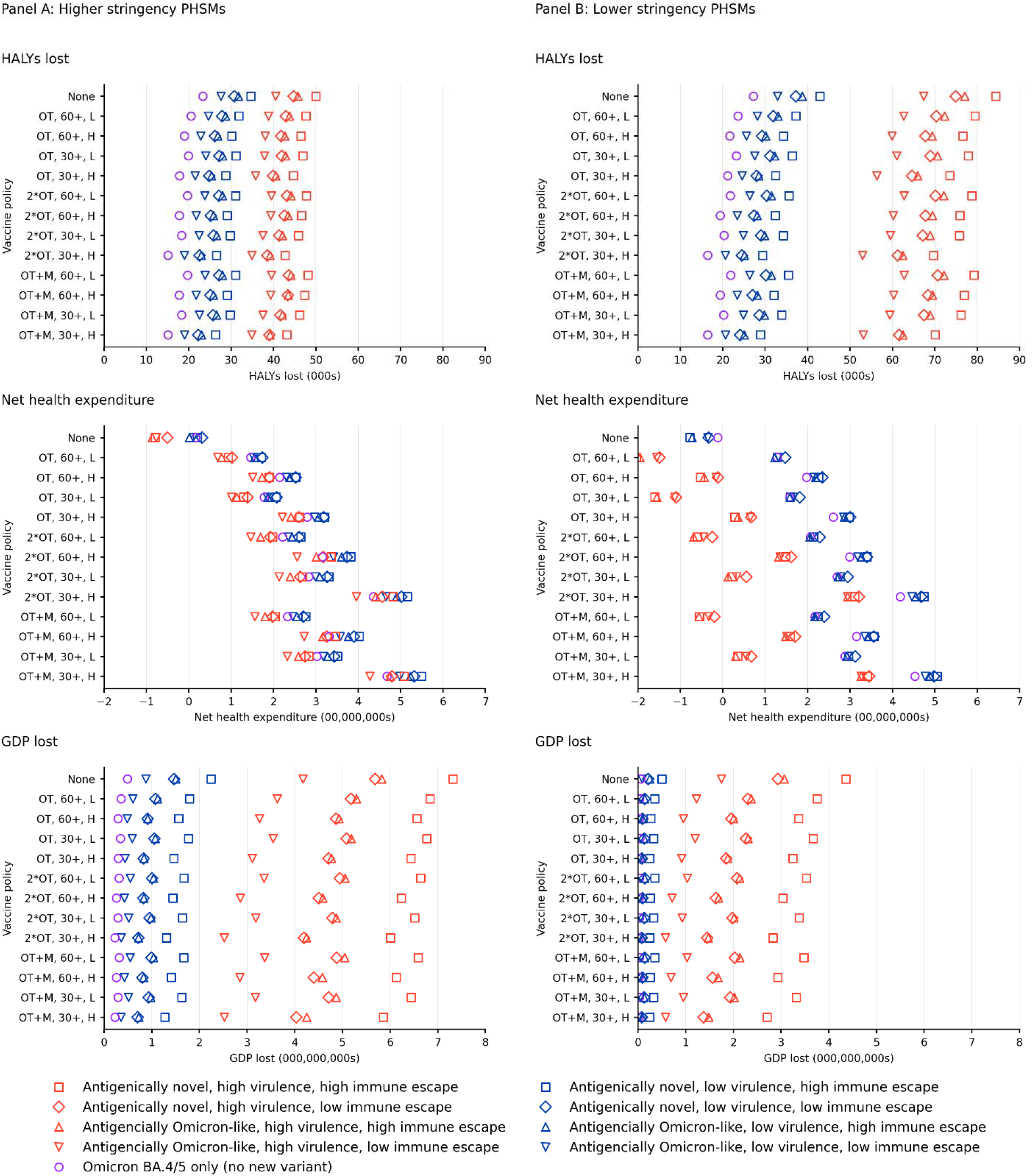
Lifetime HALY loss, health expenditure and GDP loss over 12 months for packages of policy options and nine future SARS-CoV-2 variant scenarios, averaged across mask policies. OT: Omicron-targeted vaccine in Q4 2022; 2*OT: Omicron-targeted vaccines in Q4 2022 and Q2 2023; OT+M: Omicron-targeted vaccine in Q4 2022 and multivalent vaccine in Q2 2023; 30+: administered to people aged ≥30 years; 60+ administered to people aged ≥60 years; H: high coverage; L: low coverage

Time spent in stage ≥2 PHSMs, the number of days in which hospitals had >750 or >1500 COVID-19 patients admitted, NMB and health expenditure due to and excluding deaths incurred are shown in Supplementary Figures 11 to 14. Supplementary Figures 15 and 16 show model outputs with nil or both mask policies active. More stringent PHSMs increased the days in stage 3 or greater by an average of 42 days (23 days for low virulence new variants, 70 days for high virulence variants). Any vaccination decreased days in stage 3 or greater by an average of 15 days (36%). In this dynamic model where the stage of PHSMs is set to keep hospitalisations beneath a target, the societal gain from high versus low vaccination coverage manifested as 20% fewer days with stage 3 or greater restrictions. The number of days with >750 COVID-19 patients admitted was an average of 82 days (54%) less under the more stringent compared to less stringent PHSM policy. The mask policies, applied during large outbreaks, reduced days with >750 COVID-19 patients in hospital by 3 to 4 days (2%) across all comparisons, but for high virulence variants the mask policies reduced days with >750 hospitalisations by 8 to 10 days (4% to 5%).

### Incremental cost effectiveness ratios

Mean HALY losses and net health expenditure changes can be used to calculate ICERs. For example, Supplementary Figure 17 shows ICERs for every head-to-head comparison of any ongoing vaccination schedule incremental to no further vaccination (from a health system perspective). The mean ICER for any vaccine schedule for ≥60-year-olds, averaged across all 432 possible incremental comparisons with nil further vaccination, was $77,500 per HALY gained (median $58,800, 95% range $33,800 to $248,000). The mean ICER for additionally vaccinating 30-to 59-year-olds was $41,600 (median $42,700, 95% range $22,700 to $65,700; generated based on the difference in health expenditure and HALYs for vaccinating all ≥30-year-olds compared to just ≥60-year-olds). Considered in isolation, these ICERs for vaccination may exceed a funder’s usual WTP. However, the benefits of vaccination for society also manifest as reduced societal costs such as time under higher stages of PHSMs, and lesser GDP loss. Accordingly, from a health plus GDP perspective any vaccination was usually cost saving (Supplementary Figure 18).

More stringent PHSMs, compared to less stringent, were dominant (i.e., cost saving and resulting in health gain) from a health system only perspective for 28% of the 468 comparisons where other policies and variant characteristics were the same. Of the remaining 72% of comparisons, the average cost was $14,500 per HALY gained from a health system-only perspective (maximum $41,800). However, from a health plus GDP perspective the average cost was $186,000 per HALY gained (median $136,000, 95% range $94,000 to $387,000). Increasing mask use when in stages ≥3 (from, for example, 45% to 62% among people aged 20 to 49 years in stage 3), was usually dominant from both health and health plus GDP perspectives, despite not dramatically reducing cumulative hospitalisations and deaths. This policy was assumed to be achieved through media promotion campaigns only, costing approximately $15,000 per day. The provision of respirators during large outbreaks by the government was not cost effective from either a health or health plus GDP perspective.

### Optimal ranking using net monetary benefit, hospital occupancy and deaths

As evident above, there are many separate policy considerations and interactions that make decision-making complex and difficult during a pandemic. Therefore, Figure 3 presents policies ranked by NMB from health system and health system plus GDP perspectives, high hospital occupancy, and deaths. Weighting these four measures evenly, the highest ranked policy was more stringent PHSMs, two further vaccine doses (an Omicron-targeted vaccine followed by a multivalent vaccine) for ≥30-year-olds with high coverage, and promotion of increased mask wearing (but not government provision of respirators). The top ten ranked policies usually included two further rounds of vaccination, vaccinating ≥30-year-olds with high coverage, and higher stringency PHSMs with increased overall mask wearing during large outbreaks. Note that whilst interventions with the respirator provision policy did occur in the top ten, they only did so after removing the same intervention without respirators; the ICERs discussed above indicate that a policy of government provision of respirators is not incrementally cost effective.

**Figure 3:**
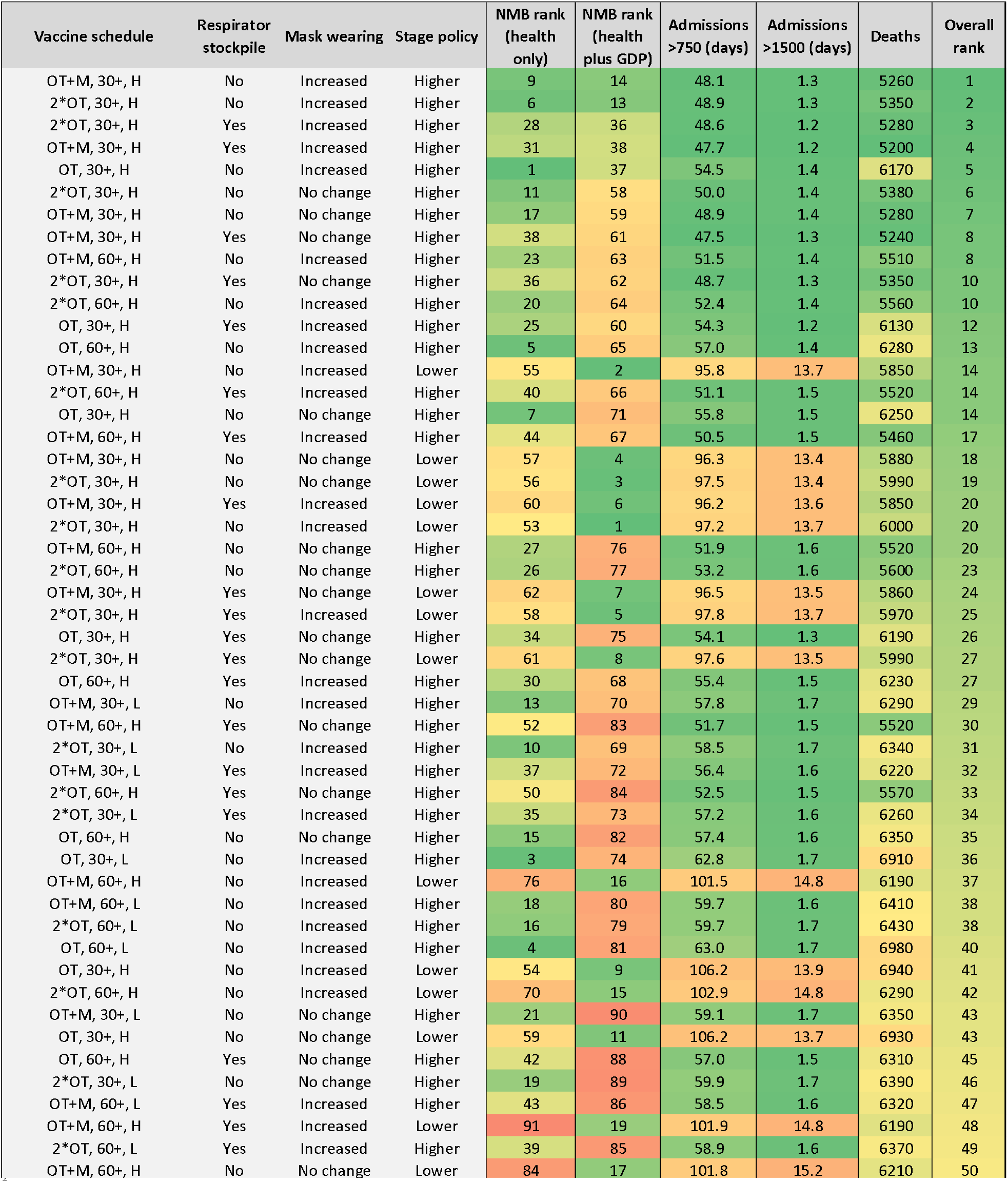
Heat map for policy options (over 12 months, considering all nine future SARS-CoV-2 scenarios equally likely) of their: rank in net monetary benefit (with a health-adjusted life year valued at AUD 70,000 = USD 50,000) from both health system and health system plus GDP perspectives^†^; number of days that >750 and >1500 people were in hospital due to COVID-19; cumulative deaths; and combined rank^€^. ^†^Using a 3% discount rate for both HALYs and costs. This ranking used sequential net monetary benefit (NMB) analyses, whereby the proportion of times each of the 104 policy options had the highest NMB across the 500 iterations (i.e., pooling the nine SARS-CoV-2 strata, meaning they were equally weighted in likelihood) was determined. The top ranked policy was selected and put aside. The remaining 103 policies were re-analyzed, the now remaining top-ranked policy identified and put aside. This ranking was repeated until there was only one (least optimal) policy left. ^€^ For each of the 104 policies the average rank of the ranking according to (a) NMB from a health system perspective, (b) the ranking according to NMB from a health plus GDP system perspective, (c) the ranking of the average of the ranking of days with >750 and > 1500 people in hospital, and (d) the ranking of deaths was calculated. Note, this inherently weights these four constructs equally in selecting the overall optimal policy; different decision makers wish to apply different weights across these four metrics. Policies ranking in the top 50 are presented here. OT: Omicron-targeted vaccine in Q4 2022; 2*OT: Omicron-targeted vaccines in Q4 2022 and Q2 2023; OT+M: Omicron-targeted vaccine in Q4 2022 and multivalent vaccine in Q2 2023; 30+: administered to people aged ≥30 years; 60+ administered to people aged ≥60 years; H: high coverage; L: low coverage

The rankings in Figure 3 consider each of the nine future SARS-CoV-2 variant scenarios equally likely; for users that wish to weight differently the likelihood of future variants (e.g., the next major variant being more likely to possess a high level of additional immune escape capacity) or alter the weighting of the four criteria, Figure 3 can be reproduced with these alternative settings at an interactive online tool.^21^

### Sensitivity analyses

Supplementary Figure 19 shows the ranking as above, and for sensitivity analyses using UK Treasury-recommended discount rates (1.5% per annum for HALYs, 3.5% per annum for costs), HALYs valued at $35,000 and $140,000, and assuming people dying of COVID-19 have 1.5 times the mortality and 1.25 times the morbidity of the average person of the same sex and age (as opposed to 2 and 1.5 times respectively as used in the main analysis). Rankings were similar across these sensitivity analyses.

The tornado plots shown in Supplementary Figure 20 indicate that uncertainty regarding the transmission potential of asymptomatic compared to symptomatic individuals drove significant amounts of uncertainty across multiple model outputs. Uncertainty regarding the waning of immunity following vaccination or infection was responsible for much of the uncertainty around cumulative hospitalisations and deaths.

## Discussion

Decision makers increasingly require frameworks to systematically weigh up the costs and benefits of pandemic policy choices.^2^ A growing number of recent publications integrate economic evaluation with epidemiologic modelling in an attempt to facilitate decision making in the context of the COVID-19 pandemic.^22-24^ However, none have examined combined interventions, accounted for SARS-CoV-2 evolution, accurately represented waning immunity, and included both acute COVID-19 and long COVID morbidity simultaneously. Our model addresses all these imperatives.

This study examines 936 future scenarios, formed by cross-classifying 104 policy options with nine future variant scenarios. Irrespective of the policies implemented, significant ongoing SARS-CoV-2-related morbidity and mortality is predicted by this model in the coming months and years; control over cumulative viral transmission was limited given the scenario specifications. Nevertheless, meaningful policy recommendations still emerge from this analysis.

Firstly, we found that higher stringency PHSMs tended to perform better when outcomes were assessed from a health system perspective, but this was often in conflict with findings when GDP losses were considered. This conflict of perspectives makes explicit the tension between protecting public health and protecting the economic interests of society at this stage of the pandemic. Earlier in the pandemic, in the absence of vaccines and especially for countries pursuing a zero-COVID strategy, optimal policies from health and societal perspectives were more clearly aligned.^17^ This tradeoff between health and societal criteria emphasises the importance of developing explicit frameworks such as ours to manage these competing interests. Overall, when considering cost effectiveness, hospital system pressure and population mortality, lower thresholds for escalating restrictions (i.e., higher stringency PHSMs) consistently performed better despite their associated economic trade-offs.

Secondly, government provision of respirators to the public (to use as an alternative to surgical or cloth masks, increasing respirator use from 20% to 80% of mask-wearers) and increases in mask wearing once large outbreaks had already occurred only had modest effects on morbidity and mortality. A likely reason for the limited impact of these policies on health outcomes is that during infection peaks (when these policies were activated) many people are confined at home where masks are not worn. Another reason may be the now high innate transmissibility of SARS-CoV-2 variants; masks reduce the immediate risk of transmission,^5^ but do not make it zero. This may serve more to increase the number of days to infection or delay the interval between infections rather than prevent it altogether. Delay of infection may still be useful to ‘flatten’ the epidemic curve and protect health services from being overwhelmed. Indeed, reduced days with hospital capacity exceeding thresholds of >750 and >1,500 COVID-19 patients were seen when respirator substitution and increased mask wearing policies were active, although these reductions were modest. We are exploring further these short- and long-term tradeoffs of mask wearing elsewhere, including through modelling increased mask-wearing at all times rather than just during surges of infection.

Third, the provision of any vaccine booster was consistently seen to be more beneficial than not providing ongoing vaccination. While differences in the vaccine schedules modelled were not dramatic when considering individual model outputs in isolation, and the incremental cost effectiveness ratios of vaccination policies from a health system only perspective were often at the threshold of what funders might be prepared to pay per HALY gained, when also considering societal gains (i.e., reduced time in higher stage PHSMs) our policy ranking results suggest that ongoing regular vaccination should continue to play a key role in the pandemic response despite the associated financial costs. Of note, vaccinating people aged 30 years and over appeared to be more optimal than targeted vaccination of people aged ≥60 – at least within our model that had dynamic PHSMs in response to hospital occupancy.

Our modelling parameterized Omicron-targeted vaccines as having twice the VE on the odds scale against Omicron BA.1/2 (and antigenically similar variants) compared to first-generation mRNA vaccines (e.g., BNT162b and mRNA1273). This is equivalent to increasing peak VE for agents in the model against any infection from 51.6% (the value we use at two weeks post second dose for younger adults; see Appendix for details) to 68.0%, or VE against death from 96.9% to 98.4%. These ratio increases in VE are supported by preliminary serologic data for bivalent ancestral- and Omicron-targeted vaccines, compared to ancestral-targeted vaccine only.^25^ Our model also includes the same waning of protection over time for first- and next-generation vaccines, and a 50% higher unit cost for multivalent vaccines compared to both first-generation and Omicron-targeted vaccines. Whilst these seem reasonable assumptions at the time of writing, it will be important to revise these assumptions as with updated estimates of expected VE and waning (e.g., based on in vitro antibody titers^26,27^ or, ideally, real-world VE studies) and updated costs. Such model flexibility, augmented by both comprehensive surveillance systems monitoring SARS-CoV-2 variant emergence and close links to vaccine producers with the capacity to rapidly deliver new vaccines, suggests a fruitful policy pathway to better population health outcomes over the remainder of this (and future) pandemics.

In comparison to similar models our framework has many advantages. We simulated combined interventions to reflect the fact that policy choices are not made in isolation, modelled viral evolution, developed a novel method to quantify long COVID morbidity, and accounted for acute COVID-19, long COVID and intervention costs in addition to future health expenditure and the economic consequences of PHSMs. Uniquely, our model also includes a data-driven representation of waning protection following vaccination, previous infection, or both, in contrast to most previously published COVID-19 transmission models.^28^ While there are substantial gaps in the COVID-19 literature (e.g., regarding the risk and symptom profile of long COVID, the waning of natural infection-derived immunity, the effect of immunity on onward transmission once infected, and the infectiousness of asymptomatic individuals), we incorporated generous uncertainty in model inputs, reflect the impact of uncertainty in our results, and still find important differences that lead to certain policies being quantified as more optimal. Sensitivity analyses provide insights into priority areas for research to better parameterise future models, including the need to develop a greater understanding of the transmission potential of asymptomatic compared to symptomatic individuals and the kinetics of immunity waning following vaccination or infection.

Our model only allows for the emergence of one new variant during the 12-month period from October 2022. Future modelling could allow for important new variants emerging more frequently, but (if using our modelling framework) this will increase the number of SARS-CoV-2 variant scenarios and make the results more challenging to summarize and interpret. Future modelling should also include sequelae other than the long COVID symptoms accounted for here (e.g., post-acute cardiovascular complications of COVID-19) as this evidence base improves,^29^ and would likely benefit from increased consideration of population heterogeneity (such as the distribution of underlying medical comorbidities, for example) and indirect effects of COVID-19 on the health system (such as impacting access to routine health services). Finally, a limited number of policy options were considered. It is important to recognise that minor adjustments in these policies (e.g., altering the baseline use of masks at all stages, changing vaccine schedules, modifying the cost or type of respirators modelled, or adding emerging interventions such as antiviral medications) could significantly alter the results. Modelled policies should be refined as policy discussions in the Australian and international contexts further develop to ensure relevance; for example, guidance regarding vaccination of young people may change as information regarding the risks and benefits of vaccinating this population evolves, or there may be a reluctance in future for governments to impose the kinds of restrictions included in stages 4 and 5 of the PHSMs modelled here. Coding differences between jurisdictions also mean that COVID-19-related hospitalisations, which are used as triggers for moving through PHSM stages in this model, should be locally contextualised.

Decision making during the COVID-19 pandemic is challenging and requires consideration of the costs and benefits of interventions in an increasingly complex policy environment. This model demonstrates that in the absence of, for example, a new vaccine associated with substantially less VE waning or improved neutralising protection against infection, the health system in Victoria – and similar jurisdictions internationally – should be prepared for significant ongoing COVID-19-related morbidity and mortality over the next 12 months. It reaffirms the importance of regular COVID-19 vaccination and PHSMs as key tools in the ongoing pandemic response. Crucially, this modelling provides a framework that can be rapidly updated to systematically compare the health and economic benefits and burdens of COVID-19 policy options despite a highly uncertain future.

## Data Availability

The data analysis protocol is available online at https://mspgh.unimelb.edu.au/research-groups/centre-for-epidemiology-and-biostatistics-research/population-interventions/protocols. All model inputs are detailed in the appendix. Detailed output data is provided in the appendix and the accompanying online tool. Requests for additional model output data may be granted upon reasonable request to the researchers. Model code is available at https://github.com/population-interventions/CovidABM.

## Acknowledgements

We acknowledge funding from an anonymous philanthropist and the contributions of Jason Thompson to earlier versions of the modelling that this paper builds on. We also acknowledge the work of Natalie Carvalho and Patrick Abraham for assistance with costing and Courtney Gee, Zainab Albadri and Samantha Grimshaw for assistance with data collection. This research was supported by The University of Melbourne’s Research Computing Services and the Petascale Campus Initiative.

## Author contributions

JS – conceptualization, investigation, methodology, project administration, supervision, writing – original draft, writing – review and editing

TW – conceptualization, data curation, formal analysis, investigation, methodology, software, writing – original draft, writing – review and editing

SH – conceptualization, data curation, formal analysis, investigation, methodology, writing – original draft, writing – review and editing

JZ – conceptualization, investigation, methodology, writing – review and editing

HA – data curation, formal analysis, visualization, software, writing – review and editing SR – methodology, formal analysis, writing – review and editing

TB – conceptualization, formal analysis, funding acquisition, investigation, methodology, project administration, supervision, writing – original draft, writing – review and editing.

