## Supplementary materials for "Epidemiologic and economic modelling of optimal COVID-19 policy: public health and social measures, masks and vaccines in Victoria, Australia"

### Supplementary Results

#### Supplementary Figure 1: Heat map of mean <u>infections</u> for each of 936 scenarios over a 12 month timeline (millions)

| PHSMs | | | Higher Stringency | | | | | | | | | | | | | Lower Stringency | | | | | | | | | | | | |
|---|---|---|---|---|---|---|---|---|---|---|---|---|---|---|---|---|---|---|---|---|---|---|---|---|---|---|---|---|
| Vaccine schedule | | | OT+M | | | | 2*OT | | | | OT | | | | Nil | OT+M | | | | 2*OT | | | | OT | | | | Nil |
| Vaccine age group | | | 30+ | | 60+ | | 30+ | | 60+ | | 30+ | | 60+ | | | 30+ | | 60+ | | 30+ | | 60+ | | 30+ | | 60+ | | |
| Vaccine uptake | | | *H* | *L* | *H* | *L* | *H* | *L* | *H* | *L* | *H* | *L* | *H* | *L* | | *H* | *L* | *H* | *L* | *H* | *L* | *H* | *L* | *H* | *L* | *H* | *L* | |
| **Variant** | **Respirator stockpile** | **Mask wearing** | | | | | | | | | | | | | | | | | | | | | | | | | | |
| Omicron BA.4/5 only (no new variant) | Yes | High | 3.80 | 3.86 | 4.07 | 4.00 | 3.81 | 3.87 | 4.07 | 3.99 | 3.86 | 3.81 | 3.95 | 3.87 | 3.68 | 4.05 | 4.26 | 4.39 | 4.45 | 4.05 | 4.28 | 4.40 | 4.43 | 4.45 | 4.42 | 4.43 | 4.46 | 4.39 |
| | Yes | Low | 3.81 | 3.88 | 4.09 | 4.01 | 3.81 | 3.89 | 4.09 | 4.01 | 3.89 | 3.85 | 3.97 | 3.88 | 3.71 | 4.06 | 4.26 | 4.40 | 4.44 | 4.05 | 4.28 | 4.40 | 4.43 | 4.46 | 4.42 | 4.44 | 4.45 | 4.41 |
| | No | High | 3.82 | 3.88 | 4.09 | 4.01 | 3.82 | 3.87 | 4.09 | 4.00 | 3.88 | 3.84 | 3.96 | 3.89 | 3.71 | 4.06 | 4.27 | 4.40 | 4.45 | 4.05 | 4.29 | 4.40 | 4.44 | 4.46 | 4.42 | 4.44 | 4.46 | 4.40 |
| | No | Low | 3.82 | 3.88 | 4.10 | 4.03 | 3.83 | 3.90 | 4.10 | 4.01 | 3.90 | 3.86 | 3.98 | 3.90 | 3.74 | 4.06 | 4.27 | 4.39 | 4.45 | 4.05 | 4.28 | 4.40 | 4.43 | 4.46 | 4.42 | 4.44 | 4.46 | 4.40 |
| Antigenically Omicron-like, low immune escape, low virulence | Yes | High | 4.51 | 4.47 | 4.69 | 4.54 | 4.49 | 4.47 | 4.69 | 4.54 | 4.48 | 4.37 | 4.50 | 4.39 | 4.14 | 4.84 | 4.99 | 5.08 | 5.12 | 4.84 | 5.00 | 5.08 | 5.13 | 5.15 | 5.15 | 5.14 | 5.16 | 5.13 |
| | Yes | Low | 4.52 | 4.50 | 4.72 | 4.60 | 4.52 | 4.49 | 4.73 | 4.58 | 4.50 | 4.42 | 4.54 | 4.44 | 4.19 | 4.84 | 5.00 | 5.07 | 5.12 | 4.85 | 5.01 | 5.07 | 5.13 | 5.16 | 5.16 | 5.15 | 5.16 | 5.14 |
| | No | High | 4.53 | 4.48 | 4.71 | 4.58 | 4.52 | 4.49 | 4.71 | 4.57 | 4.51 | 4.41 | 4.54 | 4.44 | 4.18 | 4.84 | 4.99 | 5.08 | 5.12 | 4.85 | 5.01 | 5.08 | 5.13 | 5.16 | 5.16 | 5.14 | 5.17 | 5.14 |
| | No | Low | 4.52 | 4.51 | 4.73 | 4.60 | 4.52 | 4.51 | 4.73 | 4.60 | 4.53 | 4.45 | 4.56 | 4.46 | 4.24 | 4.84 | 5.00 | 5.07 | 5.13 | 4.85 | 5.00 | 5.08 | 5.13 | 5.16 | 5.16 | 5.14 | 5.17 | 5.14 |
| Antigenically novel, low immune escape, low virulence | Yes | High | 4.97 | 4.88 | 5.15 | 4.95 | 4.97 | 4.87 | 5.12 | 4.91 | 4.85 | 4.73 | 4.89 | 4.75 | 4.48 | 5.37 | 5.54 | 5.61 | 5.66 | 5.44 | 5.59 | 5.65 | 5.67 | 5.69 | 5.66 | 5.68 | 5.69 | 5.68 |
| | Yes | Low | 5.01 | 4.92 | 5.17 | 5.02 | 5.04 | 4.92 | 5.16 | 4.97 | 4.91 | 4.79 | 4.94 | 4.83 | 4.57 | 5.38 | 5.54 | 5.62 | 5.67 | 5.43 | 5.60 | 5.65 | 5.68 | 5.69 | 5.67 | 5.69 | 5.70 | 5.68 |
| | No | High | 4.99 | 4.90 | 5.19 | 5.01 | 5.02 | 4.90 | 5.15 | 4.97 | 4.90 | 4.78 | 4.94 | 4.80 | 4.55 | 5.38 | 5.54 | 5.60 | 5.65 | 5.44 | 5.59 | 5.65 | 5.68 | 5.70 | 5.67 | 5.69 | 5.70 | 5.67 |
| | No | Low | 5.02 | 4.94 | 5.21 | 5.04 | 5.05 | 4.94 | 5.19 | 5.02 | 4.96 | 4.84 | 4.97 | 4.85 | 4.61 | 5.38 | 5.54 | 5.61 | 5.66 | 5.44 | 5.58 | 5.64 | 5.67 | 5.70 | 5.68 | 5.69 | 5.69 | 5.68 |
| Antigenically Omicron-like, high immune escape, low virulence | Yes | High | 4.98 | 4.84 | 5.10 | 4.90 | 4.96 | 4.85 | 5.10 | 4.90 | 4.85 | 4.70 | 4.87 | 4.71 | 4.43 | 5.43 | 5.57 | 5.64 | 5.68 | 5.42 | 5.57 | 5.63 | 5.68 | 5.66 | 5.67 | 5.68 | 5.68 | 5.66 |
| | Yes | Low | 5.03 | 4.91 | 5.15 | 4.98 | 5.01 | 4.91 | 5.15 | 4.97 | 4.91 | 4.78 | 4.92 | 4.80 | 4.52 | 5.43 | 5.57 | 5.65 | 5.68 | 5.42 | 5.57 | 5.65 | 5.69 | 5.66 | 5.68 | 5.69 | 5.69 | 5.69 |
| | No | High | 5.00 | 4.89 | 5.13 | 4.96 | 5.01 | 4.89 | 5.13 | 4.95 | 4.91 | 4.75 | 4.92 | 4.79 | 4.48 | 5.42 | 5.57 | 5.65 | 5.69 | 5.44 | 5.57 | 5.64 | 5.68 | 5.67 | 5.68 | 5.70 | 5.69 | 5.67 |
| | No | Low | 5.02 | 4.94 | 5.17 | 4.99 | 5.02 | 4.93 | 5.18 | 5.00 | 4.94 | 4.82 | 4.95 | 4.83 | 4.56 | 5.43 | 5.57 | 5.65 | 5.68 | 5.43 | 5.58 | 5.64 | 5.68 | 5.67 | 5.69 | 5.68 | 5.70 | 5.67 |
| Antigenically novel, high immune escape, low virulence | Yes | High | 5.30 | 5.12 | 5.41 | 5.15 | 5.26 | 5.09 | 5.34 | 5.11 | 5.08 | 4.92 | 5.09 | 4.94 | 4.67 | 5.85 | 6.00 | 6.07 | 6.07 | 5.91 | 6.04 | 6.08 | 6.10 | 6.08 | 6.10 | 6.10 | 6.11 | 6.06 |
| | Yes | Low | 5.35 | 5.21 | 5.47 | 5.25 | 5.34 | 5.19 | 5.40 | 5.23 | 5.17 | 5.03 | 5.18 | 5.06 | 4.78 | 5.86 | 6.00 | 6.07 | 6.07 | 5.92 | 6.04 | 6.10 | 6.10 | 6.09 | 6.10 | 6.12 | 6.12 | 6.10 |
| | No | High | 5.36 | 5.19 | 5.45 | 5.24 | 5.33 | 5.15 | 5.39 | 5.20 | 5.14 | 5.00 | 5.15 | 5.02 | 4.75 | 5.85 | 5.99 | 6.06 | 6.09 | 5.93 | 6.02 | 6.09 | 6.09 | 6.09 | 6.09 | 6.10 | 6.12 | 6.09 |
| | No | Low | 5.39 | 5.25 | 5.50 | 5.30 | 5.36 | 5.22 | 5.45 | 5.27 | 5.22 | 5.08 | 5.24 | 5.09 | 4.85 | 5.88 | 6.02 | 6.08 | 6.09 | 5.94 | 6.04 | 6.10 | 6.10 | 6.09 | 6.12 | 6.12 | 6.12 | 6.11 |
| Antigenically Omicron-like, low immune escape, high virulence | Yes | High | 2.40 | 2.17 | 2.42 | 2.18 | 2.40 | 2.17 | 2.43 | 2.18 | 2.16 | 2.02 | 2.18 | 2.04 | 1.81 | 3.88 | 3.62 | 3.99 | 3.66 | 3.87 | 3.63 | 3.98 | 3.67 | 3.59 | 3.42 | 3.64 | 3.45 | 3.14 |
| | Yes | Low | 2.50 | 2.28 | 2.54 | 2.30 | 2.51 | 2.27 | 2.54 | 2.30 | 2.26 | 2.13 | 2.30 | 2.14 | 1.92 | 3.92 | 3.70 | 4.04 | 3.74 | 3.91 | 3.69 | 4.04 | 3.73 | 3.64 | 3.49 | 3.72 | 3.52 | 3.24 |
| | No | High | 2.47 | 2.24 | 2.51 | 2.26 | 2.47 | 2.23 | 2.50 | 2.26 | 2.23 | 2.09 | 2.26 | 2.10 | 1.88 | 3.91 | 3.67 | 4.01 | 3.72 | 3.90 | 3.68 | 4.00 | 3.72 | 3.63 | 3.47 | 3.68 | 3.50 | 3.21 |
| | No | Low | 2.55 | 2.33 | 2.60 | 2.36 | 2.56 | 2.33 | 2.59 | 2.36 | 2.32 | 2.18 | 2.34 | 2.20 | 1.98 | 3.93 | 3.72 | 4.06 | 3.78 | 3.93 | 3.73 | 4.06 | 3.77 | 3.67 | 3.53 | 3.74 | 3.56 | 3.27 |
| Antigenically novel, low immune escape, high virulence | Yes | High | 2.44 | 2.22 | 2.45 | 2.23 | 2.38 | 2.18 | 2.39 | 2.20 | 2.17 | 2.06 | 2.19 | 2.07 | 1.89 | 4.11 | 3.82 | 4.17 | 3.84 | 4.04 | 3.78 | 4.10 | 3.81 | 3.75 | 3.59 | 3.78 | 3.60 | 3.31 |
| | Yes | Low | 2.57 | 2.36 | 2.60 | 2.37 | 2.51 | 2.32 | 2.53 | 2.34 | 2.31 | 2.20 | 2.33 | 2.21 | 2.01 | 4.21 | 3.92 | 4.27 | 3.96 | 4.13 | 3.88 | 4.20 | 3.91 | 3.86 | 3.69 | 3.90 | 3.72 | 3.43 |
| | No | High | 2.55 | 2.30 | 2.56 | 2.33 | 2.48 | 2.28 | 2.49 | 2.29 | 2.28 | 2.15 | 2.28 | 2.16 | 1.98 | 4.17 | 3.89 | 4.24 | 3.93 | 4.10 | 3.85 | 4.17 | 3.88 | 3.82 | 3.66 | 3.86 | 3.68 | 3.40 |
| | No | Low | 2.65 | 2.43 | 2.68 | 2.44 | 2.59 | 2.40 | 2.61 | 2.41 | 2.37 | 2.26 | 2.40 | 2.28 | 2.08 | 4.24 | 3.97 | 4.32 | 4.01 | 4.18 | 3.94 | 4.25 | 3.97 | 3.90 | 3.75 | 3.94 | 3.77 | 3.49 |
| Antigenically Omicron-like, high immune escape, high virulence | Yes | High | 2.35 | 2.15 | 2.36 | 2.16 | 2.35 | 2.15 | 2.35 | 2.16 | 2.15 | 2.02 | 2.16 | 2.03 | 1.83 | 4.01 | 3.74 | 4.08 | 3.76 | 4.03 | 3.74 | 4.07 | 3.76 | 3.72 | 3.54 | 3.75 | 3.55 | 3.23 |
| | Yes | Low | 2.49 | 2.28 | 2.51 | 2.30 | 2.49 | 2.28 | 2.51 | 2.30 | 2.28 | 2.15 | 2.29 | 2.17 | 1.95 | 4.11 | 3.85 | 4.17 | 3.88 | 4.10 | 3.85 | 4.17 | 3.88 | 3.83 | 3.65 | 3.86 | 3.67 | 3.36 |
| | No | High | 2.45 | 2.24 | 2.46 | 2.26 | 2.45 | 2.25 | 2.46 | 2.26 | 2.24 | 2.12 | 2.25 | 2.12 | 1.91 | 4.08 | 3.82 | 4.14 | 3.85 | 4.08 | 3.82 | 4.14 | 3.84 | 3.79 | 3.62 | 3.83 | 3.63 | 3.32 |
| | No | Low | 2.57 | 2.35 | 2.59 | 2.37 | 2.57 | 2.35 | 2.58 | 2.37 | 2.35 | 2.22 | 2.36 | 2.23 | 2.02 | 4.15 | 3.89 | 4.22 | 3.93 | 4.15 | 3.91 | 4.22 | 3.94 | 3.87 | 3.71 | 3.92 | 3.73 | 3.42 |
| Antigenically novel, high immune escape, high virulence | Yes | High | 2.36 | 2.17 | 2.36 | 2.18 | 2.30 | 2.13 | 2.30 | 2.15 | 2.14 | 2.03 | 2.13 | 2.04 | 1.88 | 4.08 | 3.80 | 4.11 | 3.81 | 4.00 | 3.76 | 4.03 | 3.75 | 3.73 | 3.58 | 3.75 | 3.57 | 3.34 |
| | Yes | Low | 2.52 | 2.33 | 2.53 | 2.34 | 2.46 | 2.30 | 2.47 | 2.31 | 2.28 | 2.18 | 2.29 | 2.19 | 2.03 | 4.22 | 3.96 | 4.26 | 3.96 | 4.13 | 3.92 | 4.18 | 3.92 | 3.87 | 3.74 | 3.92 | 3.75 | 3.50 |
| | No | High | 2.47 | 2.28 | 2.47 | 2.28 | 2.41 | 2.24 | 2.41 | 2.26 | 2.24 | 2.13 | 2.24 | 2.14 | 1.98 | 4.19 | 3.90 | 4.23 | 3.92 | 4.09 | 3.86 | 4.13 | 3.87 | 3.84 | 3.69 | 3.85 | 3.71 | 3.44 |
| | No | Low | 2.60 | 2.41 | 2.61 | 2.42 | 2.54 | 2.38 | 2.55 | 2.39 | 2.36 | 2.26 | 2.37 | 2.27 | 2.10 | 4.30 | 4.04 | 4.35 | 4.05 | 4.20 | 4.00 | 4.25 | 3.99 | 3.96 | 3.83 | 3.98 | 3.83 | 3.57 |

*OT: Omicron-targeted vaccine in Q4 2022; 2*OT: Omicron-targeted vaccines in Q4 2022 and Q2 2023; OT+M: Omicron-targeted vaccine in Q4 2022 and multivalent vaccine in Q2 2023; 30+: administered to people aged ≥30 years; 60+ administered to people aged ≥60 years; H: high coverage; L: low coverage*

### Supplementary Figure 2: Heat map of mean COVID-19 <u>hospitalisations</u> for each of 936 scenarios over a 12 month timeline

| | | PHSMs | Higher Stringency | | | | | | | | | | | | | Lower Stringency | | | | | | | | | | | | |
|---|---|---|---|---|---|---|---|---|---|---|---|---|---|---|---|---|---|---|---|---|---|---|---|---|---|---|---|---|
| | | Vaccine schedule | OT+M | | | | 2*OT | | | | OT | | | | Nil | OT+M | | | | 2*OT | | | | OT | | | | Nil |
| | | Vaccine age group | 30+ | | 60+ | | 30+ | | 60+ | | 30+ | | 60+ | | | 30+ | | 60+ | | 30+ | | 60+ | | 30+ | | 60+ | | |
| | | Vaccine uptake | *H* | *L* | *H* | *L* | *H* | *L* | *H* | *L* | *H* | *L* | *H* | *L* | | *H* | *L* | *H* | *L* | *H* | *L* | *H* | *L* | *H* | *L* | *H* | *L* | |
| Variant | Respirator stockpile | Mask wearing | | | | | | | | | | | | | | | | | | | | | | | | | | |
| Omicron BA.4/5 only (no new variant) | Yes | High | 19483 | 23118 | 21374 | 24159 | 19512 | 23107 | 21413 | 24176 | 23143 | 25215 | 23774 | 25681 | 28666 | 21223 | 25747 | 23520 | 27112 | 21234 | 25857 | 23555 | 27044 | 27485 | 29655 | 27213 | 29776 | 34239 |
| | Yes | Low | 19567 | 23192 | 21476 | 24158 | 19543 | 23209 | 21491 | 24233 | 23379 | 25435 | 23908 | 25737 | 28918 | 21275 | 25737 | 23561 | 27050 | 21280 | 25886 | 23576 | 27028 | 27532 | 29646 | 27232 | 29748 | 34321 |
| | No | High | 19596 | 23182 | 21432 | 24195 | 19581 | 23145 | 21463 | 24190 | 23295 | 25413 | 23817 | 25836 | 28971 | 21251 | 25764 | 23568 | 27071 | 21258 | 25942 | 23563 | 27063 | 27512 | 29633 | 27222 | 29772 | 34295 |
| | No | Low | 19640 | 23247 | 21551 | 24343 | 19667 | 23306 | 21565 | 24273 | 23497 | 25524 | 23945 | 25862 | 29133 | 21284 | 25767 | 23532 | 27084 | 21254 | 25870 | 23571 | 27037 | 27544 | 29585 | 27284 | 29796 | 34315 |
| Antigenically Omicron-like, low immune escape, low virulence | Yes | High | 24977 | 28447 | 26601 | 29142 | 24872 | 28382 | 26594 | 29104 | 28087 | 30178 | 28541 | 30444 | 33301 | 27289 | 32012 | 29116 | 33078 | 27248 | 32000 | 29102 | 33108 | 32294 | 35360 | 32298 | 35516 | 41144 |
| | Yes | Low | 25000 | 28599 | 26709 | 29431 | 25029 | 28529 | 26765 | 29315 | 28123 | 30507 | 28668 | 30670 | 33654 | 27262 | 32055 | 29079 | 33061 | 27288 | 32009 | 29054 | 33098 | 32380 | 35381 | 32313 | 35556 | 41093 |
| | No | High | 25055 | 28544 | 26720 | 29313 | 25017 | 28571 | 26744 | 29264 | 28240 | 30430 | 28718 | 30706 | 33595 | 27259 | 31953 | 29124 | 33070 | 27250 | 32002 | 29106 | 33121 | 32314 | 35341 | 32279 | 35634 | 41199 |
| | No | Low | 25036 | 28668 | 26773 | 29459 | 25004 | 28624 | 26785 | 29392 | 28336 | 30588 | 28829 | 30788 | 33983 | 27241 | 32011 | 29077 | 33154 | 27299 | 31973 | 29134 | 33115 | 32374 | 35399 | 32300 | 35568 | 41236 |
| Antigenically novel, low immune escape, low virulence | Yes | High | 29240 | 32431 | 30802 | 33057 | 29838 | 32719 | 31262 | 33302 | 32207 | 34079 | 32777 | 34290 | 36705 | 32241 | 37128 | 34206 | 38123 | 33031 | 37813 | 34830 | 38537 | 37196 | 40206 | 37329 | 40456 | 46371 |
| | Yes | Low | 29392 | 32664 | 30976 | 33444 | 30139 | 33015 | 31571 | 33618 | 32608 | 34407 | 33093 | 34715 | 37278 | 32283 | 37102 | 34192 | 38133 | 32996 | 37777 | 34824 | 38578 | 37180 | 40263 | 37294 | 40521 | 46349 |
| | No | High | 29303 | 32596 | 31089 | 33510 | 30106 | 33002 | 31510 | 33690 | 32578 | 34403 | 33100 | 34652 | 37232 | 32252 | 37046 | 34107 | 38129 | 33026 | 37745 | 34797 | 38563 | 37217 | 40210 | 37289 | 40509 | 46309 |
| | No | Low | 29487 | 32774 | 31170 | 33599 | 30222 | 33176 | 31620 | 33905 | 32825 | 34656 | 33215 | 34896 | 37568 | 32262 | 37128 | 34099 | 38170 | 33022 | 37765 | 34742 | 38526 | 37221 | 40370 | 37252 | 40478 | 46429 |
| Antigenically Omicron-like, high immune escape, low virulence | Yes | High | 30199 | 33122 | 31633 | 33781 | 30066 | 33230 | 31580 | 33805 | 32617 | 34501 | 33183 | 34729 | 37328 | 33411 | 38275 | 35211 | 39287 | 33391 | 38297 | 35181 | 39296 | 37427 | 40995 | 37866 | 41273 | 47434 |
| | Yes | Low | 30447 | 33492 | 31804 | 34217 | 30314 | 33558 | 31843 | 34196 | 32914 | 35036 | 33398 | 35251 | 37875 | 33369 | 38300 | 35227 | 39280 | 33329 | 38302 | 35265 | 39333 | 37395 | 41059 | 37857 | 41290 | 47659 |
| | No | High | 30353 | 33422 | 31819 | 34204 | 30324 | 33454 | 31846 | 34176 | 32953 | 34885 | 33457 | 35226 | 37749 | 33350 | 38246 | 35290 | 39407 | 33404 | 38259 | 35224 | 39328 | 37468 | 41031 | 37974 | 41356 | 47540 |
| | No | Low | 30484 | 33721 | 31984 | 34364 | 30443 | 33697 | 32062 | 34436 | 33140 | 35207 | 33636 | 35476 | 38280 | 33387 | 38210 | 35304 | 39330 | 33392 | 38263 | 35274 | 39269 | 37465 | 41084 | 37876 | 41344 | 47429 |
| Antigenically novel, high immune escape, low virulence | Yes | High | 34279 | 36752 | 35644 | 37350 | 34814 | 37083 | 35911 | 37601 | 36633 | 38002 | 37089 | 38358 | 40286 | 38550 | 43399 | 40566 | 44354 | 39358 | 44085 | 41121 | 44906 | 42803 | 46277 | 43490 | 46650 | 52024 |
| | Yes | Low | 34549 | 37328 | 35991 | 37864 | 35206 | 37616 | 36276 | 38177 | 37104 | 38622 | 37590 | 38987 | 41014 | 38564 | 43385 | 40516 | 44355 | 39404 | 43958 | 41145 | 44807 | 42823 | 46195 | 43411 | 46557 | 52248 |
| | No | High | 34670 | 37273 | 35962 | 37892 | 35240 | 37570 | 36266 | 38059 | 37030 | 38612 | 37534 | 38866 | 40908 | 38577 | 43414 | 40520 | 44474 | 39506 | 43955 | 41203 | 44767 | 42855 | 46176 | 43474 | 46666 | 52217 |
| | No | Low | 34848 | 37616 | 36164 | 38277 | 35401 | 37898 | 36601 | 38481 | 37518 | 39135 | 37966 | 39274 | 41616 | 38705 | 43534 | 40650 | 44435 | 39431 | 44051 | 41225 | 44810 | 42794 | 46329 | 43589 | 46673 | 52443 |
| Antigenically Omicron-like, low immune escape, high virulence | Yes | High | 23512 | 24325 | 24397 | 24690 | 23491 | 24354 | 24460 | 24660 | 23916 | 24515 | 24425 | 24674 | 25180 | 37658 | 40138 | 39560 | 40957 | 37519 | 40248 | 39461 | 41013 | 39331 | 41154 | 40224 | 41474 | 43380 |
| | Yes | Low | 24348 | 25317 | 25427 | 25831 | 24373 | 25316 | 25463 | 25804 | 24911 | 25581 | 25534 | 25776 | 26429 | 37995 | 40913 | 39955 | 41660 | 38006 | 40924 | 39957 | 41625 | 40003 | 41825 | 41008 | 42278 | 44530 |
| | No | High | 24206 | 25094 | 25218 | 25567 | 24185 | 25042 | 25217 | 25574 | 24759 | 25423 | 25327 | 25539 | 26206 | 37982 | 40676 | 39827 | 41598 | 37877 | 40782 | 39724 | 41656 | 39895 | 41672 | 40776 | 42124 | 44336 |
| | No | Low | 24857 | 25956 | 25962 | 26472 | 24911 | 25944 | 25925 | 26502 | 25553 | 26263 | 26037 | 26614 | 27304 | 38174 | 41226 | 40250 | 42110 | 38116 | 41286 | 40165 | 42134 | 40323 | 42313 | 41275 | 42780 | 45146 |
| Antigenically novel, low immune escape, high virulence | Yes | High | 25794 | 26465 | 26635 | 26888 | 25845 | 26467 | 26615 | 26885 | 26185 | 26672 | 26596 | 26872 | 27227 | 42748 | 44818 | 44216 | 45411 | 43064 | 44930 | 44551 | 45553 | 44338 | 45590 | 44977 | 45761 | 47254 |
| | Yes | Low | 26908 | 27798 | 27918 | 28135 | 27048 | 27857 | 27893 | 28194 | 27519 | 28079 | 27955 | 28284 | 28720 | 43580 | 45892 | 45205 | 46613 | 43943 | 46095 | 45481 | 46735 | 45496 | 46807 | 46181 | 47175 | 48841 |
| | No | High | 26795 | 27498 | 27685 | 28001 | 26918 | 27619 | 27701 | 28030 | 27388 | 27817 | 27691 | 28059 | 28572 | 43293 | 45607 | 44994 | 46438 | 43748 | 45829 | 45343 | 46528 | 45184 | 46622 | 45928 | 46931 | 48579 |
| | No | Low | 27737 | 28662 | 28733 | 29124 | 27871 | 28743 | 28767 | 29194 | 28421 | 28904 | 28815 | 29248 | 29841 | 43936 | 46428 | 45663 | 47247 | 44465 | 46681 | 46052 | 47373 | 45981 | 47599 | 46752 | 47932 | 49685 |
| Antigenically Omicron-like, high immune escape, high virulence | Yes | High | 25949 | 26598 | 26687 | 27013 | 25928 | 26582 | 26712 | 27041 | 26288 | 26759 | 26738 | 26957 | 27366 | 43360 | 45401 | 44901 | 45954 | 43466 | 45381 | 44829 | 45938 | 44619 | 45998 | 45224 | 46218 | 47676 |
| | Yes | Low | 27197 | 27933 | 27997 | 28347 | 27191 | 27912 | 28037 | 28394 | 27632 | 28197 | 28024 | 28354 | 28896 | 44242 | 46526 | 45776 | 47252 | 44217 | 46519 | 45701 | 47208 | 45779 | 47252 | 46350 | 47673 | 49248 |
| | No | High | 26974 | 27737 | 27879 | 28130 | 26981 | 27827 | 27829 | 28146 | 27413 | 28032 | 27794 | 28176 | 28675 | 44097 | 46344 | 45551 | 47018 | 44014 | 46326 | 45563 | 46995 | 45527 | 47071 | 46218 | 47347 | 48997 |
| | No | Low | 27981 | 28805 | 28903 | 29364 | 27965 | 28835 | 28880 | 29350 | 28522 | 29154 | 28964 | 29401 | 29988 | 44758 | 47093 | 46208 | 47903 | 44655 | 47185 | 46274 | 47947 | 46333 | 47999 | 47059 | 48378 | 50363 |
| Antigenically novel, high immune escape, high virulence | Yes | High | 28060 | 28599 | 28811 | 29003 | 28100 | 28632 | 28752 | 28972 | 28445 | 28768 | 28707 | 28908 | 29332 | 47510 | 49077 | 48720 | 49593 | 47758 | 49142 | 48850 | 49595 | 48630 | 49578 | 49112 | 49751 | 50989 |
| | Yes | Low | 29543 | 30263 | 30378 | 30610 | 29643 | 30354 | 30390 | 30633 | 30008 | 30461 | 30419 | 30605 | 31137 | 48774 | 50722 | 50233 | 51258 | 49052 | 50919 | 50334 | 51370 | 50150 | 51421 | 50893 | 51695 | 53162 |
| | No | High | 29387 | 30066 | 30108 | 30406 | 29400 | 30072 | 30100 | 30463 | 29824 | 30300 | 30145 | 30429 | 30935 | 48612 | 50392 | 50026 | 50970 | 48891 | 50554 | 50078 | 51113 | 49981 | 51099 | 50456 | 51396 | 52864 |
| | No | Low | 30654 | 31429 | 31463 | 31855 | 30733 | 31482 | 31471 | 31870 | 31221 | 31779 | 31575 | 31873 | 32483 | 49705 | 51778 | 51129 | 52388 | 49933 | 51930 | 51327 | 52484 | 51230 | 52611 | 51856 | 52940 | 54505 |

*OT: Omicron-targeted vaccine in Q4 2022; 2*OT: Omicron-targeted vaccines in Q4 2022 and Q2 2023; OT+M: Omicron-targeted vaccine in Q4 2022 and multivalent vaccine in Q2 2023; 30+: administered to people aged ≥30 years; 60+ administered to people aged ≥60 years; H: high coverage; L: low coverage*

### Supplementary Figure 3: Heat map of mean COVID-19 <u>deaths</u> for each of 936 scenarios over a 12 month timeline

| | | PHSMs | Higher Stringency | | | | | | | | | | | | | Lower Stringency | | | | | | | | | | | | |
|---|---|---|---|---|---|---|---|---|---|---|---|---|---|---|---|---|---|---|---|---|---|---|---|---|---|---|---|---|
| | | Vaccine schedule | OT+M | | | | 2*OT | | | | OT | | | | Nil | OT+M | | | | 2*OT | | | | OT | | | | Nil |
| | | Vaccine age group | 30+ | | 60+ | | 30+ | | 60+ | | 30+ | | 60+ | | | 30+ | | 60+ | | 30+ | | 60+ | | 30+ | | 60+ | | |
| | | Vaccine uptake | *H* | *L* | *H* | *L* | *H* | *L* | *H* | *L* | *H* | *L* | *H* | *L* | | *H* | *L* | *H* | *L* | *H* | *L* | *H* | *L* | *H* | *L* | *H* | *L* | |
| Variant | Respirator stockpile | Mask wearing | | | | | | | | | | | | | | | | | | | | | | | | | | |
| Omicron BA.4/5 only (no new variant) | Yes | High | 2852 | 3743 | 3144 | 3921 | 2851 | 3752 | 3146 | 3928 | 3674 | 4338 | 3768 | 4402 | 5451 | 3089 | 4151 | 3461 | 4379 | 3103 | 4172 | 3460 | 4370 | 4396 | 5071 | 4298 | 5061 | 6451 |
| | Yes | Low | 2867 | 3757 | 3159 | 3922 | 2856 | 3755 | 3154 | 3932 | 3721 | 4353 | 3778 | 4411 | 5499 | 3094 | 4143 | 3467 | 4368 | 3107 | 4170 | 3463 | 4364 | 4403 | 5066 | 4303 | 5052 | 6466 |
| | No | High | 2870 | 3757 | 3149 | 3925 | 2859 | 3744 | 3154 | 3929 | 3700 | 4358 | 3769 | 4417 | 5524 | 3092 | 4158 | 3467 | 4375 | 3104 | 4185 | 3461 | 4372 | 4402 | 5073 | 4302 | 5062 | 6455 |
| | No | Low | 2874 | 3769 | 3170 | 3949 | 2873 | 3770 | 3162 | 3939 | 3734 | 4378 | 3797 | 4417 | 5539 | 3093 | 4155 | 3462 | 4373 | 3105 | 4169 | 3461 | 4366 | 4411 | 5060 | 4312 | 5066 | 6467 |
| Antigenically Omicron-like, low immune escape, low virulence | Yes | High | 3683 | 4684 | 3959 | 4812 | 3660 | 4668 | 3955 | 4792 | 4500 | 5261 | 4595 | 5296 | 6470 | 4028 | 5226 | 4313 | 5409 | 4014 | 5226 | 4313 | 5418 | 5127 | 6059 | 5125 | 6098 | 7877 |
| | Yes | Low | 3680 | 4708 | 3969 | 4856 | 3688 | 4684 | 3965 | 4830 | 4511 | 5306 | 4606 | 5341 | 6528 | 4019 | 5232 | 4304 | 5410 | 4013 | 5223 | 4301 | 5419 | 5136 | 6057 | 5126 | 6101 | 7863 |
| | No | High | 3691 | 4685 | 3969 | 4828 | 3682 | 4688 | 3968 | 4804 | 4529 | 5285 | 4613 | 5349 | 6515 | 4023 | 5216 | 4309 | 5412 | 4009 | 5230 | 4311 | 5426 | 5127 | 6052 | 5121 | 6122 | 7884 |
| | No | Low | 3681 | 4709 | 3973 | 4847 | 3677 | 4704 | 3972 | 4822 | 4537 | 5312 | 4636 | 5336 | 6586 | 4010 | 5220 | 4311 | 5424 | 4011 | 5215 | 4319 | 5422 | 5140 | 6060 | 5126 | 6104 | 7897 |
| Antigenically novel, low immune escape, low virulence | Yes | High | 4323 | 5339 | 4592 | 5462 | 4413 | 5372 | 4668 | 5487 | 5231 | 5943 | 5319 | 5992 | 7121 | 4785 | 6054 | 5085 | 6241 | 4896 | 6155 | 5188 | 6302 | 5909 | 6856 | 5902 | 6902 | 8845 |
| | Yes | Low | 4349 | 5389 | 4620 | 5529 | 4458 | 5432 | 4709 | 5541 | 5284 | 6009 | 5377 | 6067 | 7239 | 4781 | 6045 | 5085 | 6246 | 4888 | 6146 | 5186 | 6312 | 5902 | 6858 | 5905 | 6925 | 8841 |
| | No | High | 4330 | 5389 | 4638 | 5536 | 4455 | 5437 | 4707 | 5555 | 5271 | 6004 | 5369 | 6047 | 7224 | 4781 | 6040 | 5062 | 6242 | 4894 | 6149 | 5174 | 6299 | 5917 | 6852 | 5895 | 6907 | 8831 |
| | No | Low | 4361 | 5400 | 4641 | 5551 | 4468 | 5461 | 4701 | 5583 | 5296 | 6033 | 5384 | 6085 | 7272 | 4775 | 6048 | 5070 | 6252 | 4887 | 6142 | 5172 | 6291 | 5911 | 6882 | 5886 | 6905 | 8858 |
| Antigenically Omicron-like, high immune escape, low virulence | Yes | High | 4523 | 5542 | 4766 | 5672 | 4506 | 5573 | 4761 | 5674 | 5359 | 6135 | 5452 | 6182 | 7410 | 5027 | 6361 | 5320 | 6573 | 5033 | 6374 | 5310 | 6565 | 6028 | 7121 | 6070 | 7184 | 9295 |
| | Yes | Low | 4558 | 5597 | 4808 | 5740 | 4546 | 5607 | 4810 | 5745 | 5406 | 6231 | 5500 | 6270 | 7538 | 5017 | 6369 | 5319 | 6550 | 5016 | 6375 | 5325 | 6563 | 6015 | 7131 | 6074 | 7193 | 9342 |
| | No | High | 4539 | 5605 | 4819 | 5742 | 4535 | 5613 | 4821 | 5749 | 5397 | 6210 | 5503 | 6257 | 7513 | 5011 | 6352 | 5335 | 6566 | 5017 | 6369 | 5323 | 6558 | 6018 | 7137 | 6096 | 7196 | 9324 |
| | No | Low | 4558 | 5653 | 4826 | 5773 | 4552 | 5650 | 4842 | 5784 | 5427 | 6254 | 5523 | 6308 | 7593 | 5017 | 6341 | 5342 | 6566 | 5026 | 6360 | 5337 | 6547 | 6026 | 7130 | 6074 | 7190 | 9275 |
| Antigenically novel, high immune escape, low virulence | Yes | High | 5200 | 6217 | 5465 | 6316 | 5281 | 6264 | 5515 | 6368 | 6125 | 6830 | 6228 | 6882 | 8008 | 5849 | 7267 | 6187 | 7446 | 5967 | 7391 | 6281 | 7533 | 6924 | 8101 | 7079 | 8164 | 10207 |
| | Yes | Low | 5243 | 6303 | 5516 | 6411 | 5351 | 6342 | 5568 | 6461 | 6192 | 6913 | 6305 | 6999 | 8163 | 5857 | 7279 | 6167 | 7449 | 5994 | 7371 | 6277 | 7531 | 6945 | 8089 | 7025 | 8143 | 10239 |
| | No | High | 5259 | 6286 | 5512 | 6415 | 5351 | 6338 | 5555 | 6427 | 6174 | 6905 | 6283 | 6975 | 8140 | 5850 | 7285 | 6192 | 7471 | 5996 | 7376 | 6293 | 7515 | 6939 | 8083 | 7068 | 8163 | 10217 |
| | No | Low | 5284 | 6349 | 5516 | 6487 | 5383 | 6395 | 5602 | 6508 | 6247 | 7010 | 6349 | 7036 | 8277 | 5879 | 7311 | 6205 | 7460 | 5988 | 7398 | 6298 | 7518 | 6932 | 8122 | 7063 | 8160 | 10285 |
| Antigenically Omicron-like, low immune escape, high virulence | Yes | High | 6388 | 7387 | 6702 | 7514 | 6385 | 7402 | 6724 | 7503 | 7116 | 7928 | 7290 | 7959 | 9051 | 10179 | 12168 | 10822 | 12478 | 10134 | 12201 | 10788 | 12517 | 11678 | 13247 | 12005 | 13421 | 15630 |
| | Yes | Low | 6611 | 7673 | 6987 | 7868 | 6605 | 7677 | 6991 | 7859 | 7448 | 8232 | 7644 | 8341 | 9530 | 10291 | 12402 | 10900 | 12714 | 10286 | 12437 | 10912 | 12722 | 11942 | 13472 | 12243 | 13684 | 16044 |
| | No | High | 6544 | 7611 | 6919 | 7777 | 6548 | 7592 | 6925 | 7789 | 7384 | 8204 | 7589 | 8247 | 9425 | 10289 | 12338 | 10886 | 12687 | 10257 | 12377 | 10855 | 12711 | 11902 | 13440 | 12202 | 13667 | 16021 |
| | No | Low | 6724 | 7861 | 7099 | 8038 | 6745 | 7864 | 7112 | 8057 | 7626 | 8481 | 7794 | 8603 | 9840 | 10342 | 12512 | 11002 | 12849 | 10339 | 12564 | 10986 | 12875 | 12050 | 13639 | 12345 | 13881 | 16253 |
| Antigenically novel, low immune escape, high virulence | Yes | High | 7108 | 8133 | 7409 | 8317 | 7089 | 8116 | 7419 | 8287 | 7969 | 8736 | 8136 | 8802 | 9887 | 11797 | 13724 | 12262 | 13991 | 11842 | 13750 | 12371 | 14001 | 13527 | 14870 | 13705 | 14967 | 17190 |
| | Yes | Low | 7410 | 8555 | 7758 | 8690 | 7453 | 8551 | 7753 | 8685 | 8385 | 9198 | 8537 | 9276 | 10455 | 12007 | 14106 | 12558 | 14336 | 12119 | 14124 | 12629 | 14395 | 13868 | 15287 | 14081 | 15392 | 17775 |
| | No | High | 7376 | 8469 | 7700 | 8662 | 7413 | 8473 | 7704 | 8644 | 8340 | 9099 | 8465 | 9245 | 10351 | 11933 | 14019 | 12494 | 14305 | 12037 | 14068 | 12582 | 14321 | 13759 | 15262 | 14027 | 15368 | 17630 |
| | No | Low | 7648 | 8815 | 8027 | 8993 | 7658 | 8820 | 8010 | 9005 | 8677 | 9440 | 8819 | 9591 | 10855 | 12094 | 14264 | 12658 | 14557 | 12234 | 14307 | 12806 | 14592 | 14025 | 15600 | 14265 | 15681 | 18078 |
| Antigenically Omicron-like, high immune escape, high virulence | Yes | High | 7243 | 8306 | 7552 | 8487 | 7230 | 8314 | 7562 | 8499 | 8144 | 8939 | 8304 | 9035 | 10214 | 12134 | 14170 | 12682 | 14414 | 12169 | 14171 | 12616 | 14428 | 13813 | 15290 | 13996 | 15396 | 17769 |
| | Yes | Low | 7577 | 8775 | 7882 | 8911 | 7574 | 8749 | 7903 | 8930 | 8525 | 9432 | 8682 | 9480 | 10779 | 12350 | 14548 | 12931 | 14827 | 12338 | 14575 | 12877 | 14849 | 14127 | 15764 | 14345 | 15944 | 18384 |
| | No | High | 7526 | 8668 | 7878 | 8837 | 7537 | 8730 | 7858 | 8868 | 8463 | 9360 | 8625 | 9449 | 10690 | 12317 | 14509 | 12814 | 14748 | 12301 | 14485 | 12811 | 14767 | 14085 | 15693 | 14293 | 15834 | 18283 |
| | No | Low | 7793 | 9031 | 8168 | 9264 | 7779 | 9027 | 8155 | 9231 | 8817 | 9775 | 9001 | 9861 | 11201 | 12500 | 14780 | 13003 | 15059 | 12446 | 14800 | 13039 | 15107 | 14317 | 16018 | 14583 | 16201 | 18806 |
| Antigenically novel, high immune escape, high virulence | Yes | High | 8026 | 9141 | 8319 | 9268 | 8033 | 9123 | 8331 | 9243 | 9063 | 9782 | 9165 | 9845 | 11066 | 13550 | 15665 | 14039 | 15868 | 13675 | 15618 | 14114 | 15856 | 15440 | 16839 | 15604 | 16923 | 19184 |
| | Yes | Low | 8452 | 9617 | 8761 | 9781 | 8474 | 9642 | 8804 | 9808 | 9537 | 10365 | 9715 | 10428 | 11684 | 13939 | 16089 | 14473 | 16348 | 14026 | 16154 | 14541 | 16417 | 15907 | 17407 | 16180 | 17574 | 20021 |
| | No | High | 8407 | 9572 | 8670 | 9728 | 8414 | 9579 | 8679 | 9754 | 9514 | 10308 | 9612 | 10411 | 11676 | 13859 | 16061 | 14405 | 16309 | 13955 | 16093 | 14494 | 16350 | 15828 | 17354 | 16063 | 17499 | 19964 |
| | No | Low | 8773 | 10029 | 9088 | 10197 | 8809 | 10046 | 9116 | 10196 | 9945 | 10860 | 10094 | 10860 | 12265 | 14209 | 16458 | 14678 | 16757 | 14285 | 16520 | 14807 | 16818 | 16239 | 17840 | 16489 | 18060 | 20590 |

*OT: Omicron-targeted vaccine in Q4 2022; 2*OT: Omicron-targeted vaccines in Q4 2022 and Q2 2023; OT+M: Omicron-targeted vaccine in Q4 2022 and multivalent vaccine in Q2 2023; 30+: administered to people aged ≥30 years; 60+ administered to people aged ≥60 years; H: high coverage; L: low coverage*

### Supplementary Figure 4: Heat map of mean time in stage 3 and above (days) for each of 936 scenarios over a 12 month timeline

| | | PHSMs | Higher Stringency | | | | | | | | | | | | | Lower Stringency | | | | | | | | | | | | |
|---|---|---|---|---|---|---|---|---|---|---|---|---|---|---|---|---|---|---|---|---|---|---|---|---|---|---|---|---|
| | | Vaccine schedule | OT+M | | | | 2*OT | | | | OT | | | | Nil | OT+M | | | | 2*OT | | | | OT | | | | Nil |
| | | Vaccine age group | 30+ | | 60+ | | 30+ | | 60+ | | 30+ | | 60+ | | | 30+ | | 60+ | | 30+ | | 60+ | | 30+ | | 60+ | | |
| | | Vaccine uptake | *H* | *L* | *H* | *L* | *H* | *L* | *H* | *L* | *H* | *L* | *H* | *L* | | *H* | *L* | *H* | *L* | *H* | *L* | *H* | *L* | *H* | *L* | *H* | *L* | |
| **Variant** | **Respirator stockpile** | **Mask wearing** | | | | | | | | | | | | | | | | | | | | | | | | | | |
| **Omicron BA.4/5 only (no new variant)** | Yes | High | 6 | 8 | 6 | 8 | 6 | 7 | 6 | 8 | 8 | 9 | 8 | 10 | 13 | 1 | 1 | 1 | 1 | 1 | 1 | 1 | 1 | 1 | 1 | 1 | 1 | 2 |
| | Yes | Low | 6 | 8 | 7 | 8 | 6 | 8 | 6 | 8 | 8 | 9 | 7 | 9 | 13 | 1 | 1 | 1 | 1 | 1 | 1 | 1 | 1 | 1 | 1 | 1 | 1 | 2 |
| | No | High | 6 | 8 | 6 | 8 | 6 | 8 | 7 | 8 | 8 | 9 | 8 | 9 | 13 | 1 | 1 | 1 | 1 | 1 | 1 | 1 | 1 | 1 | 1 | 1 | 1 | 2 |
| | No | Low | 6 | 7 | 7 | 8 | 6 | 7 | 7 | 8 | 8 | 9 | 8 | 10 | 13 | 1 | 1 | 1 | 1 | 1 | 1 | 1 | 1 | 1 | 1 | 1 | 1 | 2 |
| **Antigenically Omicron-like, low immune escape, low virulence** | Yes | High | 10 | 14 | 11 | 15 | 10 | 14 | 11 | 15 | 12 | 16 | 13 | 17 | 25 | 1 | 1 | 1 | 1 | 1 | 1 | 1 | 1 | 1 | 1 | 1 | 2 | 3 |
| | Yes | Low | 9 | 13 | 11 | 15 | 10 | 14 | 11 | 15 | 12 | 15 | 13 | 17 | 24 | 1 | 1 | 1 | 1 | 1 | 1 | 1 | 1 | 1 | 1 | 1 | 1 | 3 |
| | No | High | 9 | 15 | 12 | 15 | 9 | 14 | 12 | 15 | 12 | 17 | 13 | 17 | 24 | 1 | 1 | 1 | 1 | 1 | 1 | 1 | 1 | 1 | 1 | 1 | 1 | 3 |
| | No | Low | 9 | 14 | 11 | 15 | 9 | 14 | 11 | 15 | 11 | 16 | 13 | 16 | 24 | 1 | 1 | 1 | 1 | 1 | 1 | 1 | 1 | 1 | 1 | 1 | 2 | 3 |
| **Antigenically novel, low immune escape, low virulence** | Yes | High | 19 | 25 | 22 | 27 | 20 | 26 | 23 | 28 | 23 | 29 | 25 | 30 | 40 | 2 | 4 | 3 | 4 | 2 | 4 | 3 | 4 | 2 | 4 | 3 | 4 | 6 |
| | Yes | Low | 19 | 26 | 22 | 27 | 19 | 26 | 22 | 27 | 23 | 29 | 25 | 30 | 40 | 2 | 4 | 3 | 4 | 2 | 4 | 3 | 4 | 2 | 4 | 3 | 4 | 6 |
| | No | High | 20 | 26 | 22 | 28 | 20 | 26 | 23 | 28 | 23 | 29 | 25 | 30 | 41 | 2 | 4 | 3 | 4 | 2 | 4 | 3 | 4 | 2 | 4 | 3 | 4 | 6 |
| | No | Low | 20 | 26 | 23 | 28 | 20 | 27 | 23 | 28 | 23 | 29 | 26 | 31 | 41 | 2 | 4 | 3 | 4 | 2 | 4 | 3 | 4 | 2 | 4 | 3 | 4 | 6 |
| **Antigenically Omicron-like, high immune escape, low virulence** | Yes | High | 20 | 27 | 22 | 29 | 19 | 27 | 22 | 29 | 23 | 29 | 25 | 31 | 41 | 2 | 4 | 3 | 4 | 2 | 4 | 3 | 4 | 3 | 4 | 3 | 4 | 7 |
| | Yes | Low | 19 | 26 | 22 | 29 | 20 | 27 | 22 | 28 | 23 | 29 | 24 | 31 | 42 | 2 | 4 | 3 | 4 | 2 | 4 | 3 | 4 | 2 | 4 | 3 | 4 | 7 |
| | No | High | 20 | 28 | 23 | 29 | 20 | 28 | 23 | 29 | 23 | 30 | 26 | 31 | 43 | 2 | 4 | 3 | 4 | 2 | 4 | 3 | 4 | 2 | 4 | 3 | 4 | 7 |
| | No | Low | 21 | 27 | 23 | 29 | 21 | 27 | 23 | 29 | 24 | 30 | 25 | 31 | 42 | 3 | 4 | 3 | 4 | 3 | 4 | 3 | 4 | 3 | 4 | 3 | 4 | 7 |
| **Antigenically novel, high immune escape, low virulence** | Yes | High | 34 | 44 | 38 | 45 | 35 | 44 | 38 | 46 | 40 | 48 | 42 | 49 | 60 | 7 | 9 | 8 | 10 | 7 | 9 | 8 | 10 | 7 | 9 | 8 | 10 | 14 |
| | Yes | Low | 35 | 45 | 39 | 45 | 36 | 45 | 40 | 46 | 40 | 48 | 43 | 49 | 61 | 7 | 9 | 8 | 10 | 7 | 9 | 8 | 10 | 7 | 9 | 8 | 10 | 14 |
| | No | High | 36 | 45 | 39 | 47 | 37 | 45 | 40 | 47 | 41 | 48 | 43 | 50 | 62 | 7 | 9 | 8 | 10 | 7 | 9 | 8 | 10 | 7 | 9 | 8 | 10 | 14 |
| | No | Low | 36 | 46 | 40 | 46 | 37 | 48 | 41 | 46 | 42 | 51 | 44 | 50 | 63 | 7 | 10 | 8 | 11 | 7 | 10 | 8 | 11 | 7 | 10 | 8 | 11 | 15 |
| **Antigenically Omicron-like, low immune escape, high virulence** | Yes | High | 67 | 82 | 75 | 86 | 67 | 82 | 75 | 87 | 80 | 90 | 84 | 91 | 103 | 17 | 27 | 20 | 29 | 16 | 27 | 20 | 29 | 26 | 33 | 27 | 34 | 48 |
| | Yes | Low | 70 | 87 | 79 | 93 | 70 | 88 | 79 | 93 | 86 | 96 | 89 | 100 | 113 | 16 | 26 | 19 | 29 | 16 | 26 | 20 | 29 | 26 | 33 | 26 | 35 | 49 |
| | No | High | 71 | 88 | 79 | 93 | 70 | 87 | 79 | 92 | 86 | 96 | 90 | 99 | 112 | 16 | 27 | 21 | 30 | 16 | 27 | 21 | 30 | 26 | 35 | 28 | 36 | 49 |
| | No | Low | 74 | 93 | 84 | 97 | 75 | 93 | 85 | 97 | 92 | 104 | 96 | 106 | 122 | 17 | 27 | 20 | 30 | 17 | 27 | 21 | 30 | 27 | 35 | 28 | 36 | 51 |
| **Antigenically novel, low immune escape, high virulence** | Yes | High | 103 | 117 | 111 | 120 | 106 | 119 | 113 | 122 | 116 | 124 | 120 | 125 | 134 | 38 | 52 | 44 | 56 | 41 | 53 | 45 | 57 | 51 | 61 | 54 | 62 | 77 |
| | Yes | Low | 110 | 126 | 120 | 130 | 115 | 128 | 123 | 131 | 126 | 135 | 130 | 138 | 149 | 39 | 54 | 44 | 57 | 41 | 55 | 45 | 58 | 52 | 64 | 55 | 64 | 79 |
| | No | High | 109 | 126 | 118 | 131 | 113 | 128 | 120 | 132 | 125 | 134 | 129 | 136 | 147 | 40 | 54 | 44 | 57 | 42 | 55 | 46 | 58 | 53 | 63 | 55 | 64 | 81 |
| | No | Low | 119 | 135 | 129 | 139 | 122 | 139 | 131 | 142 | 135 | 145 | 139 | 147 | 159 | 40 | 55 | 45 | 58 | 42 | 57 | 47 | 60 | 53 | 66 | 56 | 67 | 86 |
| **Antigenically Omicron-like, high immune escape, high virulence** | Yes | High | 107 | 120 | 114 | 124 | 107 | 119 | 115 | 124 | 117 | 126 | 121 | 128 | 137 | 42 | 55 | 47 | 58 | 42 | 55 | 47 | 58 | 51 | 62 | 54 | 64 | 80 |
| | Yes | Low | 116 | 130 | 124 | 134 | 115 | 131 | 124 | 135 | 128 | 138 | 132 | 139 | 152 | 41 | 56 | 48 | 60 | 42 | 57 | 47 | 59 | 52 | 64 | 56 | 67 | 84 |
| | No | High | 116 | 130 | 124 | 132 | 116 | 130 | 124 | 133 | 127 | 137 | 132 | 138 | 149 | 43 | 57 | 48 | 60 | 42 | 56 | 48 | 61 | 54 | 64 | 56 | 67 | 85 |
| | No | Low | 124 | 141 | 133 | 145 | 124 | 140 | 133 | 146 | 138 | 148 | 141 | 150 | 163 | 43 | 59 | 48 | 63 | 42 | 58 | 49 | 62 | 55 | 67 | 58 | 69 | 89 |
| **Antigenically novel, high immune escape, high virulence** | Yes | High | 139 | 149 | 145 | 151 | 142 | 150 | 147 | 152 | 149 | 154 | 151 | 155 | 162 | 72 | 86 | 78 | 90 | 75 | 88 | 80 | 91 | 85 | 94 | 88 | 97 | 108 |
| | Yes | Low | 153 | 166 | 158 | 169 | 156 | 167 | 162 | 171 | 165 | 173 | 168 | 172 | 180 | 75 | 89 | 80 | 94 | 78 | 91 | 83 | 95 | 89 | 99 | 91 | 101 | 116 |
| | No | High | 151 | 163 | 158 | 165 | 154 | 164 | 160 | 167 | 162 | 168 | 164 | 170 | 177 | 75 | 90 | 81 | 95 | 78 | 93 | 84 | 96 | 90 | 99 | 93 | 102 | 117 |
| | No | Low | 165 | 178 | 172 | 181 | 169 | 179 | 174 | 182 | 177 | 185 | 180 | 185 | 194 | 77 | 94 | 84 | 99 | 81 | 96 | 87 | 101 | 93 | 104 | 97 | 106 | 124 |

*OT: Omicron-targeted vaccine in Q4 2022; 2*OT: Omicron-targeted vaccines in Q4 2022 and Q2 2023; OT+M: Omicron-targeted vaccine in Q4 2022 and multivalent vaccine in Q2 2023; 30+: administered to people aged ≥30 years; 60+ administered to people aged ≥60 years; H: high coverage; L: low coverage*

### Supplementary Figure 5: Heat map of mean <u>number of days with >750 COVID-19 patients in hospital</u> for each of 936 scenarios over a 12 month timeline

| | | PHSMs | Higher Stringency | | | | | | | | | | | | | Lower Stringency | | | | | | | | | | | | |
|---|---|---|---|---|---|---|---|---|---|---|---|---|---|---|---|---|---|---|---|---|---|---|---|---|---|---|---|---|
| | | Vaccine schedule | OT+M | | | | 2*OT | | | | OT | | | | Nil | OT+M | | | | 2*OT | | | | OT | | | | Nil |
| | | Vaccine age group | 30+ | | 60+ | | 30+ | | 60+ | | 30+ | | 60+ | | | 30+ | | 60+ | | 30+ | | 60+ | | 30+ | | 60+ | | |
| | | Vaccine uptake | *H* | *L* | *H* | *L* | *H* | *L* | *H* | *L* | *H* | *L* | *H* | *L* | | *H* | *L* | *H* | *L* | *H* | *L* | *H* | *L* | *H* | *L* | *H* | *L* | |
| Variant | Respirator stockpile | Mask wearing | | | | | | | | | | | | | | | | | | | | | | | | | | |
| Omicron BA.4/5 only (no new variant) | Yes | High | 11 | 15 | 13 | 16 | 11 | 14 | 13 | 16 | 15 | 17 | 15 | 18 | 24 | 28 | 40 | 34 | 44 | 28 | 40 | 34 | 43 | 50 | 54 | 47 | 52 | 69 |
| | Yes | Low | 11 | 15 | 12 | 15 | 11 | 15 | 13 | 16 | 15 | 17 | 15 | 18 | 24 | 28 | 40 | 34 | 44 | 28 | 40 | 34 | 43 | 51 | 54 | 47 | 53 | 69 |
| | No | High | 11 | 15 | 13 | 16 | 11 | 15 | 13 | 16 | 15 | 18 | 15 | 18 | 24 | 28 | 39 | 34 | 44 | 28 | 40 | 34 | 43 | 51 | 54 | 47 | 53 | 69 |
| | No | Low | 11 | 14 | 13 | 16 | 11 | 15 | 13 | 16 | 15 | 17 | 15 | 18 | 24 | 29 | 40 | 34 | 44 | 29 | 40 | 34 | 43 | 51 | 54 | 47 | 53 | 69 |
| Antigenically Omicron-like, low immune escape, low virulence | Yes | High | 19 | 24 | 21 | 26 | 18 | 25 | 21 | 26 | 23 | 28 | 24 | 29 | 39 | 46 | 62 | 52 | 65 | 45 | 62 | 52 | 65 | 59 | 72 | 60 | 73 | 99 |
| | Yes | Low | 18 | 24 | 21 | 26 | 18 | 24 | 21 | 25 | 22 | 27 | 24 | 28 | 38 | 46 | 62 | 51 | 64 | 46 | 62 | 52 | 65 | 60 | 72 | 60 | 73 | 98 |
| | No | High | 18 | 25 | 21 | 26 | 18 | 25 | 21 | 26 | 22 | 28 | 24 | 29 | 39 | 46 | 61 | 52 | 64 | 46 | 61 | 52 | 65 | 59 | 72 | 60 | 73 | 99 |
| | No | Low | 18 | 24 | 20 | 26 | 18 | 24 | 20 | 26 | 22 | 28 | 24 | 28 | 39 | 45 | 61 | 52 | 65 | 45 | 61 | 53 | 65 | 59 | 72 | 60 | 73 | 99 |
| Antigenically novel, low immune escape, low virulence | Yes | High | 30 | 38 | 34 | 40 | 31 | 39 | 34 | 41 | 36 | 42 | 38 | 43 | 55 | 71 | 86 | 76 | 90 | 72 | 88 | 78 | 91 | 82 | 95 | 84 | 97 | 123 |
| | Yes | Low | 30 | 39 | 34 | 40 | 31 | 39 | 34 | 41 | 36 | 43 | 38 | 44 | 55 | 71 | 86 | 77 | 90 | 72 | 87 | 78 | 91 | 82 | 94 | 84 | 97 | 124 |
| | No | High | 30 | 39 | 34 | 41 | 31 | 40 | 34 | 41 | 36 | 43 | 38 | 44 | 57 | 71 | 86 | 76 | 90 | 72 | 87 | 78 | 90 | 82 | 95 | 84 | 97 | 124 |
| | No | Low | 31 | 39 | 34 | 41 | 31 | 39 | 35 | 41 | 36 | 43 | 38 | 45 | 57 | 71 | 87 | 77 | 90 | 72 | 88 | 78 | 91 | 83 | 96 | 84 | 97 | 124 |
| Antigenically Omicron-like, high immune escape, low virulence | Yes | High | 32 | 40 | 35 | 42 | 32 | 40 | 35 | 42 | 37 | 43 | 39 | 45 | 57 | 74 | 90 | 78 | 93 | 73 | 90 | 77 | 92 | 84 | 99 | 85 | 99 | 126 |
| | Yes | Low | 31 | 40 | 34 | 42 | 31 | 41 | 35 | 42 | 36 | 44 | 38 | 45 | 57 | 74 | 90 | 77 | 93 | 73 | 90 | 78 | 93 | 83 | 99 | 85 | 100 | 128 |
| | No | High | 31 | 41 | 35 | 43 | 31 | 41 | 35 | 43 | 37 | 44 | 39 | 46 | 58 | 73 | 89 | 78 | 93 | 73 | 90 | 78 | 93 | 83 | 98 | 85 | 99 | 127 |
| | No | Low | 32 | 41 | 36 | 42 | 32 | 41 | 35 | 43 | 36 | 45 | 39 | 46 | 58 | 73 | 90 | 78 | 92 | 74 | 90 | 78 | 92 | 84 | 99 | 85 | 99 | 127 |
| Antigenically novel, high immune escape, low virulence | Yes | High | 48 | 56 | 51 | 59 | 49 | 57 | 51 | 59 | 54 | 62 | 55 | 62 | 73 | 96 | 111 | 102 | 114 | 98 | 113 | 103 | 115 | 107 | 120 | 110 | 121 | 146 |
| | Yes | Low | 47 | 58 | 52 | 59 | 49 | 58 | 52 | 60 | 54 | 62 | 57 | 64 | 75 | 97 | 111 | 101 | 115 | 98 | 113 | 103 | 117 | 107 | 120 | 109 | 122 | 147 |
| | No | High | 48 | 58 | 52 | 60 | 49 | 59 | 52 | 60 | 54 | 63 | 57 | 63 | 75 | 96 | 112 | 102 | 115 | 97 | 113 | 103 | 116 | 106 | 120 | 110 | 122 | 147 |
| | No | Low | 49 | 59 | 52 | 61 | 50 | 60 | 53 | 61 | 56 | 65 | 57 | 65 | 77 | 96 | 112 | 102 | 115 | 98 | 114 | 103 | 116 | 106 | 120 | 110 | 122 | 148 |
| Antigenically Omicron-like, low immune escape, high virulence | Yes | High | 83 | 97 | 90 | 99 | 83 | 97 | 90 | 100 | 94 | 103 | 97 | 104 | 113 | 215 | 233 | 226 | 236 | 214 | 235 | 224 | 237 | 229 | 241 | 233 | 241 | 255 |
| | Yes | Low | 87 | 101 | 95 | 107 | 86 | 102 | 96 | 107 | 100 | 109 | 104 | 112 | 122 | 220 | 243 | 230 | 245 | 219 | 243 | 230 | 245 | 236 | 249 | 242 | 251 | 266 |
| | No | High | 86 | 101 | 95 | 106 | 86 | 101 | 94 | 106 | 99 | 109 | 102 | 111 | 121 | 220 | 239 | 229 | 244 | 217 | 240 | 226 | 244 | 234 | 247 | 239 | 248 | 263 |
| | No | Low | 90 | 107 | 99 | 110 | 90 | 107 | 100 | 111 | 105 | 115 | 108 | 118 | 131 | 221 | 246 | 234 | 249 | 221 | 247 | 232 | 249 | 239 | 254 | 245 | 255 | 271 |
| Antigenically novel, low immune escape, high virulence | Yes | High | 114 | 124 | 120 | 128 | 116 | 126 | 122 | 130 | 124 | 130 | 127 | 132 | 139 | 248 | 259 | 251 | 259 | 249 | 259 | 256 | 261 | 259 | 261 | 260 | 263 | 267 |
| | Yes | Low | 120 | 134 | 130 | 137 | 125 | 135 | 132 | 138 | 134 | 141 | 137 | 142 | 151 | 257 | 268 | 264 | 272 | 260 | 271 | 267 | 272 | 269 | 273 | 272 | 277 | 280 |
| | No | High | 120 | 134 | 128 | 137 | 122 | 136 | 130 | 138 | 133 | 140 | 136 | 142 | 149 | 254 | 265 | 260 | 268 | 257 | 268 | 264 | 270 | 266 | 271 | 269 | 272 | 278 |
| | No | Low | 129 | 142 | 137 | 145 | 131 | 145 | 139 | 147 | 141 | 149 | 145 | 151 | 161 | 259 | 274 | 267 | 278 | 264 | 276 | 271 | 279 | 274 | 279 | 277 | 282 | 286 |
| Antigenically Omicron-like, high immune escape, high virulence | Yes | High | 117 | 127 | 123 | 130 | 118 | 127 | 124 | 130 | 125 | 131 | 128 | 133 | 141 | 252 | 261 | 257 | 262 | 255 | 262 | 257 | 262 | 260 | 264 | 260 | 263 | 267 |
| | Yes | Low | 126 | 137 | 132 | 141 | 126 | 137 | 133 | 141 | 135 | 143 | 139 | 144 | 154 | 262 | 275 | 269 | 277 | 262 | 274 | 267 | 276 | 271 | 278 | 272 | 278 | 282 |
| | No | High | 125 | 137 | 132 | 138 | 125 | 137 | 132 | 139 | 134 | 142 | 138 | 143 | 151 | 259 | 270 | 266 | 272 | 259 | 271 | 266 | 272 | 268 | 275 | 270 | 274 | 278 |
| | No | Low | 134 | 146 | 139 | 149 | 132 | 146 | 140 | 150 | 142 | 152 | 147 | 153 | 163 | 267 | 278 | 272 | 281 | 267 | 279 | 272 | 281 | 276 | 283 | 278 | 284 | 290 |
| Antigenically novel, high immune escape, high virulence | Yes | High | 142 | 149 | 146 | 151 | 144 | 150 | 147 | 152 | 149 | 152 | 151 | 153 | 159 | 265 | 267 | 266 | 268 | 264 | 270 | 266 | 268 | 268 | 270 | 268 | 269 | 274 |
| | Yes | Low | 155 | 165 | 159 | 165 | 157 | 166 | 162 | 168 | 163 | 169 | 165 | 169 | 176 | 278 | 285 | 281 | 283 | 279 | 285 | 283 | 286 | 283 | 286 | 286 | 286 | 290 |
| | No | High | 153 | 162 | 158 | 163 | 155 | 162 | 159 | 165 | 160 | 165 | 162 | 166 | 173 | 274 | 278 | 278 | 279 | 275 | 280 | 278 | 280 | 278 | 281 | 279 | 281 | 285 |
| | No | Low | 165 | 175 | 171 | 177 | 167 | 176 | 172 | 178 | 174 | 180 | 177 | 180 | 186 | 285 | 290 | 287 | 290 | 285 | 291 | 288 | 291 | 290 | 292 | 290 | 292 | 295 |

*OT: Omicron-targeted vaccine in Q4 2022; 2*OT: Omicron-targeted vaccines in Q4 2022 and Q2 2023; OT+M: Omicron-targeted vaccine in Q4 2022 and multivalent vaccine in Q2 2023; 30+: administered to people aged ≥30 years; 60+ administered to people aged ≥60 years; H: high coverage; L: low coverage.*

### Supplementary Figure 6: Heat map of mean days with >1,500 COVID-19 patients in hospital for each of 936 scenarios over a 12 month timeline

| | | PHSMs | Higher Stringency | | | | | | | | | | | | | Lower Stringency | | | | | | | | | | | | |
|---|---|---|---|---|---|---|---|---|---|---|---|---|---|---|---|---|---|---|---|---|---|---|---|---|---|---|---|---|
| | | Vaccine schedule | OT+M | | | | 2*OT | | | | OT | | | | Nil | OT+M | | | | 2*OT | | | | OT | | | | Nil |
| | | Vaccine age group | 30+ | | 60+ | | 30+ | | 60+ | | 30+ | | 60+ | | | 30+ | | 60+ | | 30+ | | 60+ | | 30+ | | 60+ | | |
| | | Vaccine uptake | *H* | *L* | *H* | *L* | *H* | *L* | *H* | *L* | *H* | *L* | *H* | *L* | | *H* | *L* | *H* | *L* | *H* | *L* | *H* | *L* | *H* | *L* | *H* | *L* | |
| **Variant** | **Respirator stockpile** | **Mask wearing** | | | | | | | | | | | | | | | | | | | | | | | | | | |
| **Omicron BA.4/5 only (no new variant)** | Yes | High | 1 | 1 | 1 | 1 | 1 | 1 | 1 | 1 | 1 | 1 | 1 | 1 | 1 | 2 | 3 | 2 | 3 | 2 | 3 | 2 | 3 | 3 | 4 | 3 | 3 | 4 |
| | Yes | Low | 1 | 1 | 1 | 1 | 1 | 1 | 1 | 1 | 1 | 1 | 1 | 1 | 1 | 2 | 3 | 2 | 3 | 2 | 3 | 2 | 3 | 3 | 4 | 3 | 3 | 4 |
| | No | High | 1 | 1 | 1 | 1 | 1 | 1 | 1 | 1 | 1 | 1 | 1 | 1 | 1 | 2 | 3 | 2 | 3 | 2 | 3 | 2 | 3 | 3 | 4 | 3 | 3 | 4 |
| | No | Low | 1 | 1 | 1 | 1 | 1 | 1 | 1 | 1 | 1 | 1 | 1 | 1 | 1 | 2 | 3 | 2 | 3 | 2 | 3 | 2 | 3 | 3 | 3 | 3 | 3 | 4 |
| **Antigenically Omicron-like, low immune escape, low virulence** | Yes | High | 1 | 1 | 1 | 1 | 1 | 1 | 1 | 1 | 1 | 1 | 1 | 1 | 1 | 3 | 4 | 3 | 4 | 3 | 4 | 3 | 4 | 4 | 5 | 4 | 5 | 7 |
| | Yes | Low | 1 | 1 | 1 | 1 | 1 | 1 | 1 | 1 | 1 | 1 | 1 | 1 | 1 | 3 | 4 | 4 | 4 | 3 | 4 | 4 | 4 | 4 | 4 | 4 | 5 | 7 |
| | No | High | 1 | 1 | 1 | 1 | 1 | 1 | 1 | 1 | 1 | 1 | 1 | 1 | 1 | 3 | 4 | 4 | 4 | 3 | 4 | 4 | 4 | 4 | 4 | 4 | 5 | 7 |
| | No | Low | 1 | 1 | 1 | 1 | 1 | 1 | 1 | 1 | 1 | 1 | 1 | 1 | 1 | 3 | 4 | 4 | 4 | 3 | 4 | 4 | 4 | 4 | 5 | 4 | 5 | 7 |
| **Antigenically novel, low immune escape, low virulence** | Yes | High | 1 | 1 | 1 | 1 | 1 | 1 | 1 | 1 | 1 | 1 | 1 | 1 | 1 | 7 | 9 | 7 | 10 | 7 | 9 | 7 | 9 | 7 | 9 | 7 | 10 | 14 |
| | Yes | Low | 1 | 1 | 1 | 1 | 1 | 1 | 1 | 1 | 1 | 1 | 1 | 1 | 1 | 7 | 9 | 7 | 10 | 7 | 9 | 7 | 9 | 7 | 9 | 7 | 10 | 14 |
| | No | High | 1 | 1 | 1 | 1 | 1 | 1 | 1 | 1 | 1 | 1 | 1 | 1 | 1 | 6 | 9 | 7 | 10 | 6 | 9 | 7 | 10 | 7 | 9 | 7 | 10 | 14 |
| | No | Low | 1 | 1 | 1 | 1 | 1 | 1 | 1 | 1 | 1 | 1 | 1 | 1 | 1 | 7 | 9 | 7 | 10 | 7 | 9 | 7 | 10 | 7 | 9 | 7 | 10 | 14 |
| **Antigenically Omicron-like, high immune escape, low virulence** | Yes | High | 1 | 1 | 1 | 1 | 1 | 1 | 1 | 1 | 1 | 1 | 1 | 1 | 1 | 7 | 9 | 8 | 10 | 7 | 9 | 8 | 10 | 7 | 10 | 8 | 10 | 15 |
| | Yes | Low | 1 | 1 | 1 | 1 | 1 | 1 | 1 | 1 | 1 | 1 | 1 | 1 | 1 | 7 | 9 | 8 | 10 | 7 | 9 | 8 | 10 | 7 | 10 | 8 | 10 | 15 |
| | No | High | 1 | 1 | 1 | 1 | 1 | 1 | 1 | 1 | 1 | 1 | 1 | 1 | 1 | 7 | 9 | 8 | 10 | 7 | 9 | 8 | 10 | 7 | 9 | 8 | 10 | 15 |
| | No | Low | 1 | 1 | 1 | 1 | 1 | 1 | 1 | 1 | 1 | 1 | 1 | 1 | 1 | 7 | 9 | 8 | 10 | 7 | 9 | 8 | 10 | 7 | 9 | 8 | 10 | 15 |
| **Antigenically novel, high immune escape, low virulence** | Yes | High | 1 | 2 | 1 | 2 | 1 | 2 | 2 | 2 | 1 | 2 | 1 | 2 | 2 | 14 | 17 | 15 | 18 | 14 | 17 | 15 | 18 | 14 | 17 | 15 | 18 | 24 |
| | Yes | Low | 1 | 2 | 1 | 2 | 1 | 2 | 1 | 2 | 1 | 2 | 2 | 2 | 2 | 14 | 17 | 15 | 18 | 14 | 17 | 15 | 18 | 14 | 17 | 15 | 18 | 24 |
| | No | High | 1 | 2 | 1 | 2 | 1 | 2 | 1 | 2 | 1 | 2 | 1 | 2 | 2 | 14 | 17 | 15 | 18 | 14 | 17 | 15 | 18 | 14 | 17 | 15 | 18 | 24 |
| | No | Low | 1 | 2 | 2 | 2 | 1 | 2 | 2 | 2 | 2 | 2 | 2 | 2 | 2 | 13 | 17 | 15 | 18 | 13 | 17 | 15 | 18 | 14 | 18 | 15 | 19 | 24 |
| **Antigenically Omicron-like, low immune escape, high virulence** | Yes | High | 2 | 2 | 2 | 2 | 2 | 2 | 2 | 2 | 2 | 2 | 2 | 2 | 3 | 33 | 45 | 38 | 50 | 33 | 46 | 38 | 49 | 45 | 53 | 46 | 55 | 69 |
| | Yes | Low | 1 | 2 | 2 | 2 | 2 | 2 | 2 | 2 | 2 | 3 | 2 | 3 | 4 | 32 | 45 | 37 | 49 | 32 | 46 | 38 | 50 | 44 | 53 | 46 | 55 | 71 |
| | No | High | 2 | 2 | 2 | 2 | 2 | 2 | 2 | 2 | 2 | 3 | 2 | 3 | 4 | 33 | 46 | 37 | 50 | 33 | 46 | 38 | 50 | 45 | 54 | 47 | 56 | 71 |
| | No | Low | 2 | 3 | 2 | 3 | 2 | 3 | 2 | 3 | 3 | 3 | 3 | 3 | 4 | 33 | 46 | 38 | 50 | 33 | 46 | 38 | 51 | 46 | 54 | 46 | 57 | 73 |
| **Antigenically novel, low immune escape, high virulence** | Yes | High | 3 | 4 | 4 | 4 | 3 | 4 | 4 | 4 | 4 | 5 | 5 | 5 | 7 | 60 | 74 | 65 | 78 | 63 | 76 | 67 | 78 | 72 | 83 | 76 | 82 | 96 |
| | Yes | Low | 4 | 4 | 4 | 5 | 4 | 5 | 4 | 5 | 5 | 6 | 5 | 6 | 8 | 60 | 77 | 66 | 79 | 63 | 78 | 68 | 80 | 75 | 86 | 77 | 86 | 100 |
| | No | High | 3 | 5 | 4 | 5 | 4 | 5 | 4 | 6 | 5 | 6 | 5 | 6 | 8 | 60 | 75 | 66 | 79 | 63 | 77 | 68 | 80 | 75 | 84 | 78 | 86 | 101 |
| | No | Low | 4 | 6 | 5 | 6 | 4 | 6 | 5 | 7 | 6 | 7 | 6 | 8 | 10 | 61 | 77 | 67 | 81 | 64 | 79 | 70 | 81 | 76 | 88 | 78 | 88 | 105 |
| **Antigenically Omicron-like, high immune escape, high virulence** | Yes | High | 3 | 4 | 4 | 5 | 4 | 5 | 3 | 5 | 4 | 5 | 4 | 6 | 6 | 64 | 78 | 69 | 80 | 63 | 77 | 68 | 79 | 74 | 84 | 76 | 86 | 100 |
| | Yes | Low | 4 | 5 | 4 | 5 | 4 | 5 | 4 | 5 | 5 | 6 | 5 | 6 | 8 | 64 | 79 | 69 | 83 | 64 | 80 | 69 | 82 | 76 | 87 | 78 | 90 | 104 |
| | No | High | 4 | 5 | 4 | 6 | 4 | 5 | 4 | 6 | 5 | 6 | 5 | 8 | 9 | 65 | 80 | 69 | 81 | 64 | 79 | 70 | 83 | 76 | 86 | 78 | 88 | 105 |
| | No | Low | 4 | 6 | 5 | 7 | 4 | 6 | 5 | 7 | 6 | 7 | 6 | 8 | 10 | 66 | 82 | 70 | 85 | 64 | 81 | 71 | 84 | 79 | 88 | 80 | 91 | 109 |
| **Antigenically novel, high immune escape, high virulence** | Yes | High | 7 | 8 | 8 | 9 | 7 | 9 | 8 | 10 | 9 | 10 | 9 | 11 | 13 | 92 | 105 | 97 | 108 | 95 | 106 | 99 | 109 | 103 | 111 | 106 | 113 | 122 |
| | Yes | Low | 8 | 9 | 9 | 11 | 8 | 10 | 9 | 10 | 10 | 11 | 11 | 12 | 15 | 96 | 108 | 101 | 113 | 98 | 111 | 103 | 114 | 108 | 117 | 110 | 118 | 130 |
| | No | High | 9 | 11 | 9 | 12 | 9 | 11 | 9 | 12 | 12 | 13 | 12 | 13 | 16 | 95 | 108 | 101 | 112 | 99 | 111 | 103 | 114 | 109 | 116 | 111 | 117 | 130 |
| | No | Low | 10 | 13 | 11 | 14 | 11 | 13 | 11 | 14 | 13 | 15 | 14 | 16 | 19 | 97 | 114 | 104 | 117 | 101 | 115 | 107 | 118 | 112 | 122 | 115 | 124 | 137 |

*OT: Omicron-targeted vaccine in Q4 2022; 2*OT: Omicron-targeted vaccines in Q4 2022 and Q2 2023; OT+M: Omicron-targeted vaccine in Q4 2022 and multivalent vaccine in Q2 2023; 30+: administered to people aged ≥30 years; 60+ administered to people aged ≥60 years; H: high coverage; L: low coverage*

### Supplementary Figure 7: Heat map of mean net health expenditure for each of 936 scenarios over a 12 month timeline (2021 AUD; millions)

| | | PHSMs | Higher Stringency | | | | | | | | | | | | | Lower Stringency | | | | | | | | | | | | |
|---|---|---|---|---|---|---|---|---|---|---|---|---|---|---|---|---|---|---|---|---|---|---|---|---|---|---|---|---|
| | | Vaccine schedule | OT+M | | | | 2*OT | | | | OT | | | | Nil | OT+M | | | | 2*OT | | | | OT | | | | Nil |
| | | Vaccine age group | 30+ | | 60+ | | 30+ | | 60+ | | 30+ | | 60+ | | | 30+ | | 60+ | | 30+ | | 60+ | | 30+ | | 60+ | | |
| | | Vaccine uptake | *H* | *L* | *H* | *L* | *H* | *L* | *H* | *L* | *H* | *L* | *H* | *L* | | *H* | *L* | *H* | *L* | *H* | *L* | *H* | *L* | *H* | *L* | *H* | *L* | |
| Variant | Respirator stockpile | Mask wearing | | | | | | | | | | | | | | | | | | | | | | | | | | |
| Omicron BA.4/5 only (no new variant) | Yes | High | 493.58 | 332.22 | 358.97 | 263.99 | 461.94 | 311.93 | 343.25 | 251.52 | 311.23 | 210.98 | 243.55 | 180.02 | 60.91 | 457.10 | 293.00 | 319.45 | 222.96 | 422.61 | 274.58 | 303.75 | 211.95 | 266.69 | 164.29 | 203.33 | 135.60 | -2.14 |
| | Yes | Low | 493.41 | 332.46 | 359.31 | 264.97 | 459.97 | 313.07 | 343.26 | 253.42 | 311.79 | 213.92 | 245.75 | 180.39 | 58.37 | 457.64 | 294.30 | 320.13 | 222.74 | 423.02 | 275.91 | 303.77 | 212.35 | 266.68 | 164.55 | 203.70 | 135.23 | -2.04 |
| | No | High | 442.52 | 272.77 | 303.96 | 201.68 | 410.58 | 253.17 | 286.84 | 189.96 | 247.66 | 143.20 | 184.14 | 111.59 | -26.99 | 449.22 | 283.23 | 311.75 | 212.34 | 415.26 | 264.95 | 295.29 | 201.67 | 255.51 | 150.88 | 192.15 | 122.27 | -21.16 |
| | No | Low | 442.30 | 272.49 | 303.77 | 201.89 | 410.42 | 254.38 | 287.56 | 190.87 | 247.13 | 144.08 | 184.45 | 113.08 | -26.16 | 449.68 | 283.12 | 311.10 | 212.68 | 415.06 | 264.85 | 295.08 | 201.69 | 255.15 | 151.25 | 191.85 | 122.15 | -21.59 |
| Antigenically Omicron-like, low immune escape, low virulence | Yes | High | 531.56 | 361.82 | 397.74 | 290.93 | 499.49 | 342.26 | 381.83 | 280.80 | 335.33 | 234.19 | 273.62 | 202.64 | 78.66 | 482.87 | 303.13 | 341.61 | 231.56 | 451.36 | 284.80 | 324.90 | 221.05 | 292.01 | 175.05 | 221.85 | 139.87 | -21.69 |
| | Yes | Low | 532.81 | 360.20 | 395.40 | 291.32 | 500.23 | 342.48 | 380.80 | 280.90 | 338.25 | 231.37 | 272.79 | 202.37 | 74.51 | 482.75 | 303.44 | 341.41 | 231.06 | 451.91 | 285.43 | 324.61 | 220.42 | 292.60 | 175.33 | 222.37 | 139.96 | -20.77 |
| | No | High | 464.16 | 272.81 | 317.60 | 199.43 | 431.18 | 254.72 | 301.25 | 189.07 | 256.34 | 137.23 | 188.64 | 101.77 | -55.83 | 472.08 | 287.70 | 329.78 | 214.95 | 440.90 | 269.73 | 312.45 | 203.49 | 278.46 | 158.81 | 209.04 | 121.70 | -49.17 |
| | No | Low | 464.13 | 274.66 | 318.89 | 199.78 | 431.27 | 255.65 | 301.65 | 189.66 | 257.20 | 138.59 | 188.41 | 104.40 | -54.90 | 472.04 | 288.13 | 328.63 | 215.28 | 441.29 | 270.30 | 312.52 | 203.54 | 278.07 | 158.88 | 208.97 | 122.56 | -48.99 |
| Antigenically novel, low immune escape, low virulence | Yes | High | 588.61 | 411.18 | 451.31 | 340.78 | 558.69 | 394.94 | 435.31 | 331.26 | 381.12 | 282.83 | 320.72 | 249.23 | 127.37 | 509.93 | 328.83 | 369.52 | 259.05 | 480.07 | 312.14 | 354.04 | 247.84 | 312.44 | 198.69 | 248.85 | 167.59 | -9.90 |
| | Yes | Low | 587.76 | 412.93 | 451.29 | 340.78 | 558.54 | 398.67 | 436.18 | 331.85 | 379.66 | 282.58 | 317.96 | 248.95 | 128.35 | 511.39 | 330.10 | 370.41 | 258.77 | 480.34 | 312.73 | 354.41 | 248.02 | 313.50 | 199.81 | 248.75 | 165.39 | -6.95 |
| | No | High | 475.69 | 273.32 | 327.11 | 199.03 | 443.44 | 255.59 | 309.00 | 187.76 | 254.44 | 131.14 | 184.30 | 96.98 | -65.98 | 485.96 | 296.00 | 342.20 | 219.13 | 455.46 | 277.19 | 326.18 | 209.26 | 287.39 | 164.16 | 220.68 | 128.57 | -57.51 |
| | No | Low | 475.79 | 274.95 | 328.87 | 200.18 | 444.92 | 256.60 | 312.01 | 189.27 | 257.46 | 133.08 | 186.30 | 98.79 | -64.53 | 486.50 | 294.94 | 342.23 | 220.32 | 456.28 | 277.00 | 325.52 | 209.80 | 288.65 | 163.33 | 220.81 | 127.74 | -58.96 |
| Antigenically Omicron-like, high immune escape, low virulence | Yes | High | 580.05 | 397.91 | 439.21 | 327.44 | 545.12 | 377.99 | 423.61 | 317.36 | 367.57 | 262.46 | 307.05 | 233.05 | 100.29 | 501.35 | 310.01 | 357.89 | 235.47 | 466.93 | 290.06 | 341.27 | 224.81 | 298.66 | 177.20 | 233.75 | 142.29 | -43.82 |
| | Yes | Low | 580.18 | 399.34 | 441.33 | 330.92 | 548.18 | 381.81 | 425.28 | 318.87 | 369.73 | 263.51 | 305.76 | 233.80 | 100.96 | 501.41 | 310.73 | 358.33 | 237.30 | 467.78 | 289.72 | 342.12 | 225.44 | 300.41 | 176.92 | 234.35 | 142.01 | -46.41 |
| | No | High | 465.98 | 257.23 | 312.60 | 181.17 | 434.24 | 237.44 | 296.08 | 168.00 | 243.40 | 109.55 | 170.01 | 77.04 | -98.14 | 476.43 | 274.63 | 328.86 | 197.86 | 443.96 | 254.19 | 311.79 | 185.87 | 275.80 | 139.92 | 204.26 | 102.71 | -101.68 |
| | No | Low | 466.27 | 257.10 | 314.68 | 180.78 | 434.06 | 238.36 | 298.10 | 169.54 | 244.07 | 111.64 | 170.66 | 77.03 | -97.35 | 476.11 | 275.40 | 328.29 | 197.66 | 442.81 | 255.10 | 311.72 | 187.14 | 273.97 | 140.69 | 204.23 | 104.01 | -97.61 |
| Antigenically novel, high immune escape, low virulence | Yes | High | 634.62 | 453.47 | 493.05 | 383.91 | 602.11 | 435.11 | 473.67 | 369.69 | 413.62 | 318.20 | 350.50 | 284.74 | 153.75 | 535.64 | 330.64 | 388.95 | 260.85 | 504.70 | 311.08 | 371.10 | 251.43 | 328.67 | 192.00 | 255.61 | 164.31 | -33.29 |
| | Yes | Low | 636.91 | 457.66 | 497.94 | 383.65 | 606.67 | 436.39 | 479.81 | 374.10 | 419.34 | 319.32 | 357.25 | 286.44 | 158.83 | 534.77 | 330.95 | 389.91 | 262.96 | 503.82 | 312.11 | 372.57 | 252.15 | 326.60 | 193.09 | 259.68 | 167.89 | -33.29 |
| | No | High | 462.63 | 245.74 | 307.78 | 168.80 | 426.94 | 224.71 | 288.49 | 156.92 | 223.29 | 92.78 | 150.58 | 57.30 | -115.04 | 476.36 | 263.45 | 324.45 | 187.11 | 445.97 | 242.45 | 308.62 | 175.34 | 269.53 | 125.74 | 193.27 | 90.92 | -119.89 |
| | No | Low | 463.83 | 247.14 | 310.31 | 168.74 | 427.79 | 226.94 | 290.06 | 157.53 | 225.49 | 94.18 | 152.59 | 58.91 | -115.51 | 476.23 | 263.96 | 326.27 | 187.83 | 445.93 | 244.11 | 309.72 | 176.91 | 268.87 | 125.35 | 196.84 | 89.85 | -121.67 |
| Antigenically Omicron-like, low immune escape, high virulence | Yes | High | 571.80 | 405.75 | 430.25 | 334.58 | 540.29 | 386.08 | 411.38 | 329.04 | 390.76 | 289.92 | 328.35 | 258.44 | 141.03 | 373.69 | 128.21 | 205.34 | 43.50 | 342.19 | 104.48 | 191.48 | 31.83 | 134.10 | -28.65 | 58.57 | -67.11 | -271.18 |
| | Yes | Low | 581.96 | 420.28 | 442.95 | 356.26 | 550.47 | 401.76 | 430.37 | 345.65 | 406.12 | 306.98 | 343.91 | 283.63 | 161.45 | 372.30 | 121.53 | 206.63 | 41.35 | 340.08 | 96.30 | 190.86 | 27.04 | 130.70 | -30.10 | 51.85 | -69.03 | -281.03 |
| | No | High | 278.42 | 53.51 | 107.06 | -31.01 | 246.20 | 34.51 | 90.39 | -42.71 | 45.09 | -95.03 | -34.59 | -129.38 | -297.41 | 280.16 | -12.14 | 97.33 | -107.08 | 248.17 | -33.13 | 80.11 | -118.12 | -2.03 | -195.92 | -86.81 | -240.71 | -502.54 |
| | No | Low | 277.60 | 50.24 | 108.29 | -35.21 | 244.98 | 30.85 | 89.95 | -46.87 | 40.30 | -98.38 | -34.99 | -138.93 | -312.12 | 280.98 | -14.48 | 95.59 | -109.25 | 247.93 | -35.32 | 79.20 | -121.71 | -2.96 | -198.52 | -86.05 | -245.27 | -511.99 |
| Antigenically novel, low immune escape, high virulence | Yes | High | 685.93 | 510.93 | 546.52 | 440.02 | 666.58 | 502.20 | 538.93 | 435.12 | 491.25 | 388.66 | 426.63 | 352.45 | 223.82 | 436.64 | 187.96 | 273.62 | 108.83 | 417.36 | 179.63 | 266.57 | 107.23 | 184.58 | 31.44 | 114.14 | -6.89 | -217.24 |
| | Yes | Low | 710.88 | 536.67 | 579.78 | 471.55 | 698.35 | 532.41 | 577.35 | 468.08 | 524.76 | 419.97 | 464.17 | 389.22 | 257.71 | 437.59 | 191.51 | 271.17 | 107.81 | 416.28 | 179.49 | 266.37 | 103.26 | 183.60 | 30.50 | 109.75 | -6.99 | -226.68 |
| | No | High | 259.95 | 27.45 | 89.36 | -58.09 | 229.62 | 11.16 | 76.28 | -64.03 | 13.06 | -123.71 | -62.09 | -164.40 | -334.44 | 253.44 | -52.78 | 68.54 | -147.08 | 225.93 | -67.18 | 57.96 | -152.61 | -47.85 | -247.75 | -134.00 | -289.16 | -561.57 |
| | No | Low | 260.36 | 22.31 | 87.02 | -62.80 | 230.89 | 6.14 | 76.11 | -70.64 | 9.59 | -130.86 | -66.48 | -170.29 | -349.48 | 253.67 | -57.62 | 69.40 | -148.67 | 225.67 | -70.06 | 54.49 | -156.90 | -52.45 | -255.04 | -135.94 | -294.70 | -575.98 |
| Antigenically Omicron-like, high immune escape, high virulence | Yes | High | 688.54 | 500.19 | 542.73 | 429.97 | 654.50 | 479.47 | 530.99 | 417.21 | 473.16 | 370.16 | 415.17 | 337.54 | 193.23 | 425.22 | 156.64 | 252.74 | 77.79 | 390.25 | 138.11 | 239.32 | 66.77 | 153.43 | -11.59 | 79.97 | -45.55 | -272.88 |
| | Yes | Low | 722.48 | 530.81 | 578.19 | 458.19 | 685.86 | 513.76 | 561.79 | 451.38 | 507.90 | 397.97 | 448.55 | 366.44 | 229.03 | 421.13 | 155.58 | 257.61 | 75.40 | 391.84 | 140.50 | 241.28 | 61.65 | 158.61 | -11.08 | 84.55 | -50.85 | -280.74 |
| | No | High | 245.46 | 5.47 | 73.38 | -78.81 | 212.19 | -15.50 | 56.34 | -92.15 | -5.67 | -153.14 | -83.16 | -192.30 | -374.43 | 230.22 | -93.29 | 41.27 | -186.80 | 198.32 | -110.78 | 24.27 | -199.30 | -82.17 | -295.77 | -168.13 | -338.98 | -632.71 |
| | No | Low | 246.21 | -0.73 | 71.08 | -89.29 | 213.21 | -20.45 | 54.49 | -99.07 | -10.73 | -162.93 | -89.21 | -202.53 | -392.73 | 229.71 | -97.70 | 41.58 | -191.18 | 198.63 | -117.46 | 22.77 | -204.38 | -85.94 | -303.24 | -171.95 | -346.13 | -650.54 |
| Antigenically novel, high immune escape, high virulence | Yes | High | 776.30 | 573.32 | 626.47 | 498.11 | 759.27 | 562.50 | 620.23 | 496.98 | 553.81 | 431.70 | 486.04 | 402.09 | 247.46 | 497.66 | 222.65 | 324.60 | 138.74 | 469.58 | 212.55 | 320.83 | 136.06 | 210.46 | 47.09 | 138.06 | 11.78 | -231.41 |
| | Yes | Low | 829.48 | 628.82 | 672.54 | 554.97 | 806.31 | 614.50 | 670.37 | 552.15 | 603.16 | 489.82 | 533.75 | 449.64 | 293.86 | 503.87 | 227.55 | 332.72 | 145.34 | 479.54 | 217.24 | 325.63 | 141.98 | 219.96 | 49.40 | 138.53 | 8.34 | -230.34 |
| | No | High | 214.17 | -30.09 | 44.58 | -114.34 | 182.62 | -46.46 | 30.69 | -123.51 | -52.15 | -194.48 | -125.62 | -233.69 | -418.98 | 181.49 | -152.20 | -8.92 | -245.55 | 148.01 | -167.62 | -24.06 | -254.41 | -154.26 | -363.62 | -239.74 | -409.22 | -707.65 |
| | No | Low | 214.10 | -38.54 | 39.28 | -124.13 | 181.09 | -55.47 | 25.02 | -133.63 | -57.97 | -209.37 | -135.36 | -243.65 | -440.40 | 181.66 | -157.67 | -10.07 | -256.96 | 147.78 | -174.30 | -26.28 | -263.75 | -161.56 | -374.51 | -247.16 | -424.09 | -728.05 |

*OT: Omicron-targeted vaccine in Q4 2022; 2*OT: Omicron-targeted vaccines in Q4 2022 and Q2 2023; OT+M: Omicron-targeted vaccine in Q4 2022 and multivalent vaccine in Q2 2023; 30+: administered to people aged ≥30 years; 60+ administered to people aged ≥60 years; H: high coverage; L: low coverage*

### Supplementary Figure 8: Heat map of mean health-adjusted life years lost for each of 936 scenarios over a 12 month timeline

| | | PHSMs | Higher Stringency | | | | | | | | | | | | | Lower Stringency | | | | | | | | | | | | |
|---|---|---|---|---|---|---|---|---|---|---|---|---|---|---|---|---|---|---|---|---|---|---|---|---|---|---|---|---|
| | | Vaccine schedule | OT+M | | | | 2*OT | | | | OT | | | | Nil | OT+M | | | | 2*OT | | | | OT | | | | Nil |
| | | Vaccine age group | 30+ | | 60+ | | 30+ | | 60+ | | 30+ | | 60+ | | | 30+ | | 60+ | | 30+ | | 60+ | | 30+ | | 60+ | | |
| | | Vaccine uptake | *H* | *L* | *H* | *L* | *H* | *L* | *H* | *L* | *H* | *L* | *H* | *L* | | *H* | *L* | *H* | *L* | *H* | *L* | *H* | *L* | *H* | *L* | *H* | *L* | |
| Variant | Respirator stockpile | Mask wearing | | | | | | | | | | | | | | | | | | | | | | | | | | |
| Omicron BA.4/5 only (no new variant) | Yes | High | 15046 | 18292 | 17695 | 19684 | 15075 | 18304 | 17728 | 19700 | 17672 | 19849 | 18919 | 20508 | 23153 | 16417 | 20211 | 19400 | 21896 | 16422 | 20314 | 19395 | 21803 | 21127 | 23230 | 21654 | 23591 | 27217 |
| | Yes | Low | 15106 | 18328 | 17767 | 19657 | 15088 | 18333 | 17780 | 19723 | 17872 | 19942 | 19009 | 20521 | 23338 | 16456 | 20181 | 19443 | 21851 | 16466 | 20300 | 19416 | 21786 | 21181 | 23231 | 21671 | 23582 | 27282 |
| | No | High | 15149 | 18306 | 17747 | 19715 | 15114 | 18282 | 17772 | 19685 | 17794 | 19936 | 18974 | 20610 | 23377 | 16447 | 20224 | 19430 | 21873 | 16445 | 20366 | 19394 | 21822 | 21151 | 23214 | 21663 | 23620 | 27234 |
| | No | Low | 15172 | 18336 | 17832 | 19815 | 15179 | 18383 | 17836 | 19721 | 17957 | 20016 | 19049 | 20601 | 23496 | 16462 | 20235 | 19410 | 21883 | 16450 | 20321 | 19398 | 21797 | 21203 | 23179 | 21700 | 23623 | 27256 |
| Antigenically Omicron-like, low immune escape, low virulence | Yes | High | 18993 | 22461 | 21677 | 23718 | 18903 | 22380 | 21682 | 23688 | 21455 | 23854 | 22720 | 24503 | 27353 | 20667 | 24866 | 23435 | 26348 | 20604 | 24857 | 23420 | 26370 | 24539 | 27542 | 25594 | 28136 | 32910 |
| | Yes | Low | 18972 | 22523 | 21776 | 23928 | 19003 | 22447 | 21806 | 23814 | 21455 | 24077 | 22827 | 24660 | 27615 | 20651 | 24902 | 23396 | 26340 | 20636 | 24857 | 23384 | 26372 | 24588 | 27565 | 25609 | 28153 | 32885 |
| | No | High | 19051 | 22503 | 21745 | 23812 | 19006 | 22499 | 21762 | 23757 | 21530 | 24026 | 22859 | 24701 | 27592 | 20651 | 24817 | 23421 | 26335 | 20616 | 24868 | 23421 | 26398 | 24562 | 27525 | 25575 | 28232 | 32974 |
| | No | Low | 19012 | 22578 | 21783 | 23944 | 18988 | 22522 | 21794 | 23869 | 21601 | 24145 | 22926 | 24731 | 27851 | 20639 | 24867 | 23405 | 26406 | 20655 | 24832 | 23428 | 26391 | 24611 | 27571 | 25582 | 28163 | 32960 |
| Antigenically novel, low immune escape, low virulence | Yes | High | 22137 | 25625 | 24959 | 26917 | 22368 | 25649 | 25033 | 26877 | 24566 | 26980 | 25977 | 27614 | 30357 | 24059 | 28705 | 27069 | 30153 | 24495 | 29057 | 27378 | 30367 | 27969 | 31133 | 29254 | 31860 | 37254 |
| | Yes | Low | 22247 | 25772 | 25041 | 27199 | 22593 | 25858 | 25219 | 27103 | 24847 | 27217 | 26177 | 27921 | 30785 | 24074 | 28604 | 27065 | 30187 | 24470 | 29010 | 27391 | 30408 | 27971 | 31149 | 29260 | 31975 | 37224 |
| | No | High | 22191 | 25670 | 25149 | 27201 | 22555 | 25822 | 25181 | 27125 | 24818 | 27182 | 26203 | 27840 | 30728 | 24085 | 28598 | 26990 | 30174 | 24507 | 29003 | 27372 | 30384 | 28015 | 31150 | 29248 | 31923 | 37153 |
| | No | Low | 22313 | 25821 | 25190 | 27279 | 22628 | 25949 | 25240 | 27317 | 25010 | 27369 | 26290 | 28012 | 31017 | 24078 | 28658 | 26983 | 30187 | 24502 | 28989 | 27309 | 30345 | 28008 | 31230 | 29223 | 31912 | 37241 |
| Antigenically Omicron-like, high immune escape, low virulence | Yes | High | 22881 | 26360 | 25684 | 27709 | 22793 | 26413 | 25648 | 27718 | 25193 | 27701 | 26659 | 28369 | 31373 | 25083 | 29849 | 28109 | 31465 | 25085 | 29894 | 28081 | 31482 | 28492 | 32210 | 30141 | 32995 | 38689 |
| | Yes | Low | 23045 | 26607 | 25829 | 28025 | 22957 | 26631 | 25850 | 28014 | 25382 | 28081 | 26829 | 28777 | 31789 | 25057 | 29834 | 28147 | 31439 | 25054 | 29861 | 28137 | 31503 | 28461 | 32227 | 30135 | 33071 | 38894 |
| | No | High | 23013 | 26545 | 25806 | 27997 | 22978 | 26586 | 25829 | 27976 | 25401 | 27962 | 26891 | 28705 | 31641 | 25055 | 29826 | 28177 | 31531 | 25118 | 29855 | 28111 | 31500 | 28500 | 32240 | 30204 | 33105 | 38774 |
| | No | Low | 23093 | 26769 | 25933 | 28096 | 23062 | 26750 | 25987 | 28150 | 25528 | 28222 | 27036 | 28894 | 32047 | 25091 | 29783 | 28171 | 31468 | 25108 | 29858 | 28125 | 31426 | 28536 | 32267 | 30113 | 33082 | 38658 |
| Antigenically novel, high immune escape, low virulence | Yes | High | 26170 | 29448 | 28973 | 30719 | 26338 | 29501 | 28850 | 30697 | 28470 | 30698 | 29866 | 31411 | 34181 | 28887 | 33935 | 32133 | 35480 | 29356 | 34328 | 32351 | 35731 | 32489 | 36430 | 34373 | 37289 | 42823 |
| | Yes | Low | 26338 | 29859 | 29190 | 31156 | 26618 | 29907 | 29135 | 31164 | 28808 | 31142 | 30219 | 31970 | 34763 | 28937 | 33899 | 32074 | 35441 | 29412 | 34223 | 32396 | 35659 | 32571 | 36370 | 34269 | 37221 | 42985 |
| | No | High | 26426 | 29810 | 29159 | 31140 | 26623 | 29849 | 29077 | 31072 | 28737 | 31142 | 30148 | 31816 | 34668 | 28886 | 33919 | 32112 | 35533 | 29430 | 34253 | 32416 | 35655 | 32518 | 36342 | 34327 | 37303 | 42904 |
| | No | Low | 26523 | 30051 | 29308 | 31429 | 26733 | 30079 | 29315 | 31367 | 29069 | 31505 | 30508 | 32121 | 35222 | 29011 | 34008 | 32160 | 35473 | 29398 | 34274 | 32419 | 35645 | 32487 | 36451 | 34377 | 37326 | 43083 |
| Antigenically Omicron-like, low immune escape, high virulence | Yes | High | 33813 | 36181 | 37984 | 38055 | 33804 | 36262 | 38088 | 37950 | 34516 | 36422 | 36602 | 37349 | 38719 | 52811 | 58595 | 59806 | 61836 | 52502 | 58782 | 59709 | 61902 | 55589 | 60244 | 59010 | 61653 | 65826 |
| | Yes | Low | 35098 | 37756 | 39645 | 39877 | 35132 | 37743 | 39713 | 39849 | 35992 | 38074 | 38373 | 39065 | 40843 | 53224 | 59678 | 60410 | 62908 | 53170 | 59679 | 60453 | 62839 | 56551 | 61131 | 60238 | 62852 | 67766 |
| | No | High | 34821 | 37359 | 39287 | 39322 | 34778 | 37252 | 39208 | 39333 | 35645 | 37845 | 38007 | 38529 | 40318 | 53238 | 59319 | 60191 | 62790 | 53018 | 59539 | 60053 | 62829 | 56360 | 60922 | 59931 | 62603 | 67361 |
| | No | Low | 35754 | 38701 | 40408 | 40837 | 35858 | 38677 | 40370 | 40904 | 36885 | 39110 | 39104 | 40342 | 42140 | 53415 | 60166 | 60872 | 63629 | 53319 | 60259 | 60712 | 63588 | 56966 | 61914 | 60627 | 63677 | 68703 |
| Antigenically novel, low immune escape, high virulence | Yes | High | 37549 | 39834 | 41558 | 41738 | 36966 | 39420 | 40773 | 41257 | 38097 | 40071 | 39930 | 40932 | 42501 | 60675 | 66225 | 67322 | 69176 | 60304 | 65801 | 66675 | 68622 | 63300 | 67379 | 66316 | 68526 | 72820 |
| | Yes | Low | 39214 | 41950 | 43743 | 43777 | 38781 | 41606 | 42833 | 43405 | 40176 | 42269 | 42149 | 43261 | 44963 | 61850 | 67812 | 68630 | 71046 | 61483 | 67482 | 67984 | 70472 | 65051 | 69245 | 68266 | 70746 | 75389 |
| | No | High | 38981 | 41358 | 43175 | 43468 | 38479 | 41209 | 42467 | 42993 | 39910 | 41835 | 41602 | 42755 | 44722 | 61439 | 67310 | 68419 | 70718 | 61153 | 67022 | 67753 | 70130 | 64473 | 68919 | 67725 | 70272 | 74859 |
| | No | Low | 40311 | 43285 | 44967 | 45329 | 39877 | 42975 | 44119 | 44882 | 41383 | 43556 | 43431 | 44650 | 46754 | 62336 | 68586 | 69390 | 71896 | 62166 | 68353 | 68919 | 71454 | 65710 | 70364 | 68971 | 71907 | 76709 |
| Antigenically Omicron-like, high immune escape, high virulence | Yes | High | 37581 | 40244 | 41594 | 42168 | 37524 | 40275 | 41572 | 42219 | 38753 | 40828 | 40878 | 41680 | 43536 | 61475 | 67469 | 68456 | 70550 | 61658 | 67492 | 68362 | 70467 | 64595 | 69029 | 67867 | 70518 | 74793 |
| | Yes | Low | 39484 | 42375 | 43782 | 44411 | 39471 | 42357 | 43814 | 44444 | 40854 | 43140 | 42939 | 44072 | 46102 | 62709 | 69173 | 69702 | 72532 | 62632 | 69214 | 69590 | 72497 | 66317 | 70866 | 69661 | 72768 | 77432 |
| | No | High | 39087 | 41978 | 43435 | 43931 | 39118 | 42145 | 43365 | 44001 | 40452 | 42808 | 42479 | 43687 | 45592 | 62532 | 68911 | 69414 | 72170 | 62412 | 68854 | 69439 | 72062 | 65942 | 70654 | 69387 | 72273 | 76975 |
| | No | Low | 40572 | 43691 | 45119 | 45981 | 40616 | 43761 | 45093 | 45944 | 42082 | 44583 | 44387 | 45619 | 47786 | 63397 | 69996 | 70412 | 73488 | 63280 | 70159 | 70548 | 73597 | 67120 | 72176 | 70768 | 73844 | 79162 |
| Antigenically novel, high immune escape, high virulence | Yes | High | 41192 | 43898 | 45149 | 45761 | 40683 | 43496 | 44375 | 45297 | 42491 | 44470 | 44079 | 45191 | 47297 | 68451 | 74021 | 74969 | 76987 | 68111 | 73519 | 74086 | 76325 | 71618 | 75356 | 74395 | 76677 | 81283 |
| | Yes | Low | 43442 | 46551 | 47833 | 48412 | 43087 | 46265 | 47076 | 48045 | 44956 | 47132 | 46932 | 47992 | 50463 | 70303 | 76596 | 77309 | 79710 | 70007 | 76404 | 76383 | 79293 | 73835 | 78419 | 77205 | 80048 | 84934 |
| | No | High | 43143 | 46120 | 47183 | 47898 | 42613 | 45692 | 46442 | 47614 | 44555 | 46807 | 46342 | 47573 | 49947 | 70194 | 76014 | 76875 | 79052 | 69699 | 75649 | 75799 | 78742 | 73516 | 77727 | 76371 | 79522 | 84379 |
| | No | Low | 45014 | 48312 | 49448 | 50361 | 44515 | 47975 | 48735 | 49953 | 46698 | 49229 | 48691 | 49944 | 52500 | 71646 | 78225 | 78688 | 81443 | 71104 | 77906 | 77850 | 80874 | 75455 | 80220 | 78623 | 81858 | 87057 |

*OT: Omicron-targeted vaccine in Q4 2022; 2*OT: Omicron-targeted vaccines in Q4 2022 and Q2 2023; OT+M: Omicron-targeted vaccine in Q4 2022 and multivalent vaccine in Q2 2023; 30+: administered to people aged ≥30 years; 60+ administered to people aged ≥60 years; H: high coverage; L: low coverage*

### Supplementary Figure 9: Heat map of mean <u>net monetary benefit from a health system perspective</u> (with a health-adjusted life year valued at AUD 70,000, 3% discount rate on both health-adjusted life years and costs; billions) for each of 936 scenarios over a 12 month timeline

| | | PHSMs | Higher Stringency | | | | | | | | | | | | | Lower Stringency | | | | | | | | | | | | |
|---|---|---|---|---|---|---|---|---|---|---|---|---|---|---|---|---|---|---|---|---|---|---|---|---|---|---|---|---|
| | | Vaccine schedule | OT+M | | | | 2*OT | | | | OT | | | | Nil | OT+M | | | | 2*OT | | | | OT | | | | Nil |
| | | Vaccine age group | 30+ | | 60+ | | 30+ | | 60+ | | 30+ | | 60+ | | | 30+ | | 60+ | | 30+ | | 60+ | | 30+ | | 60+ | | |
| | | Vaccine uptake | *H* | *L* | *H* | *L* | *H* | *L* | *H* | *L* | *H* | *L* | *H* | *L* | | *H* | *L* | *H* | *L* | *H* | *L* | *H* | *L* | *H* | *L* | *H* | *L* | |
| Variant | Respirator stockpile | Mask wearing | | | | | | | | | | | | | | | | | | | | | | | | | | |
| Omicron BA.4/5 only (no new variant) | Yes | High | -1.55 | -1.61 | -1.60 | -1.64 | -1.52 | -1.59 | -1.58 | -1.63 | -1.55 | -1.60 | -1.57 | -1.62 | -1.68 | -1.61 | -1.71 | -1.68 | -1.76 | -1.57 | -1.70 | -1.66 | -1.74 | -1.75 | -1.79 | -1.72 | -1.79 | -1.90 |
| | Yes | Low | -1.55 | -1.62 | -1.60 | -1.64 | -1.52 | -1.60 | -1.59 | -1.63 | -1.56 | -1.61 | -1.58 | -1.62 | -1.69 | -1.61 | -1.71 | -1.68 | -1.75 | -1.58 | -1.70 | -1.66 | -1.74 | -1.75 | -1.79 | -1.72 | -1.79 | -1.91 |
| | No | High | -1.50 | -1.55 | -1.55 | -1.58 | -1.47 | -1.53 | -1.53 | -1.57 | -1.49 | -1.54 | -1.51 | -1.55 | -1.61 | -1.60 | -1.70 | -1.67 | -1.74 | -1.57 | -1.69 | -1.65 | -1.73 | -1.74 | -1.78 | -1.71 | -1.78 | -1.89 |
| | No | Low | -1.50 | -1.56 | -1.55 | -1.59 | -1.47 | -1.54 | -1.54 | -1.57 | -1.50 | -1.55 | -1.52 | -1.56 | -1.62 | -1.60 | -1.70 | -1.67 | -1.74 | -1.57 | -1.69 | -1.65 | -1.73 | -1.74 | -1.77 | -1.71 | -1.78 | -1.89 |
| Antigenically Omicron-like, low immune escape, low virulence | Yes | High | -1.86 | -1.93 | -1.92 | -1.95 | -1.82 | -1.91 | -1.90 | -1.94 | -1.84 | -1.90 | -1.86 | -1.92 | -1.99 | -1.93 | -2.04 | -1.98 | -2.08 | -1.89 | -2.02 | -1.96 | -2.07 | -2.01 | -2.10 | -2.01 | -2.11 | -2.28 |
| | Yes | Low | -1.86 | -1.94 | -1.92 | -1.97 | -1.83 | -1.91 | -1.91 | -1.95 | -1.84 | -1.92 | -1.87 | -1.93 | -2.01 | -1.93 | -2.05 | -1.98 | -2.07 | -1.90 | -2.03 | -1.96 | -2.07 | -2.01 | -2.10 | -2.01 | -2.11 | -2.28 |
| | No | High | -1.80 | -1.85 | -1.84 | -1.87 | -1.76 | -1.83 | -1.82 | -1.85 | -1.76 | -1.82 | -1.79 | -1.83 | -1.88 | -1.92 | -2.02 | -1.97 | -2.06 | -1.88 | -2.01 | -1.95 | -2.05 | -2.00 | -2.09 | -2.00 | -2.10 | -2.26 |
| | No | Low | -1.79 | -1.86 | -1.84 | -1.88 | -1.76 | -1.83 | -1.83 | -1.86 | -1.77 | -1.83 | -1.79 | -1.84 | -1.89 | -1.92 | -2.03 | -1.97 | -2.06 | -1.89 | -2.01 | -1.95 | -2.05 | -2.00 | -2.09 | -2.00 | -2.09 | -2.26 |
| Antigenically novel, low immune escape, low virulence | Yes | High | -2.14 | -2.20 | -2.20 | -2.22 | -2.12 | -2.19 | -2.19 | -2.21 | -2.10 | -2.17 | -2.14 | -2.18 | -2.25 | -2.19 | -2.34 | -2.26 | -2.37 | -2.19 | -2.35 | -2.27 | -2.37 | -2.27 | -2.38 | -2.30 | -2.40 | -2.60 |
| | Yes | Low | -2.15 | -2.22 | -2.20 | -2.24 | -2.14 | -2.21 | -2.20 | -2.23 | -2.12 | -2.19 | -2.15 | -2.20 | -2.28 | -2.20 | -2.33 | -2.26 | -2.37 | -2.19 | -2.34 | -2.27 | -2.38 | -2.27 | -2.38 | -2.30 | -2.40 | -2.60 |
| | No | High | -2.03 | -2.07 | -2.09 | -2.10 | -2.02 | -2.06 | -2.07 | -2.09 | -1.99 | -2.03 | -2.02 | -2.05 | -2.08 | -2.17 | -2.30 | -2.23 | -2.33 | -2.17 | -2.31 | -2.24 | -2.34 | -2.25 | -2.34 | -2.27 | -2.36 | -2.54 |
| | No | Low | -2.04 | -2.08 | -2.09 | -2.11 | -2.03 | -2.07 | -2.08 | -2.10 | -2.01 | -2.05 | -2.03 | -2.06 | -2.11 | -2.17 | -2.30 | -2.23 | -2.33 | -2.17 | -2.31 | -2.24 | -2.33 | -2.25 | -2.35 | -2.27 | -2.36 | -2.55 |
| Antigenically Omicron-like, high immune escape, low virulence | Yes | High | -2.18 | -2.24 | -2.24 | -2.27 | -2.14 | -2.23 | -2.22 | -2.26 | -2.13 | -2.20 | -2.17 | -2.22 | -2.30 | -2.26 | -2.40 | -2.33 | -2.44 | -2.22 | -2.38 | -2.31 | -2.43 | -2.29 | -2.43 | -2.34 | -2.45 | -2.66 |
| | Yes | Low | -2.19 | -2.26 | -2.25 | -2.29 | -2.16 | -2.25 | -2.23 | -2.28 | -2.15 | -2.23 | -2.18 | -2.25 | -2.33 | -2.26 | -2.40 | -2.33 | -2.44 | -2.22 | -2.38 | -2.31 | -2.43 | -2.29 | -2.43 | -2.34 | -2.46 | -2.68 |
| | No | High | -2.08 | -2.12 | -2.12 | -2.14 | -2.04 | -2.10 | -2.10 | -2.13 | -2.02 | -2.07 | -2.05 | -2.09 | -2.12 | -2.23 | -2.36 | -2.30 | -2.41 | -2.20 | -2.34 | -2.28 | -2.39 | -2.27 | -2.40 | -2.32 | -2.42 | -2.61 |
| | No | Low | -2.08 | -2.13 | -2.13 | -2.15 | -2.05 | -2.11 | -2.12 | -2.14 | -2.03 | -2.09 | -2.06 | -2.10 | -2.15 | -2.23 | -2.36 | -2.30 | -2.40 | -2.20 | -2.35 | -2.28 | -2.39 | -2.27 | -2.40 | -2.31 | -2.42 | -2.61 |
| Antigenically novel, high immune escape, low virulence | Yes | High | -2.47 | -2.51 | -2.52 | -2.53 | -2.45 | -2.50 | -2.49 | -2.52 | -2.41 | -2.47 | -2.44 | -2.48 | -2.55 | -2.56 | -2.71 | -2.64 | -2.74 | -2.56 | -2.71 | -2.64 | -2.75 | -2.60 | -2.74 | -2.66 | -2.77 | -2.96 |
| | Yes | Low | -2.48 | -2.55 | -2.54 | -2.56 | -2.47 | -2.53 | -2.52 | -2.56 | -2.44 | -2.50 | -2.47 | -2.52 | -2.59 | -2.56 | -2.70 | -2.64 | -2.74 | -2.56 | -2.71 | -2.64 | -2.75 | -2.61 | -2.74 | -2.66 | -2.77 | -2.98 |
| | No | High | -2.31 | -2.33 | -2.35 | -2.35 | -2.29 | -2.31 | -2.32 | -2.33 | -2.23 | -2.27 | -2.26 | -2.28 | -2.31 | -2.50 | -2.64 | -2.57 | -2.67 | -2.51 | -2.64 | -2.58 | -2.67 | -2.55 | -2.67 | -2.60 | -2.70 | -2.88 |
| | No | Low | -2.32 | -2.35 | -2.36 | -2.37 | -2.30 | -2.33 | -2.34 | -2.35 | -2.26 | -2.30 | -2.29 | -2.31 | -2.35 | -2.51 | -2.64 | -2.58 | -2.67 | -2.50 | -2.64 | -2.58 | -2.67 | -2.54 | -2.68 | -2.60 | -2.70 | -2.89 |
| Antigenically Omicron-like, low immune escape, high virulence | Yes | High | -2.94 | -2.94 | -3.09 | -3.00 | -2.91 | -2.92 | -3.08 | -2.99 | -2.81 | -2.84 | -2.89 | -2.87 | -2.85 | -4.07 | -4.23 | -4.39 | -4.37 | -4.02 | -4.22 | -4.37 | -4.36 | -4.03 | -4.19 | -4.19 | -4.25 | -4.34 |
| | Yes | Low | -3.04 | -3.06 | -3.22 | -3.15 | -3.01 | -3.04 | -3.21 | -3.14 | -2.93 | -2.97 | -3.03 | -3.02 | -3.02 | -4.10 | -4.30 | -4.44 | -4.44 | -4.06 | -4.27 | -4.42 | -4.43 | -4.09 | -4.25 | -4.27 | -4.33 | -4.46 |
| | No | High | -2.72 | -2.67 | -2.86 | -2.72 | -2.68 | -2.64 | -2.83 | -2.71 | -2.54 | -2.55 | -2.63 | -2.57 | -2.52 | -4.01 | -4.14 | -4.31 | -4.29 | -3.96 | -4.13 | -4.28 | -4.28 | -3.94 | -4.07 | -4.11 | -4.14 | -4.21 |
| | No | Low | -2.78 | -2.76 | -2.94 | -2.82 | -2.76 | -2.74 | -2.92 | -2.82 | -2.62 | -2.64 | -2.70 | -2.68 | -2.64 | -4.02 | -4.20 | -4.36 | -4.34 | -3.98 | -4.18 | -4.33 | -4.33 | -3.98 | -4.14 | -4.16 | -4.21 | -4.30 |
| Antigenically novel, low immune escape, high virulence | Yes | High | -3.31 | -3.30 | -3.46 | -3.36 | -3.25 | -3.26 | -3.39 | -3.32 | -3.16 | -3.19 | -3.22 | -3.22 | -3.20 | -4.68 | -4.82 | -4.99 | -4.95 | -4.64 | -4.79 | -4.93 | -4.91 | -4.62 | -4.75 | -4.76 | -4.79 | -4.88 |
| | Yes | Low | -3.46 | -3.47 | -3.64 | -3.54 | -3.41 | -3.44 | -3.58 | -3.51 | -3.34 | -3.38 | -3.41 | -3.42 | -3.41 | -4.77 | -4.94 | -5.08 | -5.08 | -4.72 | -4.90 | -5.03 | -5.04 | -4.74 | -4.88 | -4.89 | -4.95 | -5.05 |
| | No | High | -2.99 | -2.92 | -3.11 | -2.98 | -2.92 | -2.90 | -3.05 | -2.95 | -2.81 | -2.80 | -2.85 | -2.83 | -2.80 | -4.55 | -4.66 | -4.86 | -4.80 | -4.51 | -4.62 | -4.80 | -4.76 | -4.47 | -4.58 | -4.61 | -4.63 | -4.68 |
| | No | Low | -3.08 | -3.05 | -3.23 | -3.11 | -3.02 | -3.01 | -3.16 | -3.07 | -2.91 | -2.92 | -2.97 | -2.96 | -2.92 | -4.62 | -4.74 | -4.93 | -4.88 | -4.58 | -4.71 | -4.88 | -4.84 | -4.55 | -4.67 | -4.69 | -4.74 | -4.79 |
| Antigenically Omicron-like, high immune escape, high virulence | Yes | High | -3.32 | -3.32 | -3.45 | -3.38 | -3.28 | -3.30 | -3.44 | -3.37 | -3.19 | -3.23 | -3.28 | -3.26 | -3.24 | -4.73 | -4.88 | -5.04 | -5.02 | -4.71 | -4.86 | -5.02 | -5.00 | -4.68 | -4.82 | -4.83 | -4.89 | -4.96 |
| | Yes | Low | -3.49 | -3.50 | -3.64 | -3.57 | -3.45 | -3.48 | -3.63 | -3.56 | -3.37 | -3.42 | -3.45 | -3.45 | -3.46 | -4.81 | -5.00 | -5.14 | -5.15 | -4.78 | -4.99 | -5.11 | -5.14 | -4.80 | -4.95 | -4.96 | -5.04 | -5.14 |
| | No | High | -2.98 | -2.94 | -3.11 | -3.00 | -2.95 | -2.93 | -3.09 | -2.99 | -2.83 | -2.84 | -2.89 | -2.87 | -2.82 | -4.61 | -4.73 | -4.90 | -4.87 | -4.57 | -4.71 | -4.89 | -4.85 | -4.53 | -4.65 | -4.69 | -4.72 | -4.76 |
| | No | Low | -3.09 | -3.06 | -3.23 | -3.13 | -3.06 | -3.04 | -3.21 | -3.12 | -2.93 | -2.96 | -3.02 | -2.99 | -2.95 | -4.67 | -4.80 | -4.97 | -4.95 | -4.63 | -4.79 | -4.96 | -4.95 | -4.61 | -4.75 | -4.78 | -4.82 | -4.89 |
| Antigenically novel, high immune escape, high virulence | Yes | High | -3.66 | -3.65 | -3.79 | -3.70 | -3.61 | -3.61 | -3.73 | -3.67 | -3.53 | -3.54 | -3.57 | -3.57 | -3.56 | -5.29 | -5.40 | -5.57 | -5.53 | -5.24 | -5.36 | -5.51 | -5.48 | -5.22 | -5.32 | -5.35 | -5.38 | -5.46 |
| | Yes | Low | -3.87 | -3.89 | -4.02 | -3.94 | -3.82 | -3.85 | -3.97 | -3.92 | -3.75 | -3.79 | -3.82 | -3.81 | -3.83 | -5.43 | -5.59 | -5.74 | -5.73 | -5.38 | -5.57 | -5.67 | -5.69 | -5.39 | -5.54 | -5.54 | -5.61 | -5.72 |
| | No | High | -3.23 | -3.20 | -3.35 | -3.24 | -3.17 | -3.15 | -3.28 | -3.21 | -3.07 | -3.08 | -3.12 | -3.10 | -3.08 | -5.10 | -5.17 | -5.37 | -5.29 | -5.03 | -5.13 | -5.28 | -5.26 | -4.99 | -5.08 | -5.11 | -5.16 | -5.20 |
| | No | Low | -3.37 | -3.34 | -3.50 | -3.40 | -3.30 | -3.30 | -3.44 | -3.36 | -3.21 | -3.24 | -3.27 | -3.25 | -3.23 | -5.20 | -5.32 | -5.50 | -5.44 | -5.13 | -5.28 | -5.42 | -5.40 | -5.12 | -5.24 | -5.26 | -5.31 | -5.37 |

*OT: Omicron-targeted vaccine in Q4 2022; 2*OT: Omicron-targeted vaccines in Q4 2022 and Q2 2023; OT+M: Omicron-targeted vaccine in Q4 2022 and multivalent vaccine in Q2 2023; 30+: administered to people aged ≥30 years; 60+ administered to people aged ≥60 years; H: high coverage; L: low coverage*

### Supplementary Figure 10: Heat map of mean net monetary benefit from a health system plus GDP perspective (with a health-adjusted life year valued at AUD 70,000, 3% discount rate on both health-adjusted life years and costs; billions) for each of 936 scenarios over a 12 month timeline

| | | PHSMs | Higher Stringency | | | | | | | | | | | | | Lower Stringency | | | | | | | | | | | | |
|---|---|---|---|---|---|---|---|---|---|---|---|---|---|---|---|---|---|---|---|---|---|---|---|---|---|---|---|---|
| | | Vaccine schedule | OT+M | | | | 2*OT | | | | OT | | | | Nil | OT+M | | | | 2*OT | | | | OT | | | | Nil |
| | | Vaccine age group | 30+ | | 60+ | | 30+ | | 60+ | | 30+ | | 60+ | | | 30+ | | 60+ | | 30+ | | 60+ | | 30+ | | 60+ | | |
| | | Vaccine uptake | *H* | *L* | *H* | *L* | *H* | *L* | *H* | *L* | *H* | *L* | *H* | *L* | | *H* | *L* | *H* | *L* | *H* | *L* | *H* | *L* | *H* | *L* | *H* | *L* | |
| Variant | Respirator stockpile | Mask wearing | | | | | | | | | | | | | | | | | | | | | | | | | | |
| Omicron BA.4/5 only (no new variant) | Yes | High | -1.77 | -1.91 | -1.86 | -1.96 | -1.75 | -1.88 | -1.84 | -1.95 | -1.84 | -1.95 | -1.86 | -1.98 | -2.17 | -1.63 | -1.74 | -1.70 | -1.79 | -1.60 | -1.73 | -1.69 | -1.77 | -1.78 | -1.83 | -1.75 | -1.82 | -1.96 |
| | Yes | Low | -1.79 | -1.91 | -1.86 | -1.96 | -1.74 | -1.90 | -1.85 | -1.95 | -1.86 | -1.95 | -1.87 | -1.97 | -2.18 | -1.63 | -1.74 | -1.71 | -1.78 | -1.60 | -1.73 | -1.69 | -1.77 | -1.78 | -1.83 | -1.75 | -1.82 | -1.97 |
| | No | High | -1.74 | -1.85 | -1.80 | -1.89 | -1.70 | -1.84 | -1.80 | -1.89 | -1.80 | -1.90 | -1.81 | -1.91 | -2.12 | -1.63 | -1.73 | -1.70 | -1.77 | -1.59 | -1.72 | -1.68 | -1.76 | -1.77 | -1.82 | -1.74 | -1.81 | -1.95 |
| | No | Low | -1.74 | -1.84 | -1.82 | -1.92 | -1.71 | -1.82 | -1.80 | -1.89 | -1.81 | -1.88 | -1.81 | -1.92 | -2.10 | -1.63 | -1.73 | -1.70 | -1.78 | -1.59 | -1.72 | -1.68 | -1.76 | -1.78 | -1.81 | -1.75 | -1.81 | -1.94 |
| Antigenically Omicron-like, low immune escape, low virulence | Yes | High | -2.22 | -2.45 | -2.35 | -2.51 | -2.19 | -2.43 | -2.33 | -2.50 | -2.28 | -2.50 | -2.36 | -2.53 | -2.88 | -1.96 | -2.09 | -2.02 | -2.13 | -1.93 | -2.07 | -2.00 | -2.12 | -2.05 | -2.15 | -2.06 | -2.16 | -2.37 |
| | Yes | Low | -2.22 | -2.43 | -2.34 | -2.50 | -2.19 | -2.42 | -2.32 | -2.49 | -2.30 | -2.48 | -2.34 | -2.54 | -2.88 | -1.96 | -2.09 | -2.02 | -2.13 | -1.93 | -2.07 | -2.00 | -2.12 | -2.05 | -2.15 | -2.06 | -2.16 | -2.37 |
| | No | High | -2.15 | -2.38 | -2.27 | -2.42 | -2.12 | -2.35 | -2.26 | -2.41 | -2.20 | -2.44 | -2.27 | -2.44 | -2.76 | -1.95 | -2.07 | -2.01 | -2.11 | -1.92 | -2.06 | -1.99 | -2.10 | -2.04 | -2.14 | -2.04 | -2.15 | -2.35 |
| | No | Low | -2.15 | -2.36 | -2.27 | -2.42 | -2.12 | -2.34 | -2.26 | -2.40 | -2.20 | -2.42 | -2.29 | -2.42 | -2.77 | -1.95 | -2.08 | -2.01 | -2.11 | -1.92 | -2.06 | -1.99 | -2.10 | -2.04 | -2.14 | -2.04 | -2.15 | -2.35 |
| Antigenically novel, low immune escape, low virulence | Yes | High | -2.83 | -3.13 | -3.00 | -3.21 | -2.84 | -3.14 | -3.01 | -3.22 | -2.93 | -3.24 | -3.06 | -3.24 | -3.70 | -2.27 | -2.46 | -2.36 | -2.50 | -2.27 | -2.47 | -2.36 | -2.51 | -2.35 | -2.51 | -2.39 | -2.53 | -2.81 |
| | Yes | Low | -2.84 | -3.16 | -3.00 | -3.21 | -2.85 | -3.16 | -3.01 | -3.22 | -2.93 | -3.23 | -3.05 | -3.27 | -3.74 | -2.28 | -2.46 | -2.35 | -2.51 | -2.27 | -2.47 | -2.36 | -2.51 | -2.35 | -2.51 | -2.39 | -2.54 | -2.81 |
| | No | High | -2.74 | -2.99 | -2.88 | -3.10 | -2.74 | -3.01 | -2.89 | -3.09 | -2.84 | -3.09 | -2.92 | -3.12 | -3.56 | -2.25 | -2.43 | -2.33 | -2.48 | -2.25 | -2.44 | -2.34 | -2.48 | -2.33 | -2.48 | -2.36 | -2.51 | -2.76 |
| | No | Low | -2.75 | -3.04 | -2.93 | -3.14 | -2.76 | -3.05 | -2.93 | -3.13 | -2.84 | -3.11 | -2.96 | -3.17 | -3.60 | -2.25 | -2.43 | -2.33 | -2.48 | -2.25 | -2.44 | -2.33 | -2.48 | -2.33 | -2.48 | -2.36 | -2.50 | -2.77 |
| Antigenically Omicron-like, high immune escape, low virulence | Yes | High | -2.91 | -3.21 | -3.05 | -3.30 | -2.84 | -3.20 | -3.05 | -3.29 | -2.96 | -3.26 | -3.10 | -3.33 | -3.76 | -2.34 | -2.53 | -2.42 | -2.58 | -2.31 | -2.51 | -2.40 | -2.57 | -2.38 | -2.56 | -2.44 | -2.59 | -2.90 |
| | Yes | Low | -2.91 | -3.22 | -3.06 | -3.33 | -2.88 | -3.22 | -3.06 | -3.31 | -2.98 | -3.28 | -3.08 | -3.37 | -3.83 | -2.34 | -2.53 | -2.43 | -2.57 | -2.30 | -2.51 | -2.41 | -2.57 | -2.38 | -2.56 | -2.44 | -2.60 | -2.92 |
| | No | High | -2.80 | -3.12 | -2.96 | -3.19 | -2.77 | -3.10 | -2.95 | -3.17 | -2.86 | -3.16 | -2.98 | -3.22 | -3.65 | -2.31 | -2.49 | -2.40 | -2.55 | -2.28 | -2.47 | -2.38 | -2.53 | -2.35 | -2.53 | -2.42 | -2.56 | -2.86 |
| | No | Low | -2.84 | -3.11 | -2.98 | -3.18 | -2.80 | -3.10 | -2.96 | -3.18 | -2.89 | -3.18 | -2.98 | -3.24 | -3.67 | -2.32 | -2.49 | -2.40 | -2.54 | -2.29 | -2.47 | -2.38 | -2.53 | -2.36 | -2.53 | -2.41 | -2.56 | -2.86 |
| Antigenically novel, high immune escape, low virulence | Yes | High | -3.70 | -4.11 | -3.90 | -4.18 | -3.72 | -4.10 | -3.89 | -4.18 | -3.84 | -4.22 | -3.96 | -4.25 | -4.71 | -2.79 | -3.02 | -2.90 | -3.09 | -2.80 | -3.03 | -2.90 | -3.10 | -2.84 | -3.06 | -2.93 | -3.12 | -3.46 |
| | Yes | Low | -3.74 | -4.18 | -3.94 | -4.21 | -3.76 | -4.15 | -3.96 | -4.22 | -3.87 | -4.26 | -4.05 | -4.31 | -4.82 | -2.80 | -3.02 | -2.90 | -3.09 | -2.80 | -3.03 | -2.90 | -3.10 | -2.85 | -3.06 | -2.92 | -3.12 | -3.45 |
| | No | High | -3.61 | -3.95 | -3.79 | -4.06 | -3.62 | -3.95 | -3.80 | -4.04 | -3.72 | -4.02 | -3.84 | -4.10 | -4.58 | -2.75 | -2.97 | -2.83 | -3.03 | -2.76 | -2.97 | -2.84 | -3.03 | -2.80 | -3.00 | -2.86 | -3.06 | -3.39 |
| | No | Low | -3.64 | -4.04 | -3.80 | -4.05 | -3.65 | -4.06 | -3.82 | -4.04 | -3.78 | -4.15 | -3.88 | -4.12 | -4.66 | -2.77 | -2.99 | -2.85 | -3.04 | -2.76 | -2.99 | -2.85 | -3.04 | -2.80 | -3.02 | -2.88 | -3.08 | -3.42 |
| Antigenically Omicron-like, low immune escape, high virulence | Yes | High | -5.33 | -5.91 | -5.78 | -6.12 | -5.30 | -5.90 | -5.76 | -6.13 | -5.69 | -6.11 | -5.92 | -6.19 | -6.65 | -4.66 | -5.20 | -5.09 | -5.40 | -4.59 | -5.16 | -5.08 | -5.39 | -4.93 | -5.36 | -5.14 | -5.45 | -6.03 |
| | Yes | Low | -5.55 | -6.23 | -6.05 | -6.52 | -5.52 | -6.22 | -6.06 | -6.50 | -6.03 | -6.50 | -6.27 | -6.69 | -7.20 | -4.67 | -5.23 | -5.11 | -5.47 | -4.63 | -5.16 | -5.12 | -5.46 | -4.99 | -5.42 | -5.19 | -5.54 | -6.19 |
| | No | High | -5.25 | -5.86 | -5.70 | -6.13 | -5.20 | -5.83 | -5.68 | -6.08 | -5.66 | -6.12 | -5.89 | -6.21 | -6.71 | -4.57 | -5.08 | -5.03 | -5.31 | -4.54 | -5.08 | -5.02 | -5.32 | -4.85 | -5.28 | -5.09 | -5.37 | -5.96 |
| | No | Low | -5.45 | -6.14 | -5.97 | -6.41 | -5.43 | -6.13 | -5.98 | -6.37 | -5.96 | -6.46 | -6.22 | -6.59 | -7.17 | -4.61 | -5.17 | -5.04 | -5.38 | -4.56 | -5.13 | -5.05 | -5.38 | -4.94 | -5.37 | -5.12 | -5.47 | -6.13 |
| Antigenically novel, low immune escape, high virulence | Yes | High | -7.06 | -7.63 | -7.52 | -7.83 | -7.10 | -7.65 | -7.54 | -7.84 | -7.47 | -7.84 | -7.67 | -7.93 | -8.33 | -6.03 | -6.67 | -6.54 | -6.92 | -6.07 | -6.68 | -6.54 | -6.92 | -6.41 | -6.90 | -6.67 | -7.00 | -7.66 |
| | Yes | Low | -7.49 | -8.17 | -8.06 | -8.40 | -7.63 | -8.24 | -8.10 | -8.41 | -8.06 | -8.48 | -8.30 | -8.60 | -9.11 | -6.11 | -6.89 | -6.63 | -7.11 | -6.15 | -6.88 | -6.65 | -7.12 | -6.56 | -7.14 | -6.83 | -7.25 | -7.94 |
| | No | High | -6.96 | -7.63 | -7.46 | -7.88 | -7.08 | -7.66 | -7.50 | -7.92 | -7.51 | -7.86 | -7.71 | -8.03 | -8.47 | -5.95 | -6.60 | -6.41 | -6.85 | -5.98 | -6.62 | -6.42 | -6.82 | -6.34 | -6.84 | -6.56 | -6.94 | -7.61 |
| | No | Low | -7.46 | -8.16 | -8.00 | -8.38 | -7.57 | -8.23 | -8.05 | -8.45 | -8.01 | -8.46 | -8.24 | -8.57 | -9.14 | -6.03 | -6.73 | -6.52 | -6.95 | -6.04 | -6.77 | -6.56 | -6.99 | -6.45 | -7.05 | -6.71 | -7.13 | -7.93 |
| Antigenically Omicron-like, high immune escape, high virulence | Yes | High | -7.23 | -7.77 | -7.65 | -8.01 | -7.20 | -7.76 | -7.68 | -7.99 | -7.54 | -7.98 | -7.77 | -8.07 | -8.49 | -6.19 | -6.84 | -6.69 | -7.08 | -6.16 | -6.81 | -6.68 | -7.07 | -6.49 | -7.05 | -6.76 | -7.17 | -7.85 |
| | Yes | Low | -7.77 | -8.37 | -8.21 | -8.58 | -7.72 | -8.34 | -8.17 | -8.58 | -8.16 | -8.61 | -8.37 | -8.71 | -9.30 | -6.25 | -6.99 | -6.82 | -7.26 | -6.25 | -7.00 | -6.80 | -7.22 | -6.65 | -7.25 | -6.98 | -7.41 | -8.16 |
| | No | High | -7.21 | -7.83 | -7.72 | -8.03 | -7.20 | -7.81 | -7.70 | -8.01 | -7.60 | -8.02 | -7.84 | -8.18 | -8.61 | -6.12 | -6.76 | -6.59 | -7.01 | -6.06 | -6.71 | -6.56 | -6.99 | -6.45 | -6.95 | -6.69 | -7.10 | -7.86 |
| | No | Low | -7.67 | -8.32 | -8.18 | -8.66 | -7.61 | -8.33 | -8.17 | -8.67 | -8.09 | -8.58 | -8.37 | -8.78 | -9.37 | -6.19 | -6.90 | -6.67 | -7.18 | -6.11 | -6.88 | -6.70 | -7.18 | -6.56 | -7.15 | -6.85 | -7.29 | -8.15 |
| Antigenically novel, high immune escape, high virulence | Yes | High | -8.98 | -9.44 | -9.37 | -9.59 | -9.04 | -9.46 | -9.36 | -9.63 | -9.33 | -9.59 | -9.50 | -9.68 | -10.12 | -7.88 | -8.58 | -8.39 | -8.81 | -7.95 | -8.56 | -8.43 | -8.80 | -8.32 | -8.78 | -8.55 | -8.93 | -9.49 |
| | Yes | Low | -9.73 | -10.35 | -10.12 | -10.55 | -9.83 | -10.38 | -10.20 | -10.60 | -10.19 | -10.57 | -10.38 | -10.66 | -11.17 | -8.12 | -8.86 | -8.64 | -9.20 | -8.19 | -8.91 | -8.65 | -9.18 | -8.62 | -9.20 | -8.84 | -9.33 | -10.06 |
| | No | High | -9.07 | -9.64 | -9.47 | -9.83 | -9.13 | -9.67 | -9.52 | -9.81 | -9.49 | -9.85 | -9.68 | -9.93 | -10.38 | -7.83 | -8.52 | -8.33 | -8.79 | -7.88 | -8.56 | -8.34 | -8.82 | -8.27 | -8.77 | -8.51 | -8.93 | -9.59 |
| | No | Low | -9.78 | -10.41 | -10.23 | -10.67 | -9.92 | -10.48 | -10.27 | -10.68 | -10.31 | -10.72 | -10.47 | -10.82 | -11.31 | -8.01 | -8.81 | -8.56 | -9.10 | -8.08 | -8.83 | -8.63 | -9.15 | -8.52 | -9.12 | -8.82 | -9.30 | -10.04 |

*OT: Omicron-targeted vaccine in Q4 2022; 2*OT: Omicron-targeted vaccines in Q4 2022 and Q2 2023; OT+M: Omicron-targeted vaccine in Q4 2022 and multivalent vaccine in Q2 2023; 30+: administered to people aged ≥30 years; 60+ administered to people aged ≥60 years; H: high coverage; L: low coverage*

**Supplementary Figure 11: Mean time spent in stages 2, 3 and 4 or higher over 12 months for packages of policy options and nine future SARS-CoV-2 variant scenarios, averaged across mask policies**

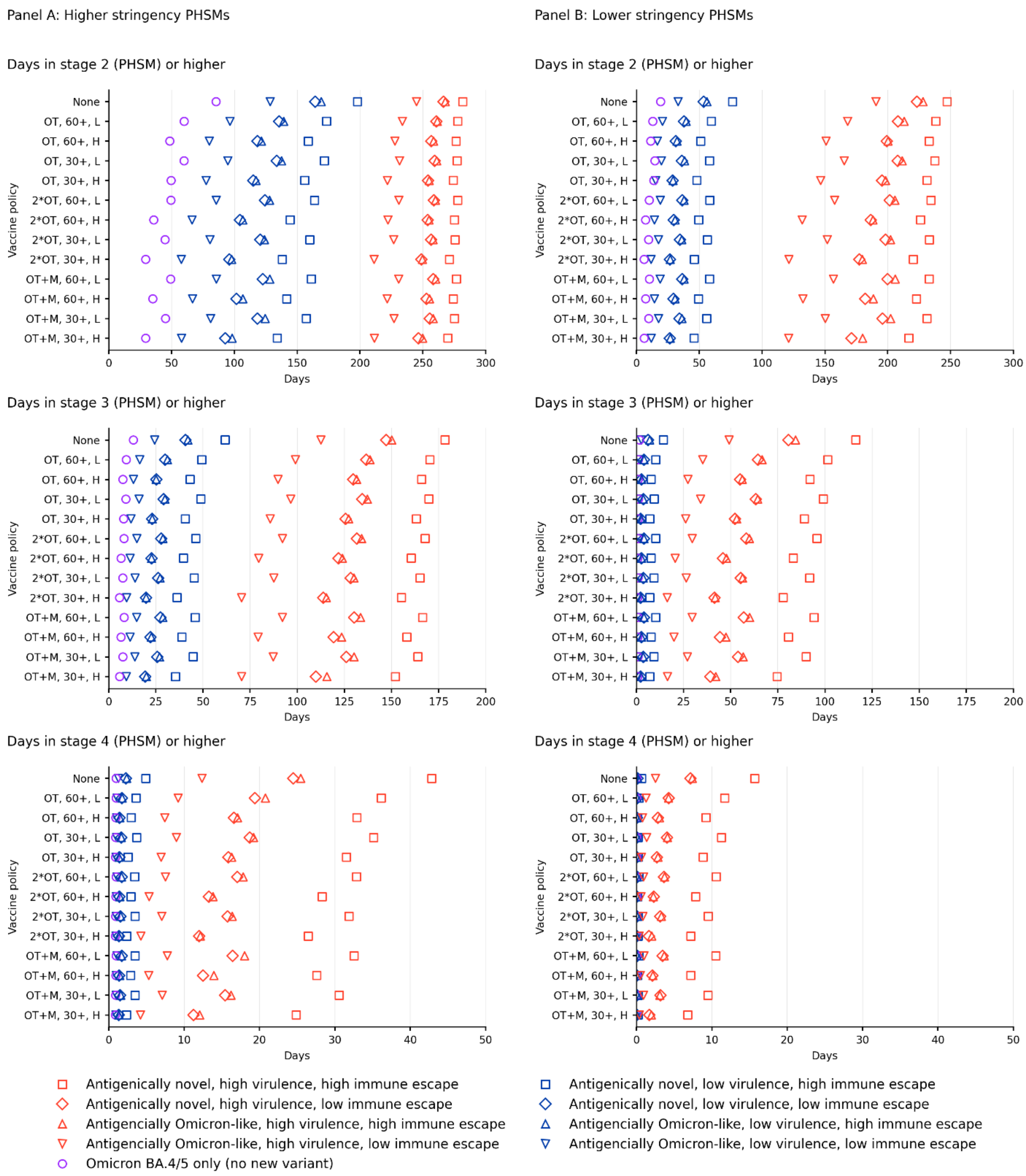

*OT: Omicron-targeted vaccine in Q4 2022; 2*OT: Omicron-targeted vaccines in Q4 2022 and Q2 2023; OT+M: Omicron-targeted vaccine in Q4 2022 and multivalent vaccine in Q2 2023; 30+: administered to people aged ≥30 years; 60+ administered to people aged ≥60 years; H: high coverage; L: low coverage*

**Supplementary Figure 12: Mean days spent with >750 and >1500 COVID-19 patients admitted to hospital per million population over 12 months for packages of policy options and nine future SARS-CoV-2 variant scenarios, averaged across mask policies**

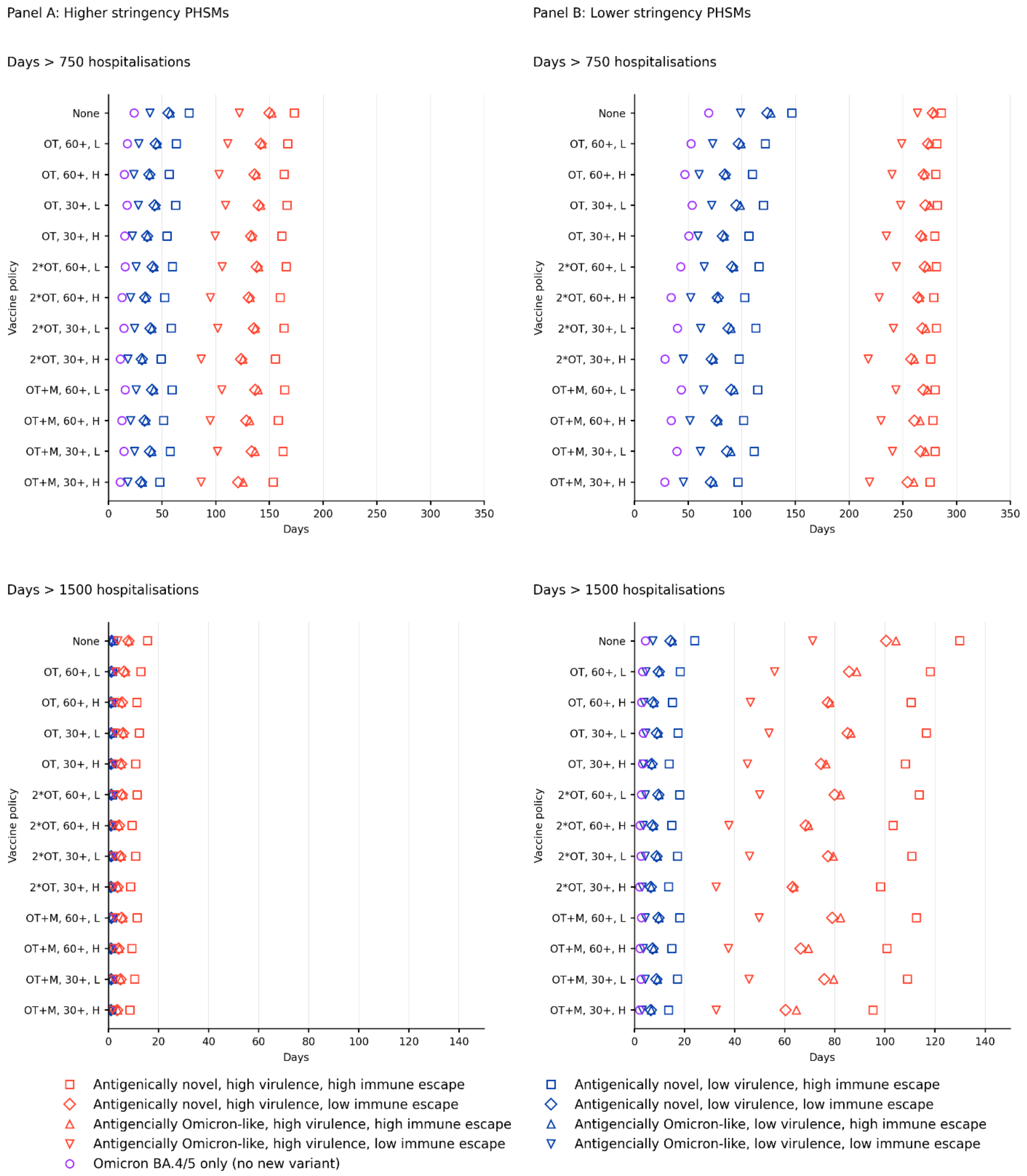

*OT: Omicron-targeted vaccine in Q4 2022; 2*OT: Omicron-targeted vaccines in Q4 2022 and Q2 2023; OT+M: Omicron-targeted vaccine in Q4 2022 and multivalent vaccine in Q2 2023; 30+: administered to people aged ≥30 years; 60+ administered to people aged ≥60 years; H: high coverage; L: low coverage*

### Supplementary Figure 13: Mean net health expenditure (due to and excluding deaths incurred) over 12 months for packages of policy options and nine future SARS-CoV-2 variant scenarios, averaged across mask policies

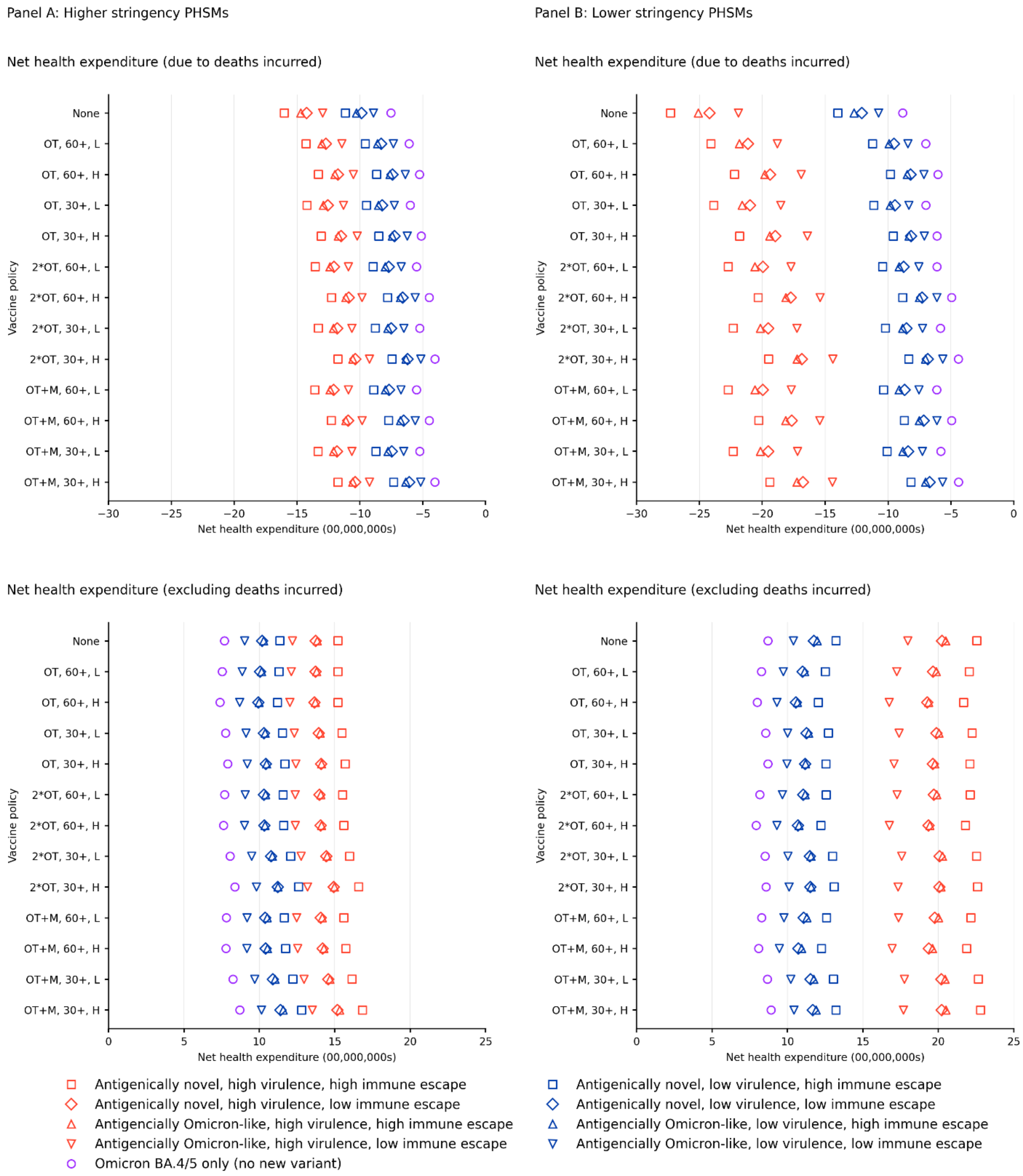

*OT: Omicron-targeted vaccine in Q4 2022; 2*OT: Omicron-targeted vaccines in Q4 2022 and Q2 2023; OT+M: Omicron-targeted vaccine in Q4 2022 and multivalent vaccine in Q2 2023; 30+: administered to people aged ≥30 years; 60+ administered to people aged ≥60 years; H: high coverage; L: low coverage*

**Supplementary Figure 14: Mean net monetary benefit for a willingness to pay per health-adjusted life year of AUD 70,000 (USD 50,000) from health system and health system plus GDP perspectives, over 12 months for packages of policy options and nine future SARS-CoV-2 variant scenarios, averaged across mask policies.**

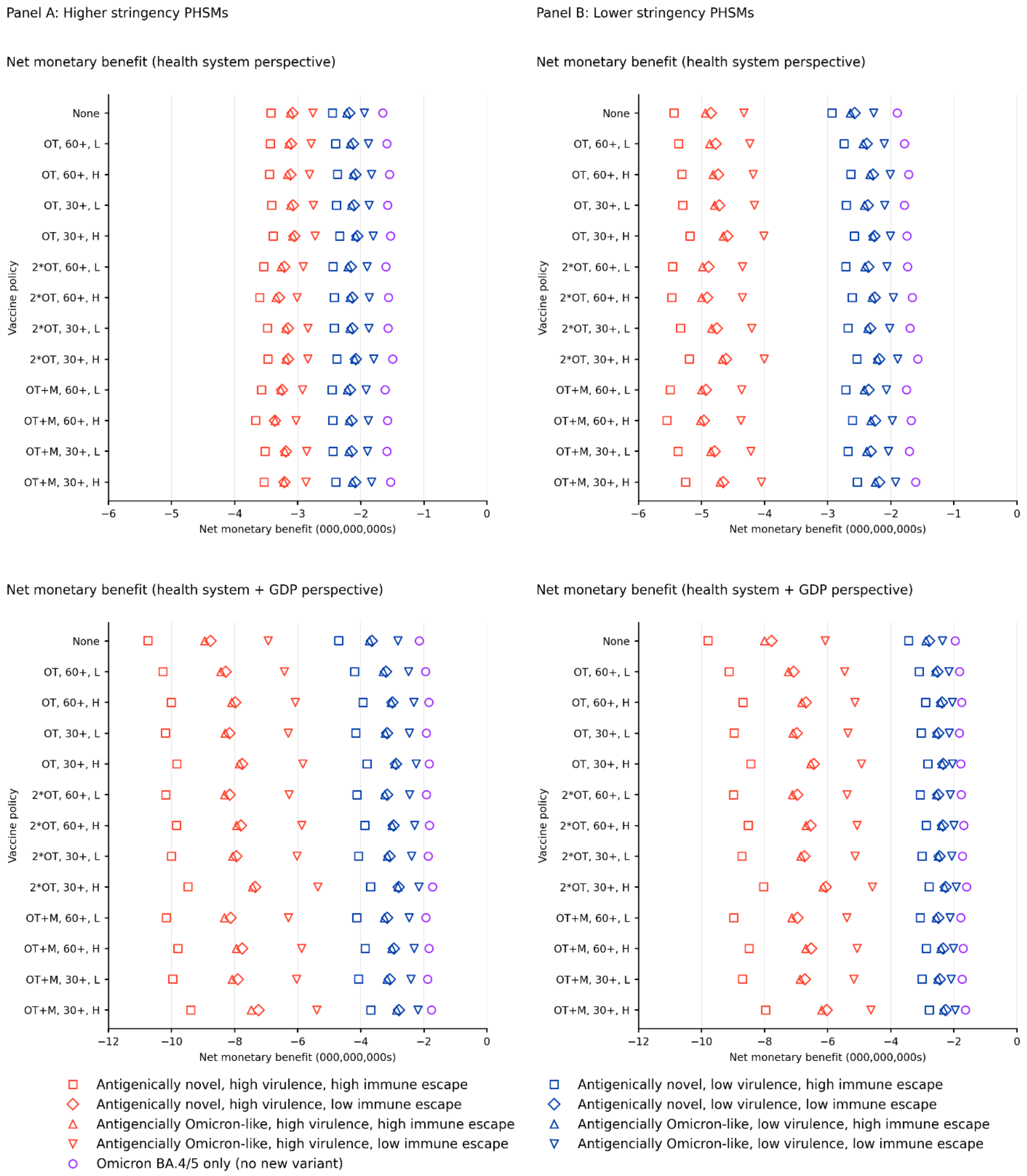

*OT: Omicron-targeted vaccine in Q4 2022; 2*OT: Omicron-targeted vaccines in Q4 2022 and Q2 2023; OT+M: Omicron-targeted vaccine in Q4 2022 and multivalent vaccine in Q2 2023; 30+: administered to people aged ≥30 years; 60+ administered to people aged ≥60 years; H: high coverage; L: low coverage*

### Supplementary Figure 15: Mean cumulative infections, hospitalisations and deaths over 12 months for packages of policy options and nine future SARS-CoV-2 scenarios, with no or both mask policies active (averaged over the two PHSM policies)

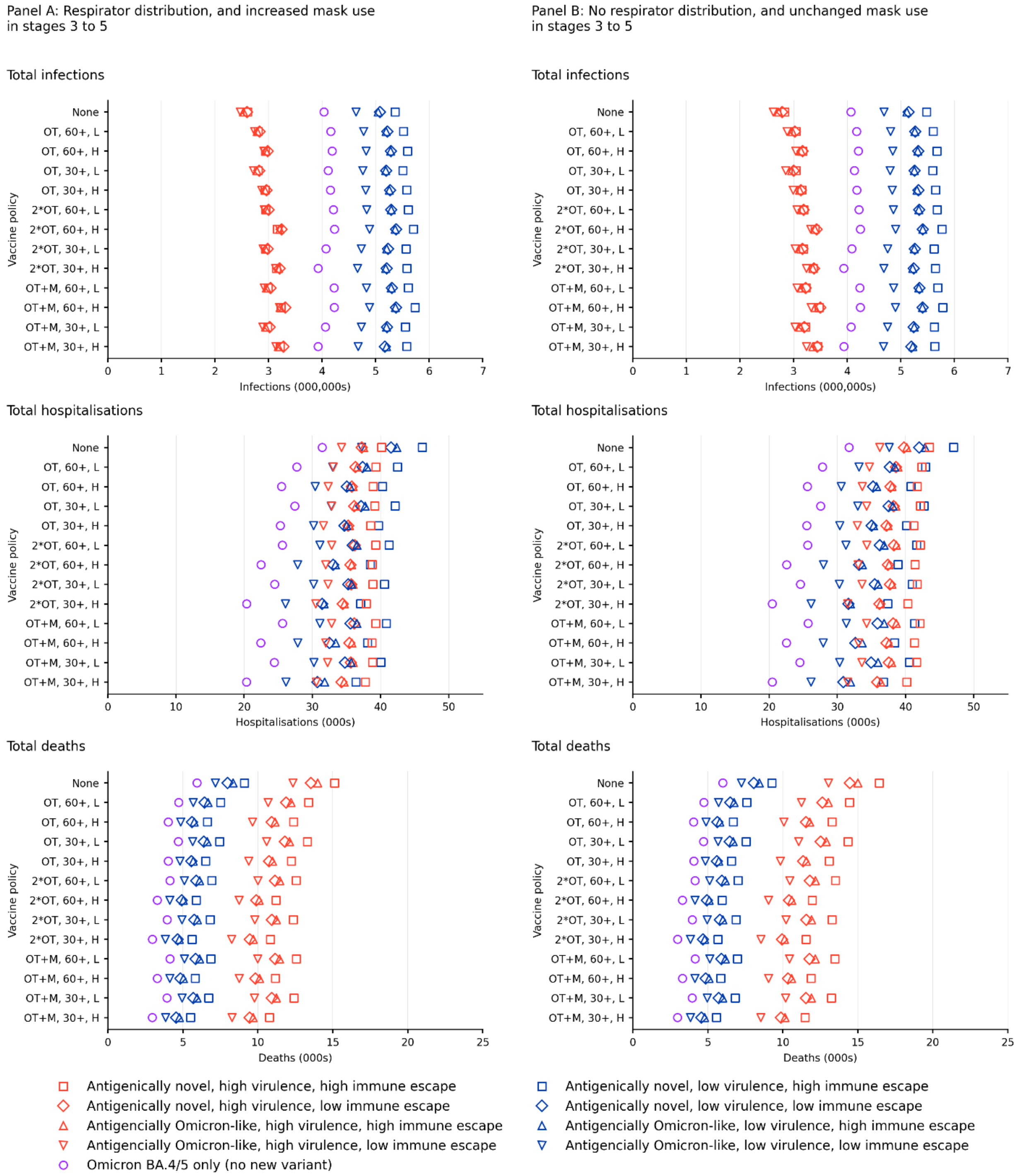

*OT: Omicron-targeted vaccine in Q4 2022; 2*OT: Omicron-targeted vaccines in Q4 2022 and Q2 2023; OT+M: Omicron-targeted vaccine in Q4 2022 and multivalent vaccine in Q2 2023; 30+: administered to people aged ≥30 years; 60+ administered to people aged ≥60 years; H: high coverage; L: low coverage*

**Supplementary Figure 16: Mean days spent with >750 and >1500 COVID-19 patients admitted to hospital per million population over 12 months for packages of policy options and nine future SARS-CoV-2 variant scenarios, with no or both mask policies active (averaged over the two PHSM policies)**

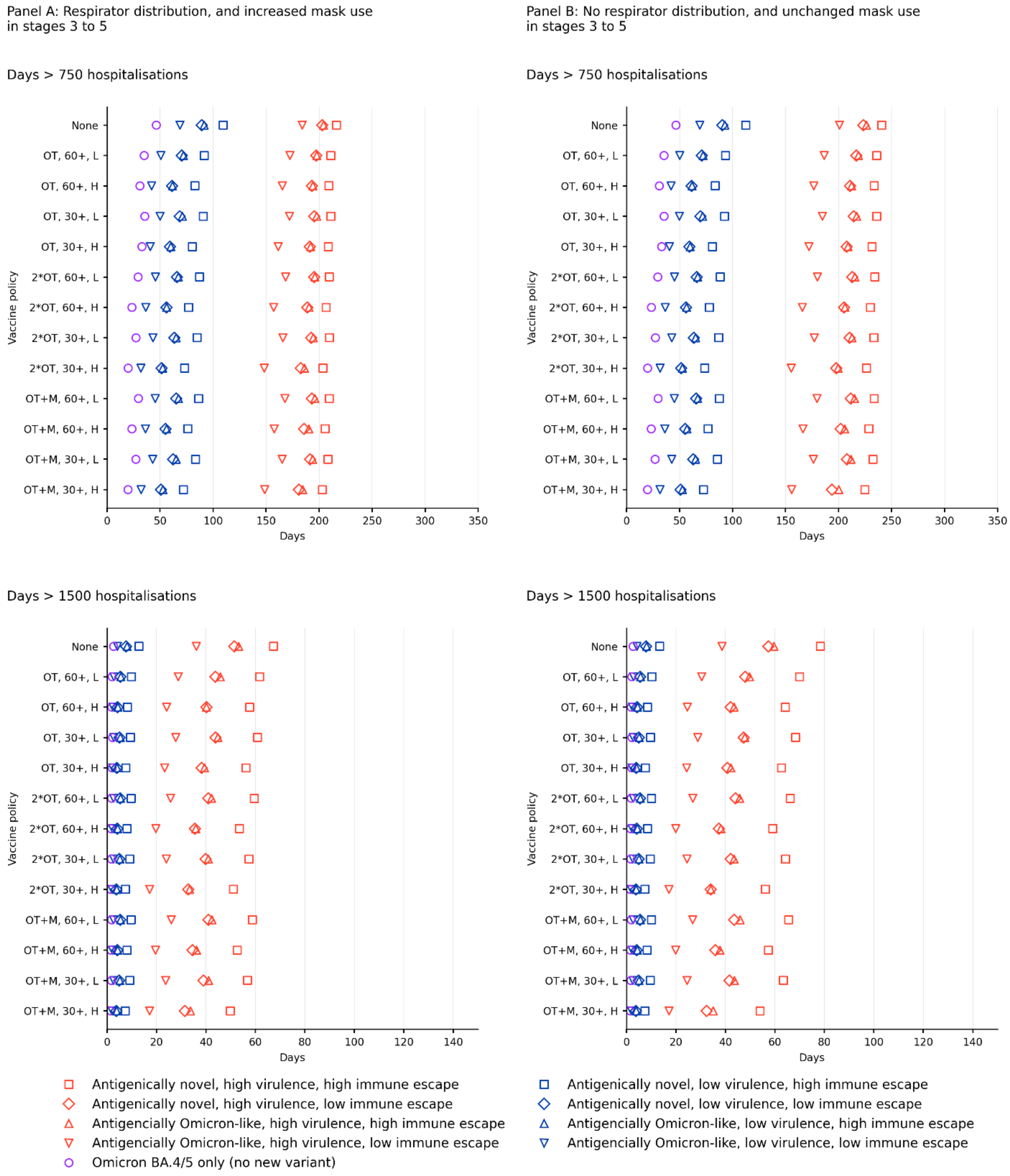

*OT: Omicron-targeted vaccine in Q4 2022; 2*OT: Omicron-targeted vaccines in Q4 2022 and Q2 2023; OT+M: Omicron-targeted vaccine in Q4 2022 and multivalent vaccine in Q2 2023; 30+: administered to people aged ≥30 years; 60+ administered to people aged ≥60 years; H: high coverage; L: low coverage*

### Supplementary Figure 17: Incremental cost effectiveness ratios for head-to-head comparison of any ongoing vaccination schedule incremental to no further vaccination (health system perspective)

| | | PHSMs | Higher Stringency | | | | | | | | | | | | | Lower Stringency | | | | | | | | | | | | |
|---|---|---|---|---|---|---|---|---|---|---|---|---|---|---|---|---|---|---|---|---|---|---|---|---|---|---|---|---|
| | | Vaccine schedule | OT+M | | | | 2*OT | | | | OT | | | | Nil | OT+M | | | | 2*OT | | | | OT | | | | Nil |
| | | Vaccine age group | 30+ | | 60+ | | 30+ | | 60+ | | 30+ | | 60+ | | | 30+ | | 60+ | | 30+ | | 60+ | | 30+ | | 60+ | | |
| | | Vaccine uptake | H | L | H | L | H | L | H | L | H | L | H | L | | H | L | H | L | H | L | H | L | H | L | H | L | |
| Variant | Respirator stockpile | Mask wearing | | | | | | | | | | | | | | | | | | | | | | | | | | |
| Omicron BA.4/5 only (no new variant) | Yes | High | 53365 | 55813 | 54607 | 58528 | 49640 | 51769 | 52042 | 55192 | 45671 | 45422 | 43137 | 45030 | | 42523 | 42125 | 41141 | 42305 | 39348 | 40087 | 39105 | 39543 | 44140 | 41744 | 36937 | 37987 | |
| | Yes | Low | 52845 | 54709 | 54023 | 56120 | 48678 | 50886 | 51258 | 53962 | 46368 | 45798 | 43288 | 43320 | | 42462 | 41738 | 41101 | 41391 | 39300 | 39810 | 38878 | 39015 | 44049 | 41133 | 36670 | 37104 | |
| | No | High | 57061 | 59111 | 58789 | 62440 | 52954 | 54992 | 55991 | 58758 | 49197 | 49458 | 47952 | 50090 | | 43609 | 43427 | 42661 | 43557 | 40453 | 41659 | 40366 | 41177 | 45487 | 42800 | 38291 | 39689 | |
| | No | Low | 56280 | 57882 | 58254 | 61968 | 52493 | 54868 | 55430 | 57493 | 49338 | 48927 | 47368 | 48104 | | 43662 | 43401 | 42403 | 43602 | 40410 | 41303 | 40303 | 40905 | 45721 | 42394 | 38421 | 39568 | |
| Antigenically Omicron-like, low immune escape, low virulence | Yes | High | 54179 | 57883 | 56219 | 58395 | 49801 | 53014 | 53463 | 55158 | 43517 | 44449 | 42084 | 43503 | | 41211 | 40380 | 38344 | 38591 | 38439 | 38058 | 36520 | 37115 | 37476 | 36653 | 33290 | 33839 | |
| | Yes | Low | 53028 | 56116 | 54964 | 58810 | 49434 | 51853 | 52730 | 54306 | 42816 | 44345 | 41414 | 43279 | | 41160 | 40616 | 38168 | 38481 | 38592 | 38144 | 36353 | 37032 | 37770 | 36861 | 33418 | 33972 | |
| | No | High | 60888 | 64582 | 63875 | 67534 | 56727 | 60983 | 61257 | 63874 | 51498 | 54146 | 51657 | 54519 | | 42300 | 41295 | 39666 | 39783 | 39656 | 39340 | 37853 | 38418 | 38946 | 38168 | 34897 | 36028 | |
| | No | Low | 58721 | 62493 | 61599 | 65175 | 54854 | 58278 | 58866 | 61411 | 49932 | 52209 | 49401 | 51065 | | 42286 | 41653 | 39520 | 40321 | 39845 | 39283 | 37927 | 38442 | 39174 | 38576 | 34962 | 35758 | |
| Antigenically novel, low immune escape, low virulence | Yes | High | 56106 | 59967 | 60004 | 62038 | 53986 | 56828 | 57830 | 58590 | 43818 | 46035 | 44141 | 44425 | | 39394 | 39622 | 37253 | 37875 | 38403 | 39288 | 36853 | 37424 | 34717 | 34079 | 32345 | 32905 | |
| | Yes | Low | 53808 | 56773 | 56220 | 59240 | 52513 | 54865 | 55304 | 55277 | 42321 | 43222 | 41147 | 42112 | | 39419 | 39101 | 37145 | 37762 | 38207 | 38918 | 36750 | 37409 | 34632 | 34035 | 32107 | 32833 | |
| | No | High | 63450 | 67086 | 70460 | 75143 | 62332 | 65539 | 67600 | 70420 | 54219 | 55589 | 55312 | 56428 | | 41587 | 41321 | 39329 | 39637 | 40563 | 41064 | 39226 | 39406 | 37742 | 36925 | 35190 | 35576 | |
| | No | Low | 62075 | 65328 | 67511 | 70815 | 60727 | 63363 | 65177 | 68577 | 53596 | 54160 | 53053 | 54345 | | 41438 | 41229 | 39109 | 39588 | 40444 | 40710 | 38710 | 38970 | 37647 | 36975 | 34892 | 35030 | |
| Antigenically Omicron-like, high immune escape, low virulence | Yes | High | 56492 | 59369 | 59568 | 61987 | 51843 | 55990 | 56473 | 59393 | 43248 | 44158 | 43860 | 44186 | | 40069 | 40029 | 37971 | 38660 | 37544 | 37963 | 36303 | 37276 | 33588 | 34116 | 32471 | 32689 | |
| | Yes | Low | 54801 | 57571 | 57110 | 61093 | 50634 | 54440 | 54603 | 57712 | 41945 | 43832 | 41282 | 44090 | | 39590 | 39420 | 37659 | 38056 | 37153 | 37211 | 36117 | 36780 | 33242 | 33495 | 32055 | 32361 | |
| | No | High | 65386 | 69734 | 70399 | 76659 | 61459 | 66391 | 67836 | 72627 | 54735 | 56448 | 56459 | 59659 | | 42137 | 42053 | 40627 | 41356 | 39956 | 39899 | 38775 | 39528 | 36741 | 36971 | 35697 | 36049 | |
| | No | Low | 62944 | 67151 | 67386 | 70388 | 59141 | 63372 | 65253 | 68475 | 52371 | 54624 | 53480 | 55298 | | 42290 | 42032 | 40614 | 41070 | 39884 | 40082 | 38863 | 39377 | 36711 | 37288 | 35327 | 36164 | |
| Antigenically novel, high immune escape, low virulence | Yes | High | 60027 | 63320 | 65152 | 66469 | 57166 | 60122 | 60005 | 61971 | 45500 | 47217 | 45590 | 47291 | | 40823 | 40946 | 39496 | 40058 | 39947 | 40538 | 38615 | 40143 | 35024 | 35236 | 34189 | 35707 | |
| | Yes | Low | 56746 | 60934 | 60849 | 62328 | 54985 | 57164 | 57034 | 59821 | 43747 | 44324 | 43663 | 45691 | | 40437 | 40087 | 38786 | 39269 | 39571 | 39421 | 38330 | 38964 | 34560 | 34225 | 33614 | 34905 | |
| | No | High | 70089 | 74262 | 76746 | 80446 | 67367 | 70494 | 72168 | 75624 | 57040 | 58947 | 58762 | 60432 | | 42536 | 42666 | 41175 | 41652 | 41996 | 41887 | 40859 | 40730 | 37497 | 37432 | 36512 | 37637 | |
| | No | Low | 66596 | 70132 | 72007 | 74943 | 63996 | 66581 | 68664 | 70827 | 55418 | 56409 | 56870 | 56240 | | 42489 | 42491 | 41009 | 40670 | 41473 | 41522 | 40452 | 40138 | 36856 | 37245 | 36585 | 36735 | |
| Antigenically Omicron-like, low immune escape, high virulence | Yes | High | 87812 | 104336 | 393672 | 291684 | 81241 | 99748 | 428643 | 244543 | 59418 | 64840 | 88502 | 85700 | | 49548 | 55232 | 79163 | 78880 | 46037 | 53336 | 75634 | 77228 | 39589 | 43449 | 48385 | 48905 | |
| | Yes | Low | 73190 | 83853 | 234959 | 201746 | 68118 | 77511 | 237983 | 185282 | 50433 | 52561 | 73858 | 68701 | | 44927 | 49770 | 66293 | 66361 | 42552 | 46656 | 64529 | 62526 | 36710 | 37818 | 44215 | 43140 | |
| | No | High | 104763 | 118584 | 392295 | 267630 | 98132 | 108279 | 349404 | 258514 | 73288 | 81824 | 113722 | 93911 | | 55421 | 60980 | 83667 | 86519 | 52341 | 60010 | 79730 | 84839 | 45501 | 47620 | 55954 | 55034 | |
| | No | Low | 92344 | 105386 | 242733 | 212509 | 88685 | 99060 | 227173 | 214600 | 67065 | 70542 | 91301 | 96334 | | 51868 | 58276 | 77584 | 79361 | 49395 | 56450 | 73976 | 76301 | 43369 | 46169 | 52737 | 53063 | |
| Antigenically novel, low immune escape, high virulence | Yes | High | 93317 | 107656 | 341991 | 283404 | 79987 | 90342 | 182298 | 169785 | 60723 | 67834 | 78868 | 81979 | | 53837 | 61432 | 89272 | 89479 | 50701 | 56538 | 78732 | 77284 | 42204 | 45704 | 50949 | 48977 | |
| | Yes | Low | 78830 | 92601 | 263933 | 180372 | 71276 | 81834 | 150074 | 135036 | 55790 | 60242 | 73364 | 77272 | | 49064 | 55197 | 73664 | 77016 | 46238 | 51373 | 66589 | 67112 | 39689 | 41859 | 47236 | 47322 | |
| | No | High | 103539 | 107579 | 273915 | 220353 | 90360 | 98375 | 182188 | 156449 | 72216 | 72987 | 87300 | 86465 | | 60729 | 67401 | 97847 | 100101 | 57455 | 63080 | 87181 | 86486 | 49465 | 52826 | 59936 | 59392 | |
| | No | Low | 94653 | 107196 | 244353 | 201248 | 84395 | 94111 | 161532 | 148995 | 66855 | 68374 | 85161 | 85184 | | 57723 | 63818 | 88179 | 88795 | 55125 | 60548 | 80935 | 79757 | 47599 | 50582 | 56873 | 58582 | |
| Antigenically Omicron-like, high immune escape, high virulence | Yes | High | 83175 | 93254 | 180015 | 173055 | 76726 | 87793 | 172008 | 170107 | 58531 | 65331 | 83496 | 77781 | | 52418 | 58644 | 82944 | 82647 | 50485 | 56289 | 79641 | 78508 | 41802 | 45331 | 50943 | 53179 | |
| | Yes | Low | 74562 | 80979 | 150504 | 135496 | 68887 | 76027 | 145458 | 134089 | 53139 | 57027 | 69389 | 67666 | | 47671 | 52828 | 69639 | 72673 | 45443 | 51257 | 66566 | 69376 | 39528 | 41067 | 47005 | 49294 | |
| | No | High | 95290 | 105111 | 207656 | 178002 | 90618 | 104125 | 193465 | 177405 | 71741 | 79477 | 93563 | 95621 | | 59747 | 66888 | 89141 | 92795 | 57064 | 64266 | 87182 | 88221 | 49897 | 53303 | 61228 | 62474 | |
| | No | Low | 88573 | 95721 | 173933 | 168121 | 84514 | 92497 | 166074 | 159441 | 66966 | 71741 | 89307 | 87756 | | 55835 | 60309 | 79095 | 80947 | 53467 | 59212 | 78162 | 80165 | 46885 | 49710 | 57012 | 57233 | |
| Antigenically novel, high immune escape, high virulence | Yes | High | 86621 | 95887 | 176494 | 163190 | 77387 | 82894 | 127568 | 124755 | 63743 | 65168 | 74143 | 73429 | | 56815 | 62519 | 88050 | 86144 | 53217 | 57177 | 76727 | 74114 | 45719 | 46989 | 53637 | 52788 | |
| | Yes | Low | 76291 | 85629 | 143973 | 127294 | 69472 | 76371 | 111149 | 106810 | 56164 | 58822 | 67939 | 63034 | | 50184 | 54917 | 73851 | 71916 | 47558 | 52472 | 65022 | 66005 | 40573 | 42944 | 47730 | 48854 | |
| | No | High | 93052 | 101609 | 167727 | 148721 | 82034 | 87551 | 128290 | 126636 | 68037 | 71504 | 81375 | 78048 | | 62678 | 66397 | 93108 | 86739 | 58286 | 61854 | 79668 | 80402 | 50941 | 51711 | 58430 | 61443 | |
| | No | Low | 87433 | 95971 | 157186 | 147892 | 77839 | 85065 | 123629 | 120436 | 65918 | 70638 | 80081 | 76978 | | 59033 | 64585 | 85794 | 83916 | 54902 | 60513 | 76221 | 75102 | 48828 | 51712 | 57021 | 58473 | |

*OT: Omicron-targeted vaccine in Q4 2022; 2*OT: Omicron-targeted vaccines in Q4 2022 and Q2 2023; OT+M: Omicron-targeted vaccine in Q4 2022 and multivalent vaccine in Q2 2023; 30+: administered to people aged ≥30 years; 60+ administered to people aged ≥60 years; H: high coverage; L: low coverage*

*NB: the missing column for 'nil vaccination' is the comparator; the left panel of ICERs are all compared to nil vaccination (in the same row) under more stringent PHSMs; the right panel of ICERs are all compared to nil vaccination (in the same row, final empty column) under less stringent PHSMs.*

### Supplementary Figure 18: Incremental cost effectiveness ratios for head-to-head comparison of any ongoing vaccination schedule incremental to no further vaccination (health system and GDP perspective)

| | | PHSMs | Higher Stringency | | | | | | | | | | | | | Lower Stringency | | | | | | | | | | | | |
|---|---|---|---|---|---|---|---|---|---|---|---|---|---|---|---|---|---|---|---|---|---|---|---|---|---|---|---|---|
| | | Vaccine schedule | OT+M | | | | 2*OT | | | | OT | | | | Nil | OT+M | | | | 2*OT | | | | OT | | | | Nil |
| | | Vaccine age group | 30+ | | 60+ | | 30+ | | 60+ | | 30+ | | 60+ | | | 30+ | | 60+ | | 30+ | | 60+ | | 30+ | | 60+ | | |
| | | Vaccine uptake | *H* | *L* | *H* | *L* | *H* | *L* | *H* | *L* | *H* | *L* | *H* | *L* | | *H* | *L* | *H* | *L* | *H* | *L* | *H* | *L* | *H* | *L* | *H* | *L* | |
| Variant | Respirator stockpile | Mask wearing | | | | | | | | | | | | | | | | | | | | | | | | | | |
| Omicron BA.4/5 only (no new variant) | Yes | High | 20423 | 16493 | 12210 | 8628 | 17307 | 10921 | 8915 | 4645 | 10101 | 2876 | dominant | dominant | | 39425 | 38141 | 36915 | 36693 | 36251 | 36124 | 34902 | 34007 | 39803 | 36885 | 32580 | 31343 | |
| | Yes | Low | 22513 | 17814 | 13919 | 12464 | 17534 | 14351 | 10808 | 6619 | 12448 | 4535 | dominant | dominant | | 39389 | 37995 | 37057 | 36025 | 36241 | 36030 | 34833 | 33743 | 40129 | 36479 | 32360 | 30858 | |
| | No | High | 23748 | 17978 | 14428 | 8878 | 19261 | 14605 | 12495 | 7220 | 13225 | 5559 | dominant | dominant | | 40211 | 39383 | 38273 | 37721 | 37056 | 37621 | 36026 | 35427 | 41161 | 38180 | 33772 | 32663 | |
| | No | Low | 26188 | 19247 | 19972 | 19494 | 22214 | 15366 | 17169 | 14323 | 16762 | 6367 | 4720 | 7156 | | 40526 | 39602 | 38455 | 38494 | 37305 | 37406 | 36424 | 35959 | 41981 | 37713 | 34431 | 33776 | |
| Antigenically Omicron-like, low immune escape, low virulence | Yes | High | dominant | dominant | dominant | dominant | dominant | dominant | dominant | dominant | dominant | dominant | dominant | dominant | | 36673 | 35445 | 33106 | 32633 | 33848 | 33145 | 31292 | 31215 | 31517 | 29486 | 26924 | 26353 | |
| | Yes | Low | dominant | dominant | dominant | dominant | dominant | dominant | dominant | dominant | dominant | dominant | dominant | dominant | | 36888 | 35661 | 33037 | 32898 | 34264 | 33185 | 31303 | 31509 | 32033 | 29854 | 27091 | 26770 | |
| | No | High | dominant | dominant | dominant | dominant | dominant | dominant | dominant | dominant | dominant | dominant | dominant | dominant | | 37745 | 36300 | 34374 | 33629 | 34997 | 34335 | 32582 | 32314 | 32837 | 31147 | 28340 | 27858 | |
| | No | Low | dominant | dominant | dominant | dominant | dominant | dominant | dominant | dominant | dominant | dominant | dominant | dominant | | 37162 | 35910 | 33678 | 33439 | 34679 | 33513 | 32080 | 31655 | 32375 | 30356 | 27757 | 26957 | |
| Antigenically novel, low immune escape, low virulence | Yes | High | dominant | dominant | dominant | dominant | dominant | dominant | dominant | dominant | dominant | dominant | dominant | dominant | | 29265 | 30088 | 25730 | 27621 | 27900 | 29513 | 24974 | 26971 | 20361 | 21096 | 17861 | 19438 | |
| | Yes | Low | dominant | dominant | dominant | dominant | dominant | dominant | dominant | dominant | dominant | dominant | dominant | dominant | | 29314 | 28771 | 24583 | 26699 | 27688 | 28090 | 23844 | 25925 | 20258 | 19645 | 16287 | 17911 | |
| | No | High | dominant | dominant | dominant | dominant | dominant | dominant | dominant | dominant | dominant | dominant | dominant | dominant | | 31414 | 31635 | 27647 | 30019 | 30023 | 30983 | 27161 | 29531 | 23233 | 23432 | 20364 | 22674 | |
| | No | Low | dominant | dominant | dominant | dominant | dominant | dominant | dominant | dominant | dominant | dominant | dominant | dominant | | 30686 | 30585 | 26937 | 28453 | 29382 | 29675 | 26189 | 27539 | 22405 | 21855 | 19518 | 20177 | |
| Antigenically Omicron-like, high immune escape, low virulence | Yes | High | dominant | dominant | dominant | dominant | dominant | dominant | dominant | dominant | dominant | dominant | dominant | dominant | | 29020 | 27749 | 24274 | 24967 | 26467 | 25652 | 22659 | 23544 | 18999 | 17937 | 15628 | 15761 | |
| | Yes | Low | dominant | dominant | dominant | dominant | dominant | dominant | dominant | dominant | dominant | dominant | dominant | dominant | | 27962 | 26701 | 23983 | 23631 | 25528 | 24431 | 22470 | 22108 | 18007 | 16492 | 15462 | 14154 | |
| | No | High | dominant | dominant | dominant | dominant | dominant | dominant | dominant | dominant | dominant | dominant | dominant | dominant | | 30194 | 29279 | 26570 | 27073 | 27907 | 27012 | 24821 | 25329 | 20949 | 19674 | 18514 | 17817 | |
| | No | Low | dominant | dominant | dominant | dominant | dominant | dominant | dominant | dominant | dominant | dominant | dominant | dominant | | 30243 | 28027 | 26103 | 25495 | 27818 | 25925 | 24449 | 23855 | 20812 | 17951 | 17626 | 16566 | |
| Antigenically novel, high immune escape, low virulence | Yes | High | dominant | dominant | dominant | dominant | dominant | dominant | dominant | dominant | dominant | dominant | dominant | dominant | | 22455 | 21296 | 18141 | 20024 | 20934 | 19883 | 16704 | 19251 | 10393 | 8423 | 7282 | 9105 | |
| | Yes | Low | dominant | dominant | dominant | dominant | dominant | dominant | dominant | dominant | dominant | dominant | dominant | dominant | | 23523 | 22703 | 18823 | 21987 | 22053 | 21512 | 17795 | 21358 | 11907 | 10853 | 8640 | 12548 | |
| | No | High | dominant | dominant | dominant | dominant | dominant | dominant | dominant | dominant | dominant | dominant | dominant | dominant | | 24078 | 22230 | 17902 | 20548 | 22721 | 20832 | 16935 | 19181 | 12665 | 9520 | 7387 | 10239 | |
| | No | Low | dominant | dominant | dominant | dominant | dominant | dominant | dominant | dominant | dominant | dominant | dominant | dominant | | 23177 | 22514 | 17496 | 19827 | 21570 | 20831 | 16461 | 18842 | 11288 | 9780 | 7121 | 9433 | |
| Antigenically Omicron-like, low immune escape, high virulence | Yes | High | dominant | dominant | dominant | dominant | dominant | dominant | dominant | dominant | dominant | dominant | dominant | dominant | | dominant | dominant | dominant | dominant | dominant | dominant | dominant | dominant | dominant | dominant | dominant | dominant | |
| | Yes | Low | dominant | dominant | dominant | dominant | dominant | dominant | dominant | dominant | dominant | dominant | dominant | dominant | | dominant | dominant | dominant | dominant | dominant | dominant | dominant | dominant | dominant | dominant | dominant | dominant | |
| | No | High | dominant | dominant | dominant | dominant | dominant | dominant | dominant | dominant | dominant | dominant | dominant | dominant | | dominant | dominant | dominant | dominant | dominant | dominant | dominant | dominant | dominant | dominant | dominant | dominant | |
| | No | Low | dominant | dominant | dominant | dominant | dominant | dominant | dominant | dominant | dominant | dominant | dominant | dominant | | dominant | dominant | dominant | dominant | dominant | dominant | dominant | dominant | dominant | dominant | dominant | dominant | |
| Antigenically novel, low immune escape, high virulence | Yes | High | dominant | dominant | dominant | dominant | dominant | dominant | dominant | dominant | dominant | dominant | dominant | dominant | | dominant | dominant | dominant | dominant | dominant | dominant | dominant | dominant | dominant | dominant | dominant | dominant | |
| | Yes | Low | dominant | dominant | dominant | dominant | dominant | dominant | dominant | dominant | dominant | dominant | dominant | dominant | | dominant | dominant | dominant | dominant | dominant | dominant | dominant | dominant | dominant | dominant | dominant | dominant | |
| | No | High | dominant | dominant | dominant | dominant | dominant | dominant | dominant | dominant | dominant | dominant | dominant | dominant | | dominant | dominant | dominant | dominant | dominant | dominant | dominant | dominant | dominant | dominant | dominant | dominant | |
| | No | Low | dominant | dominant | dominant | dominant | dominant | dominant | dominant | dominant | dominant | dominant | dominant | dominant | | dominant | dominant | dominant | dominant | dominant | dominant | dominant | dominant | dominant | dominant | dominant | dominant | |
| Antigenically Omicron-like, high immune escape, high virulence | Yes | High | dominant | dominant | dominant | dominant | dominant | dominant | dominant | dominant | dominant | dominant | dominant | dominant | | dominant | dominant | dominant | dominant | dominant | dominant | dominant | dominant | dominant | dominant | dominant | dominant | |
| | Yes | Low | dominant | dominant | dominant | dominant | dominant | dominant | dominant | dominant | dominant | dominant | dominant | dominant | | dominant | dominant | dominant | dominant | dominant | dominant | dominant | dominant | dominant | dominant | dominant | dominant | |
| | No | High | dominant | dominant | dominant | dominant | dominant | dominant | dominant | dominant | dominant | dominant | dominant | dominant | | dominant | dominant | dominant | dominant | dominant | dominant | dominant | dominant | dominant | dominant | dominant | dominant | |
| | No | Low | dominant | dominant | dominant | dominant | dominant | dominant | dominant | dominant | dominant | dominant | dominant | dominant | | dominant | dominant | dominant | dominant | dominant | dominant | dominant | dominant | dominant | dominant | dominant | dominant | |
| Antigenically novel, high immune escape, high virulence | Yes | High | dominant | dominant | dominant | dominant | dominant | dominant | dominant | dominant | dominant | dominant | dominant | dominant | | dominant | dominant | dominant | dominant | dominant | dominant | dominant | dominant | dominant | dominant | dominant | dominant | |
| | Yes | Low | dominant | dominant | dominant | dominant | dominant | dominant | dominant | dominant | dominant | dominant | dominant | dominant | | dominant | dominant | dominant | dominant | dominant | dominant | dominant | dominant | dominant | dominant | dominant | dominant | |
| | No | High | dominant | dominant | dominant | dominant | dominant | dominant | dominant | dominant | dominant | dominant | dominant | dominant | | dominant | dominant | dominant | dominant | dominant | dominant | dominant | dominant | dominant | dominant | dominant | dominant | |
| | No | Low | dominant | dominant | dominant | dominant | dominant | dominant | dominant | dominant | dominant | dominant | dominant | dominant | | dominant | dominant | dominant | dominant | dominant | dominant | dominant | dominant | dominant | dominant | dominant | dominant | |

*OT: Omicron-targeted vaccine in Q4 2022; 2*OT: Omicron-targeted vaccines in Q4 2022 and Q2 2023; OT+M: Omicron-targeted vaccine in Q4 2022 and multivalent vaccine in Q2 2023; 30+: administered to people aged ≥30 years; 60+ administered to people aged ≥60 years; H: high coverage; L: low coverage*

*NB: the missing column for 'nil vaccination' is the comparator; the left panel of ICERs are all compared to nil vaccination (in the same row) under more stringent PHSMs; the right panel of ICERs are all compared to nil vaccination (in the same row, final empty column) under less stringent PHSMs.*

**Supplementary Figure 19: Heat map for policy options (over 12 months, considering all nine future SARS-CoV-2 scenarios equally likely) of their overall rank in (a) the main analysis, (b) with HALYs valued at AUD 35,000, (c) with HALYs valued at AUD 140,000, (d) assuming people dying of COVID-19 have 1.5 times the mortality and 1.25 times the morbidity of the average person of the same sex and age (as opposed to 2 and 1.5 times respectively as used in the main analysis) and (e) using UK Treasury-recommended discount rates (1.5% per annum for HALYs, 3.5% per annum for costs). The top 20 policies from the main analysis are shown.**

| | | | | Ranks | | | | |
|---|---|---|---|---|---|---|---|---|
| Vaccine schedule | Respirator stockpile | Mask wearing | Stage policy | Main analysis | HALYs valued at AUD 35k | HALYs valued at AUD 140k | Alternative morbidity and mortality analysis | Alternative discounting |
| OT+M, 30+, H | No | Increased | Higher | 1 | 1 | 1 | 1 | 1 |
| 2*OT, 30+, H | No | Increased | Higher | 2 | 1 | 4 | 2 | 2 |
| 2*OT, 30+, H | Yes | Increased | Higher | 3 | 7 | 3 | 3 | 6 |
| OT+M, 30+, H | Yes | Increased | Higher | 4 | 8 | 2 | 4 | 5 |
| OT, 30+, H | No | Increased | Higher | 5 | 8 | 10 | 7 | 7 |
| 2*OT, 30+, H | No | No change | Higher | 6 | 5 | 7 | 5 | 3 |
| OT+M, 30+, H | No | No change | Higher | 7 | 4 | 5 | 5 | 4 |
| OT+M, 30+, H | Yes | No change | Higher | 8 | 11 | 5 | 8 | 9 |
| OT+M, 60+, H | No | Increased | Higher | 8 | 3 | 11 | 10 | 11 |
| 2*OT, 30+, H | Yes | No change | Higher | 10 | 13 | 8 | 11 | 10 |
| 2*OT, 60+, H | No | Increased | Higher | 10 | 5 | 12 | 12 | 12 |
| OT, 30+, H | Yes | Increased | Higher | 12 | 12 | 9 | 9 | 8 |
| OT, 60+, H | No | Increased | Higher | 13 | 10 | 17 | 13 | 14 |
| OT+M, 30+, H | No | Increased | Lower | 14 | 18 | 16 | 13 | 13 |
| 2*OT, 60+, H | Yes | Increased | Higher | 14 | 20 | 27 | 16 | 15 |
| OT, 30+, H | No | No change | Higher | 14 | 16 | 14 | 16 | 15 |
| OT+M, 60+, H | Yes | Increased | Higher | 17 | 15 | 13 | 15 | 17 |
| OT+M, 30+, H | No | No change | Lower | 18 | 26 | 29 | 18 | 19 |
| 2*OT, 30+, H | No | No change | Lower | 19 | 21 | 29 | 19 | 19 |
| OT+M, 30+, H | Yes | Increased | Lower | 20 | 33 | 28 | 20 | 18 |
| 2*OT, 30+, H | No | Increased | Lower | 20 | 14 | 19 | 20 | 22 |
| OT+M, 60+, H | No | No change | Higher | 20 | 21 | 31 | 22 | 21 |

*OT: Omicron-targeted vaccine in Q4 2022; 2*OT: Omicron-targeted vaccines in Q4 2022 and Q2 2023; OT+M: Omicron-targeted vaccine in Q4 2022 and multivalent vaccine in Q2 2023; 30+: administered to people aged ≥30 years; 60+ administered to people aged ≥60 years; H: high coverage; L: low coverage*

### Supplementary Figure 20: Tornado plots for a single* policy and future SARS-CoV-2 variant scenario modelled over 12 months, showing input parameter uncertainty that generated the most uncertainty in key model outputs

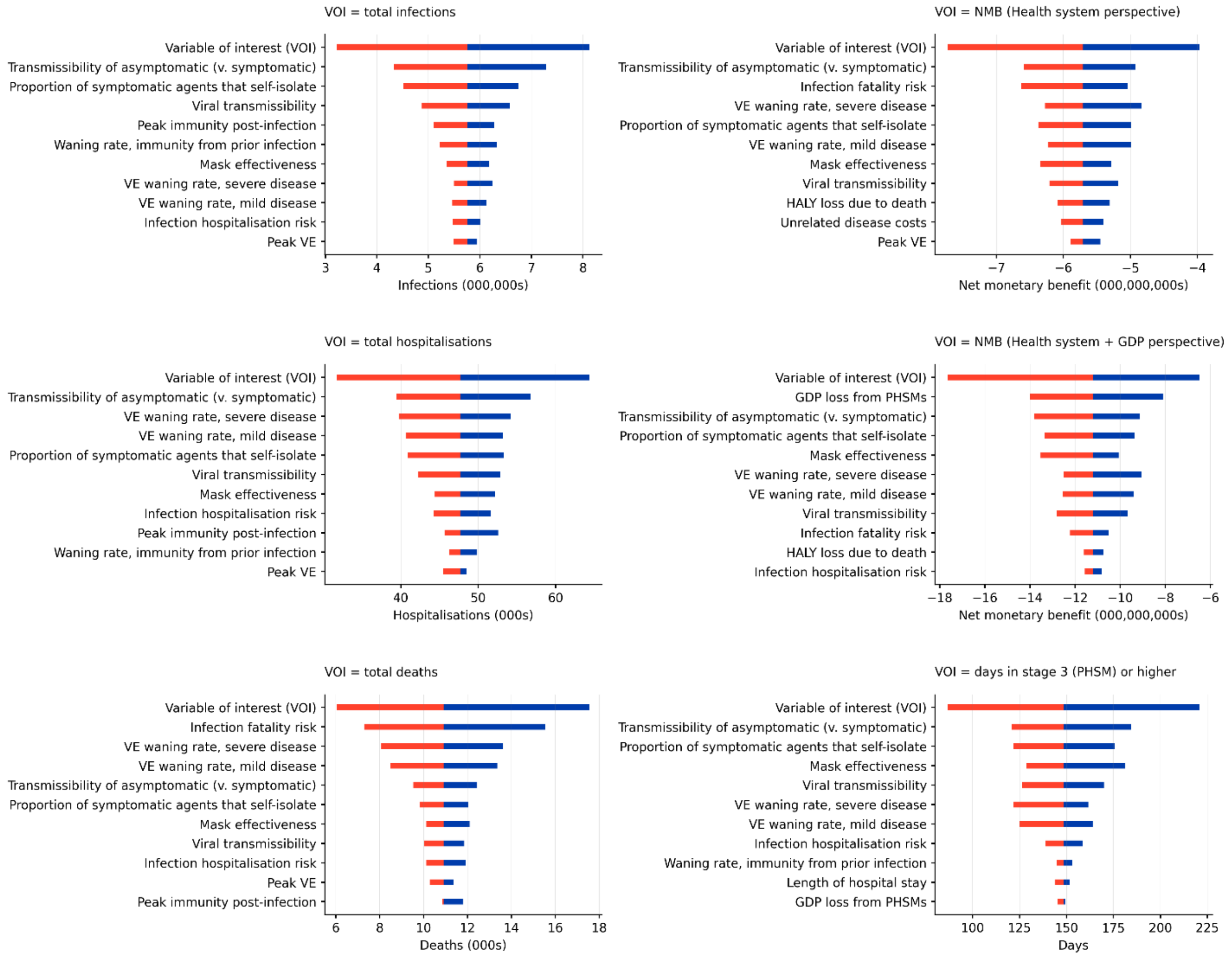

**variant with high virulence, antigenically novel and high immune escape capacity modelled in the context of respirator substitution and increased mask-wearing policies active, higher stringency PHSMs and a vaccine schedule with an Omicron-targeted vaccine administered in the final quarter of 2022 and a multivalent vaccine in the second quarter of 2023 to individuals aged ≥60 with high uptake.*

*Bars indicate the average of the variable of interest for the runs with the lowest and highest quintile values of each input parameter. The top bar in each plot is the average of the lowest and highest quintiles of the variable of interest, which is an upper bound on the width of the bars below it. Note we expect that some input variables feature in these plots primarily due to chance (e.g., GDP loss from PHSMs appearing in the plot for days in stages 3 and above).*

*VOI: variable of interest; VE: vaccine effectiveness; HALY: health-adjusted life year; GDP: gross domestic product; PHSMs: public health and social measures; severe disease: hospitalisation, ICU admission or death; mild disease: asymptomatic or symptomatic infection; proportion of symptomatic agents that self-isolate: the proportion of symptomatic agents that self-isolate when they become symptomatic.*

### Appendix

#### Agent-based model

##### *Overview and input parameters*

We used an agent-based model (ABM) with 5,000 agents and a daily cycle length (model GitHub repository available at https://github.com/population-interventions/CovidABM). Each agent was assigned attributes including age (in 10-year strata), essential worker status, and a household. Depending on the scenario, agents were also proportionally assigned vaccine status, mask usage, and other characteristics. Each day, agents move around the 2-dimensional model space, with all infected agents having a chance to infect susceptible agents at their location.

The model space contains gathering locations, which all agents have a higher probability of visiting, and home locations that are frequently visited by members of each household. Agent movement includes returning to their household, moving around the community, visiting nearby gathering locations, "superspreading" (by jumping to a random gathering location), and the possibility of noticing the presence of other agents and attempting to avoid them. The weights of various types of movement depend on the public health measures in place at any given time, as well as heterogeneity in agent behaviour (with a large age component). Agents with symptomatic infection have a chance of isolating by staying home and reducing their infectiousness.

At the end of the movement phase, each infectious agent has a chance to infect each non-infected agent at its location. The probability of transmission depends on the infectiousness and susceptibility of the two agents involved. After a non-infective period, the infectiousness of an agent increases linearly to a maximum at their peak infectiousness time, then linearly decreases to zero (Appendix Table 1). Infectiousness is also modified by whether the agent wears a mask, whether they are symptomatic, and whether they try to isolate. These parameters have per-agent draws for heterogeneity. Susceptibility to infection is modified by mask wearing and heterogeneity by age. Both infectiousness and susceptibility to infection are modified by vaccine status and recovery from previous infection, dependent upon on the type of vaccine received (or the variant responsible for previous infection), the variant responsible for exposure, and waning since vaccination or previous infection.

The model simulates the population of Victoria using scaling of the 5,000 agents. Each infected agent represents a number of people, with this number inherited from the agent that infected them. Infected agents are split or merged with similar agents when there are too few or too many infected agents in the model. The model attempts to maintain between 260 and 290 infected agents, except when the number of infections reaches the upper or lower scale bounds of the model. At low scales the model essentially operates by zooming in on the part of Victoria where infection is spreading, while being able to scale up to simulate all of Victoria in a large wave. By scaling up and down, the impact on the total number of infected, vaccination, recovered, etc., people in a population the size of Victoria (6.6 million) was tallied and retained. These metrics dynamically determine public health measures and community carefulness in the model.

Input parameters have probability distributions from which values are drawn for each of the 500 iterations for each of the Monte Carlo analyses in the ABM. The same random draws of input parameters that apply to all agents for the $i^{th}$ of 500 iterations is used across all 936 scenarios (104 policy options by 9 variant scenarios), meaning within each iteration we can compared policy options at the same random draw values of inputs such as vaccine effectiveness, mask effectiveness, etc. Key ABM input parameters are shown in Appendix Table 1. All variables (except the 'carefulness' parameter) were specified a priori and not subject to amendments in calibration. The values of the 'carefulness' parameter were set in calibration (see next section).

**Appendix Table 1: Key agent-based model input parameters**

| Parameter | Estimate |
|---|---|
| Agent infectiousness | Agent infectiousness on each day is parameterised by agent-level draws for peak infectiousness and the parameters listed below. Infectiousness prior to the peak is linearly interpolated to 0% of peak infectiousness on day 0, and infectiousness after the peak is linearly interpolated to zero at the end of the infectious period. |
| Incubation period (time from infection to development of symptoms) (days) | Gamma distribution with mean 3.485, SD 1.1953 (95% UI 1.55 to 6.19).[1] A theoretical time to symptom development was specified for all agents, but only manifest (e.g. for self-isolation) for the proportion of agents defined as symptomatic. |

| Parameter | Estimate |
|---|---|
| Time from symptom development to peak infectiousness (days) | Gamma distribution with mean 3.7486, SD 2.0499.[2] |
| Time from initially becoming infectious to peak infectiousness (days)* | Gamma distribution with mean 1.8691, SD 1.202.[2] |
| Time from peak infectiousness to end of infectiousness (days)* | Gamma distribution with mean 2.9989, SD 1.8278.[2] |
| Mean adherence with isolation of infected cases | Global beta distribution (beta(450.3, 23.7); mean = 95%, SD = 1%) |
| Infectiousness of asymptomatic v. symptomatic cases per contact | RR 0.58 (95% CI 0.34 to 0.99)[3] (parameterised as a log normal distribution with median = -0.545 and SD = 0.270) |
| Household size | Beta distribution with median = 3. Beta(2.2, 2.2) scaled to [1, 5], draws rounded to nearest integer |
| Chance of an infected household contact of a known case becoming known, per day. | 100% |
| Proportion of undetected symptomatic cases who spontaneously reported themselves | Beta(6,6) |
| Time for undetected symptomatic cases spontaneous self-reporting | At symptom development (see above) |
| Transmission multiplier of person who is complying with their isolation | 0.33 |
| Relative susceptibility to infection, by age (OR for infection given exposure) | 0 – 9 years: 0.34<br>10-19: 0.67<br>20 – 59 years: 1<br>60-69: 1.23<br>≥70 years: 1.47[4]<br>Uncertainty on all values +/- 15% SD |
| Relative 'carefulness' multiplier to susceptibility, by age | 0 – 9 years: 1.882<br>10 – 19 years: 1.179<br>20 – 59 years: 1<br>60 – 69 years: 0.975<br>70 – 79 years: 0.667<br>80 – 89 years: 0.646<br>≥90 years: 0.646<br>Uncertainty on all values +/- 15% SD |

**correlated -0.46*

#### *Incubation period and agent infectiousness*

We specified the incubation period (time from infection to symptoms) as per Manica et al.[1] for Omicron, namely Gamma(8.5,0.41) giving median 3.35 days, 95% UI 1.55 to 6.19, mean 3.49 days. A theoretical time to symptoms was specified for all agents, but only manifest (e.g., for self-isolation) for the proportion of agents set as symptomatic elsewhere.

We specified the time from first symptoms to peak infectiousness using data from Hakki et al[2] (a detailed analysis of household close contacts for pre-Alpha to Delta infections), with a Gamma distribution (3.344, 1.121) obtained by fitting to the Hakki et al. results. This gamma distribution was the best we could achieve to approximate the median (3.4 days) and interquartile interval (2.6 to 5.6 days) given by Hakki et al.[i]; our Gamma distribution gave median 3.38, 25th percentile of 2.24 and a 75th percentile of 4.86, and mean 3.75, 95% UI 0.87 to 8.71.

We specified the time from initially becoming infectious to peak infectiousness also using data from Hakki et al., with a Gamma distribution (2.418, 0.773), median 1.62 days, 95% UI 0.30 to 4.86, mean 1.87 days. Likewise, we specified the time from peak infectiousness to end of infectiousness with a Gamma distribution (2.692, 1.114), median 2.64 days, 95% UI 0.55 to 7.50, mean 3.00 days. Both distributions were good fits to output presented by

[i] Hakki et al. actually reported median 3, interquartile range 3 to 6, which is impossible to parameterize. We therefore assumed that the lower quartile (before rounding) must be close to 2.5 (specified as 2.6) and the median must be close to 3.5 (specified as 3.6). Similarly, we set the 75th percentile to 5.6.

Hakki et al. Following Hakki et al, we correlated these distributions -0.46 in the simulations (i.e., a longer time from initial to peak infectiousness predicted a shorter time from peak infectiousness to end of infectiousness).

Combining these input parameters in the simulation, the median days from infection to end of infectiousness was 10 days (95% range across agents 6 to 17 days). The generation time (average time from infection to infection of the next agent; in 'natural state' with no PHSMs) was 7.6 to 7.7 days. We deemed this a good enough fit to the generation time estimated by Manica et al. for Omicron of 6.8 days (95% credible interval 5.72 to 8.60 days). Our simulations predicted that about 20% of agents were infectious before becoming symptomatic, agreeing reasonably well with Hakki et al.'s 20% to 25% empirical estimate.

In the simulation, agents had no infectiousness on day 0 (the day they were infected), and thereafter their level of infectiousness was determined for the integer value of days since infection from an equation with linear increase in infectiousness from zero to peak, and linear decrease from peak to end of infectiousness, using the above agent-specific draws from distributions.

#### ABM calibration

Having set *a priori* variables as per Appendix Table 1, we calibrated the model to achieve $R_0$s of 8 and 10 in a situation with no vaccination, previous infection, contact tracing, or public health and social measures (PHSMs). $R_0$s were found by introducing a single infected agent into the naïve population and counting infections caused by case zero, 1000 times. The key variable 'tuned' was the base transmissibility of the virus, i.e., the probability of a single average interaction between an infected agent and susceptible agent resulting in infection of the susceptible agent. Spatial parameters were also adjusted to allow for a range of transmissibility values to achieve the required $R_0$s.

We then used a separate model to initiate the infection and vaccination status of the 5,000 agents by approximating vaccination, time since vaccination, and cumulative infection with variation by age as in Victoria on 1 April 2022 (Appendix Table 2). Infections were randomly assigned to agents, with less probability for those who were vaccinated (and allowing for waning vaccine immunity over time using a previously published logistic regression equation discussed in greater detail below).[5] No single-vaccinated agents were modelled. Pre-Omicron past infection was uncommon in Victoria and was ignored. Transmissibility was drawn uniformly from the range corresponding to $R_0$s 8 and 10. The proportion of previously infected agents was set to double that of reported cases in Victoria to allow for under-notification.[6]

With $R_0$ calibrated and baseline infection and vaccination status of agents assigned, we then iteratively calibrated the PHSMs in Stage 2 to achieve, over an average of 2,000 runs, 1.2 million infections in the 60 days following the 1st of April 2022. This corresponds to an average of 20,000 infections per day, which is twice the average daily reported cases in this period to allow for under-notification. In parallel, the 'carefulness' parameter in Appendix Table 1 was adjusted to achieve the age distribution of infections occurring in Victoria in the same period (Appendix Table 3). The 'carefulness' parameter is – we argue – theoretically justified, in that as the pandemic has evolved citizens are increasingly dynamically responding to infection rates likely in an age-dependent manner (e.g., working-age individuals working from home and avoiding public transport when infection rates are high, elderly people at highest risk of severe illness being particularly cautious to avoid infection).

**Appendix Table 2: Age, vaccination status and cumulative infection of the 5,000 initiating agents in the ABM**

| Age (years) | % distribution of population | % double vaccinated as of 1 April 2022 | % triple vaccinated as of 1 April 2022 | Cumulative infection as of 1 April 2022 |
|---|---|---|---|---|
| 0-4 | 5.7% | - | - | 32.0% |
| 5-11 | 8.7% | 30.2% | - | 34.5% |
| 12-17 | 6.9% | 95.6% | 12.4% | 45.6% |
| 18-29 | 16.5% | 87.1% | 44.5% | 48.8% |
| 30-39 | 15.5% | 93.8% | 56.2% | 40.3% |
| 40-49 | 12.8% | 96.4% | 68.2% | 34.7% |
| 50-59 | 12.0% | 96.4% | 74.4% | 25.7% |

| | | | | |
|---|---|---|---|---|
| 60-69 | 10.2% | 97.1% | 81.1% | 17.2% |
| 70+ | 11.6% | 98.8% | 88.1% | 11.3% |
| *All* | - | *83.5%* | *53.1%* | *32.9%* |

#### Appendix Table 3: Calibration target of infections by age, 60 days following 1 April 2022

| Age (years) | Target distribution of infections by age | Fraction of age group infected in this period |
|---|---|---|
| 0-9 | 12.50% | 19% |
| 10-19 | 13.00% | 19% |
| 20-29 | 14.10% | 19% |
| 30-39 | 18.90% | 22% |
| 40-49 | 15.30% | 22% |
| 50-59 | 11.80% | 18% |
| 60-69 | 7.80% | 14% |
| 70-79 | 4.30% | 11% |
| 80-89 | 1.80% | 10% |
| 90+ | 0.60% | 10% |

### Modelled scenarios

#### *Policy options*

Note all policy options could only be activated from the final quarter of 2022.

Suppression policy

Following calibration, the PHSMs associated with each stage were defined as shown in Appendix Table 4. The ABM cycled between these stages based on predicted pressure on health service infrastructure (defined by the predicted number of people hospitalised) and two suppression strategy policy options – higher and lower stringency – which aimed to keep peak hospital occupancy less than 200 and 400 per million residents respectively as per the (de)escalation rules in Appendix Table 5. The behaviour of agents was also dynamic based on infection incidence, to reflect the public modulating their interactions in response to outbreaks independent of government-imposed restrictions.

#### Appendix Table 4: Key input parameters by stage of public health and social measures

| | Stage 1 | Stage 2 | Stage 3 | Stage 4 | Stage 5 |
|---|---|---|---|---|---|
| Proportion of people who try to avoid contact with others (excluding their household)†* | 20% | 36% | 51% | 63% | 75% |
| Proportion of time spent trying to avoid contacts, for those that attempt to do so†* | 30% | 46% | 61% | 73% | 83% |
| Proportion of workers attending work in person† | 80% | 66% | 51% | 35% | 20% |
| Schools open (disable contact-avoiding behaviour among students) | Yes | Yes | Yes | No | No |
| Proportion of people that wear masks outside the home† (when increased mask wearing policy inactive) | | | | | |
| ≥20-year-olds | 6% | 16.3% | 45% | 66% | 80% |
| 10- to 19-year-olds | 4% | 10.9% | 30% | 44% | 53.3% |
| <10-year-olds | 2.7% | 7.2% | 20% | 29.3% | 35.6% |
| Proportion of mask wearing that is with a respirator# | | | | | |
| No government supply | 20% | 20% | 20% | 20% | 20% |
| Government supply policy active | 20% | 20% | 80% | 80% | 80% |
| Proportion of people that engage in super spreading behaviour each day (move to a random gathering location)† | 6% | 4.7% | 3.2% | 2.2% | 1.6% |

| | | | | | |
|---|---|---|---|---|---|
| Underlying frequency of visiting a random nearby gather location each day (e.g., a supermarket)† | 14.28% | 14.28% | 14.28% | 13% | 7.5% |
| Radius for determining whether a gather location counts as nearby | 8.8 | 8.63 | 8.12 | 6 | 5 |
| Maximum distance moved by an agent each day | 14 | 11.56 | 9 | 7.5 | 6 |

*†ORs of 2, 4 and 6 are applied to the proportion of people that wear a mask, isolation compliance, proportion of people that avoid others, and proportion of time spent avoiding others for those aged 50-59, 60-69 and ≥70 years respectively to capture increasing infection avoidance behaviour in older age groups. Reciprocals of these ORs are applied to the daily chance of visiting a gather location, and daily chance of superspreading. Note agents ≥60 years old are excluded from the category of workers.*

**In stages 1-3, the proportion of people who try to avoid contact with others and the proportion of time spent trying to avoid contacts for those attempting to do so are dynamic, based on the average number of infections over the last 7 days. Proportions are as written above if the 7-day average of infections is <5,000. The proportion of people who try to avoid contacts and proportion of time spent avoiding increase to a maximum of 15 and 10 percentage points higher than those written above, respectively, at 32,000 daily infections averaged over the last seven days. The region between 5,000 and 32,000 is linearly interpolated.*

*#Only applies to those aged ≥10 years.*

#### Appendix Table 5: Triggers for escalation and de-escalation of stages of public health and social measures, by suppression policy

| Suppression policy | Triggers |
|---|---|
| Triggers for *lower stringency* suppression policy | Escalation<br>If average expected number of people in hospital due to COVID-19 10-14 days (inclusive) into the future is:<br>- >600 per million → Stage 5<br>- >400 per million → Stage 4<br>- >270 per million → Stage 3<br>- >180 per million → Stage 2<br>De-escalation<br>If no de-escalation in last 7 days, and average expected number of people in hospital 10-14 days (inclusive) into the future is:<br>- <450 per million → Stage 4 if in Stage 5<br>- <300 per million → Stage 3 if in stage 4 or 5<br>- <200 per million → Stage 2 if in Stage 3, 4 or 5<br>- <140 per million → Stage 1 |
| Triggers for *higher stringency* suppression policy | Escalation:<br>If average expected number of people in hospital 10-14 days (inclusive) into the future is:<br>- >300 per million → Stage 5<br>- >200 per million → Stage 4<br>- >130 per million → Stage 3<br>- >90 per million → Stage 2<br>De-escalation:<br>If no de-escalation in last 7 days, and average expected number of people in hospital 10-14 days (inclusive) into the future is:<br>- <230 per million → Stage 4 if Stage 5<br>- <150 per million → Stage 3 if in stage 4 or 5<br>- <100 per million → Stage 2 if in Stage 3, 4 or 5<br>- <70 per million → Stage 1 |

### Respirator policy

At the time of writing, masks were no longer mandated in most indoor settings in Victoria but were still recommended. Masks remained mandatory in health care settings, and if a close contact of a COVID-19 case. These regulations, however, do not specify the type of mask that the public should use. We modelled the maintenance of a stockpile of 10 respirators per person for the whole Victorian population aged 10 years and older, that are distributed for use at four-week intervals (rotating masks on a five-day cycle with extra masks to cover for spoilage) if in stage 3 or higher. In essence, this changes the proportion of people wearing respirators from 20% at baseline to 80% (Appendix Table 4) in these stages while the respirator policy is active.

There is a paucity of high-quality evidence providing estimates of the degree of protection afforded by mask-wearing at the individual level in community settings for the transmission of SARS-CoV-2. A recent systematic review and meta-analysis[7] that examined the effectiveness of public health measures in reducing the incidence of COVID-19 reported a relative risk of infection of 0.47 (95% CI 0.29 to 0.75) associated with mask-wearing overall, based on six studies. The authors note, however, a high degree of heterogeneity between these studies and substantial risk of bias. In a World Health Organization-commissioned systematic review and meta-analysis published in 2020, protection afforded by wearing an N95 mask or “surgical or similar” mask was estimated at 95% (95% CI 34% to 100%) and 58% (95% CI 24% to 77%) respectively. This review, however, included literature regarding three betacoronaviruses (MERS-CoV, SARS-CoV and SARS-CoV-2) and studies in both healthcare and community-based settings,[8,9] limiting applicability to our model.

We elected to base our estimates of mask effectiveness on a test-negative case-control study enrolling individuals receiving COVID-19 test results from February to December 2021 in California, USA, which provides estimated effect sizes for protection stratified by type of mask.[10] Estimates were adjusted for vaccination status, household income, race/ethnicity, age, sex, state region and county population density. The primary analysis assessed self-reported mask use in indoor public settings during the 14 days preceding testing between those receiving positive and negative SARS-CoV-2 test results and found adjusted odds of infection of 0.44 (95% CI 0.24 to 0.82) associated with always using any type of mask or respirator. Adjusted odds for infection associated with using cloth masks, surgical masks and respirators were 0.44 (95% CI 0.17 to 1.17), 0.34 (95% CI 0.13 to 0.90) and 0.17 (0.05 to 0.64) respectively. Assuming 5% of ‘always’ mask use was N95 masks, these reported estimates were adjusted to the following for use in our model:

- Cloth/surgical masks: odds ratio (OR) 0.468 (or -0.76 on ln scale, with SD 0.31)
- N95 respirators: OR 0.204 (or -1.59 on ln scale, with SD 0.65)

We specified 100% correlation on draws of mask effectiveness and assumed the same risk reduction in onward transmission if wearing a mask.

#### Increase mask use policy

We modelled two levels of background mask-wearing compliance; one as per Appendix Table 4, and one whereby the proportion of individuals wearing masks was increased with an OR of 2 (i.e., 40% mask use shifts to 57%, 60% shifts to 75%, etc.). The proportion of people wearing cloth/surgical masks and respirators was then informed by the respirator policy outlined above.

#### Vaccination policy

Australia had a mixed rollout of mRNA-based COVID-19 vaccines (BNT162b2 and mRNA-1273) and an adenovirus vector-based vaccine (ChAdOx1) for the primary series and predominantly mRNA-based vaccines for further boosting. In our model, we combined all currently available vaccines into one generic ‘first generation’ vaccine with effectiveness as outlined below. We then specified two types of ‘next generation’ vaccines with vaccine effectiveness (VE) expected to be superior to first-generation vaccines against emerging variants to reflect advances in vaccination technology – (i) Omicron-targeted vaccines that specifically target one or more subvariants of Omicron and (ii) multivalent vaccines that confer broader spectrum protection against several variants or viruses (e.g., by targeting more conserved regions of the virus or targeting the spike protein of multiple variants within a single vaccine formulation). Examples of Omicron-targeted vaccines include monovalent candidates developed by Moderna (mRNA-1273.529) and by Pfizer and BioNTech, both targeting the BA.1 subvariant of Omicron.[11,12] Examples of multivalent vaccines include the Spike Ferritin Nanoparticle vaccine developed by the Walter Reed Army Institute of Research[13] and Vaxart’s oral formulation, VXA-CoV2-1.[14,15] Both Moderna and Pfizer have also developed Omicron BA.1-adapted bivalent formulations that have shown superiority over current-generation vaccines in early human trials.[12,16] Given that next generation efficacy data is sparsely available, these vaccines are conjectured to have likely VE against emerging Omicron-like variants and novel variants as outlined below (Appendix Table 6).

We specified 13 hypothetical vaccination schedules as shown in Appendix Table 6. These were informed by current vaccination policy discussions in the Australian context.

**Appendix Table 6: Vaccination schedules treated as policy options**

| Scenario | Apr-Jun 2022 | Jul-Sep 2022 | Oct-Dec 2022 | Jan-Mar 2023 | Apr-Jun 2023 | Jul-Sep 2023 |
|---|---|---|---|---|---|---|
| a) Nil further beyond June 2022 | FG* | | | | | |
| b) 1 OT | FG* | | OT | | | |

| | | | | | |
|---|---|---|---|---|---|
| c) 2 OT | FG* | | OT | | OT |
| d) 1 OT then 1 MV | FG* | | OT | | MV |

**first-generation vaccine, administration to match third and fourth dose vaccination coverage as of June 30, 2022, in Victoria.*
*OT: Omicron-targeted; MV: multivalent; FG: first-generation*

Vaccinations beyond June 2022 were applied either to ≥30-year-olds or ≥60-year-olds, with either low uptake (50% for ≥60-year-olds, 25% for 30- to 60-year-olds) or high uptake (75% for ≥60-year-olds, 50% for 30- to 60-year-olds). All agents who had received at least three vaccine doses by the end of June 2022 were eligible to receive ongoing vaccinations, with probability independent between rounds of vaccination (e.g. if an agent was drawn to be vaccinated in Oct-Dec 2022, this did not alter that agent's probability to be vaccinated again in Apr-May 2023).

#### *Future variant scenarios*

We specified nine future SARS-CoV-2 variant scenarios, formed by cross-classifying virulence (2 options; low and high), antigenic profile (2 options; antigenically Omicron-like and novel) and associated immune escape capacity (2 options, low or high), with one residual 'no future variant of concern' scenario; these are outlined below. All new variants modelled emerged from November 2022 following default periods of Omicron BA.1/2 and BA.4/5 dominance between April and October 2022 as occurred in Victoria. New variants retained the same innate transmissibility as earlier Omicron variants ($R_0$ 8-10).

##### Virulence

Hopes that future variants of concern will incrementally lose virulence as occurred with the Omicron variant are unfounded; virulence arises largely independent of a variant's ability to transmit or evade the immune response.[17,18] Accordingly, new variants were characterized as either high or low (approximating Omicron) virulence. We first specified low virulence by calibrating international estimates to the Victorian Omicron wave experience in April and May 2022, as outlined below. For the high virulence variant, we assumed $4^{1}= 4$, $4^{0.75}= 2.83$, $4^{0.5}= 2$ and $4^{0.25}= 1.41$ ratio increases in the infection fatality risk (IFR), risk of ICU admission, risk of hospital admission, and risk of being symptomatic, respectively, compared to low virulence variants.

##### Infection fatality risk

We fitted a logistic regression model to Institute for Health Metrics and Evaluation (IHME) estimates of age-specific IFRs (≥10 years of age) for the ancestral strain of SARS-CoV-2 (Table 1 of [19]). This gave a coefficient of 0.1049 for age in years. We then used this age variation in the calibrated ABM applied to 60 days post 1 April 2022 in Victoria that included vaccination status and prior infection status of agents. Accompanied by an intercept or constant of -11.9861, this equation when combined with vaccine effectiveness and natural protection of agents (below) generated approximately the observed number of deaths in Victoria during the first 60 days of the model run (1 Apr 2022 to 30 May 2022) when the model was also generating 20,000 infections per day (see calibration section above). The final IFR equation is shown below, as well as that similarly set as a constant IFR among the unvaccinated and previously uninfected for <10-year-olds. The equation for the high virulence variant simply had ln(4) = 1.3863 added (i.e. to achieve four-fold higher IFRs for more virulent variants).

- Current Omicron and low virulence new variant:
  - ≥10 years: logit(IFR) = 0.1049*age – 11.9861 (SD = 0.205 on logit scale)
  - <10 years: logit (IFR) = -11.6051 (SD = 0.269)
- High virulence new variant:
  - ≥10 years: logit(IFR) = 0.1049*age – 10.5999 (SD = 0.205)
  - <10 years: logit(IFR) = -10.2189 (SD = 0.269)

IFRs for unvaccinated and previously uninfected people, for low and high virulence variants, are shown in Appendix Figure 1. These IFRs are then modified according to vaccination and previous infectious status (with waning) for each agent in the ABM as outlined below.

**Appendix Figure 1: IFRs among unvaccinated and previously uninfected individuals, for low and high virulence variants**

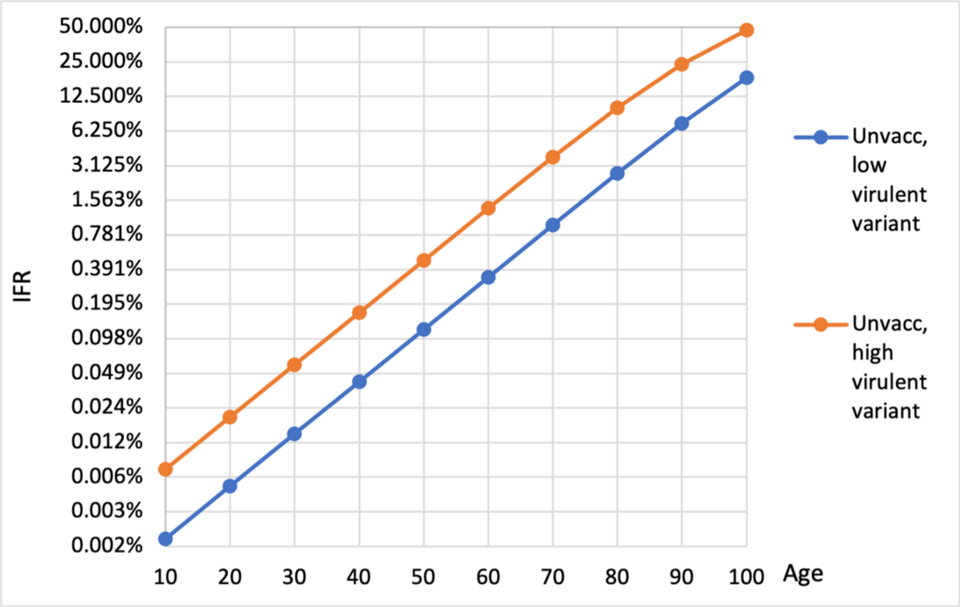

### ICU admission risk

To obtain the age-dependent probability of ICU admission given infection, we used the age gradient published by Knock et al. for ancestral SARS-CoV-2.[20] For those aged ≥30 years the probability of ICU admission given infection increases approximately linearly by age on a logit scale, described by the equation logit(ICU | inf) = 0.1293*age – 12.866. Risks in Knock et al. of ICU admission given infection on the logit scale for those aged 0 to 9 years and 10 to 29 years were -9.2629 and -10.4237 respectively. ICU admission risk beyond 80 years was set as that for an 80-year-old. We then reduced ICU admission risk for those aged 80-89 years by 25% and those aged ≥90 years by an additional 75% to reflect a lower likelihood of ICU admission in older age despite clinically severe disease.

These estimates were then adjusted during calibration (similarly to that above for IFR) for a low-virulence variant (approximating Omicron) to achieve the number of COVID-19-related ICU admissions in Victoria from 1 April to 30 May 2022 (on a background of vaccination and previous infection rates approximating that in Victoria during this period). We then applied a $4^{0.75}$= 2.83 ratio difference in ICU admission risk between the low and high virulence variants to give the following:

- Current Omicron and low-virulence new variant:
  - ≥90 years: logit(ICU | inf) = 0.129*age – 13.386 – (0.481+1.386) – 0.266 (SD = 0.1)
  - 80-89 years: logit(ICU |inf) = 0.129*age – 13.386 – (0.481 + 0.288) – 0.266 (SD = 0.1)
  - 30-79 years: logit(ICU | inf) = 0.129*age – 13.386 – 0.481 – 0.266 (SD = 0.1)
  - 10-29 years: logit(ICU | inf) = -10.943 – 0.481 – 0.266 (SD = 0.1)
  - <10 years: logit(ICU | inf) = -9.783 – 0.481 – 0.266 (SD = 0.1)
- High-virulence new variant:
  - ≥90 years: logit(ICU | inf) = 0.129*age – 12.346 – (0.481+1.386) – 0.266 (SD = 0.1)
  - 80-89 years: logit(ICU |inf) = 0.129*age – 12.346 – (0.481 + 0.288) – 0.266 (SD = 0.1)
  - 30-79 years: logit(ICU | inf) = 0.129*age – 12.346 – 0.481 – 0.266 (SD = 0.1)
  - 10-29 years: logit(ICU | inf) = -9.904 – 0.481 – 0.266 (SD = 0.1)
  - <10 years: logit(ICU | inf) = -8.743 – 0.481 – 0.266 (SD = 0.1)

The resultant infection ICU admission risks for unvaccinated and previously uninfected people, for low and high virulence variants, are shown in Appendix Figure 2. These risks are then modified according to vaccination and previous infectious status (with waning) for each agent in the ABM.

**Appendix Figure 2: Infection ICU admission risks among unvaccinated and previously uninfected individuals, for low and high virulence variants**

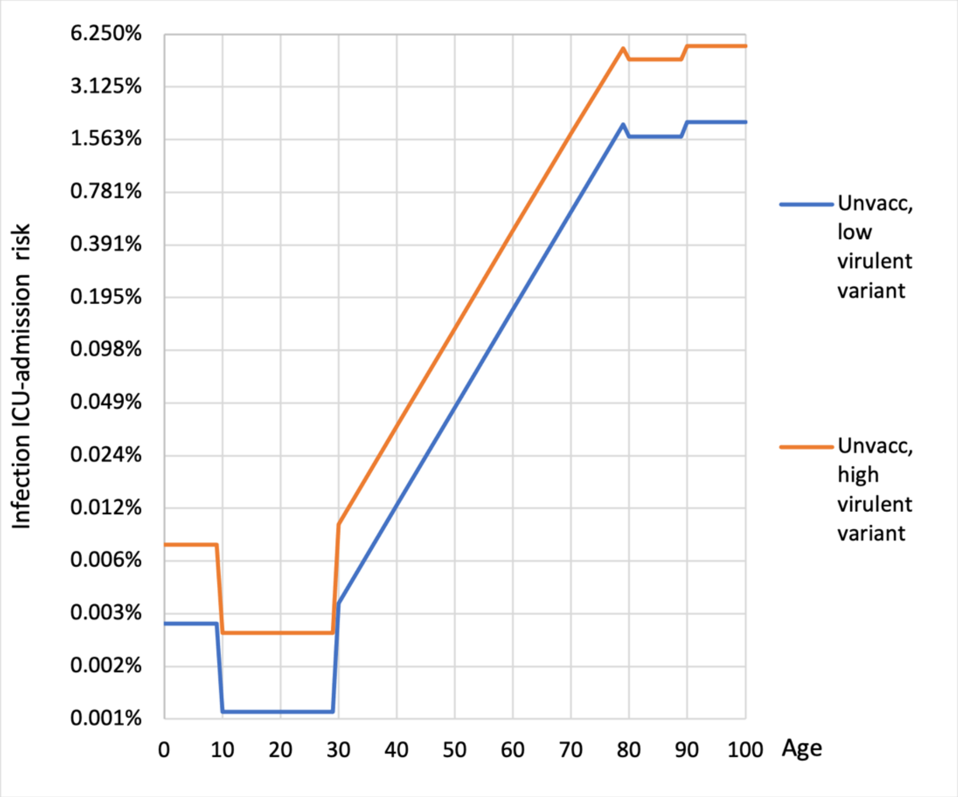

### Hospitalisation risk

To obtain the age-dependent probability of hospitalization given infection, age-variation is also based on data published by Knock et al.[20] For individuals aged ≥30 years, the Knock et al. probability of hospitalization given infection increases approximately linearly by age on the logit scale, described by the equation logit(hosp | inf) = 0.0997*age – 8.9113. For people younger than 30 years the risk of hospitalization given infection is not linear on the logit scale and as such averages of point estimates were used (-6.0308 for those aged 0 to 9 years and -7.1235 for those aged 10 to 29 years).

These estimates were then calibrated for a low-virulence variant (approximating Omicron, and as above for IFR and ICU) to achieve the number of COVID-19-related hospital admissions in Victoria from 1 April to 30 May 2022. We applied a $4^{0.5}$= 2 ratio difference in hospitalisation risk between the low and high virulence variants to give the following:

- Current Omicron and low-virulence new variant:
  - ≥30 years: logit(hosp | inf) = 0.0997*age – 9.258 – 0.601 (SD = 0.1), capped at risk for an 80-year-old (i.e. constant hospitalisation risk for those aged 80 years and older)
  - 10-29 years: logit(hosp | inf) = -7.470 – 0.601 (SD = 0.1)
  - <10 years: logit(hosp | inf) = -6.377 – 0.601 (SD = 0.1)
- High-virulence new variant:
  - ≥30 years: logit(hosp | inf) = 0.0997.age – 8.5647 – 0.601 (SD = 0.1), capped at risk for an 80-year-old
  - 10-29 years: logit(hosp | inf) = -6.777 – 0.601 (SD = 0.1)

- <10 years: logit(hosp | inf) = -5.684 – 0.601 (SD = 0.1)

The resultant infection hospitalisation risks for unvaccinated and previously uninfected people, for low and high virulence variants, are shown in Appendix Figure 3. These risks are modified according to vaccination and previous infectious status (with waning) for each agent in the ABM.

**Appendix Figure 3: Infection hospitalisation risks among unvaccinated and previously uninfected individuals, for low and high virulence variants**

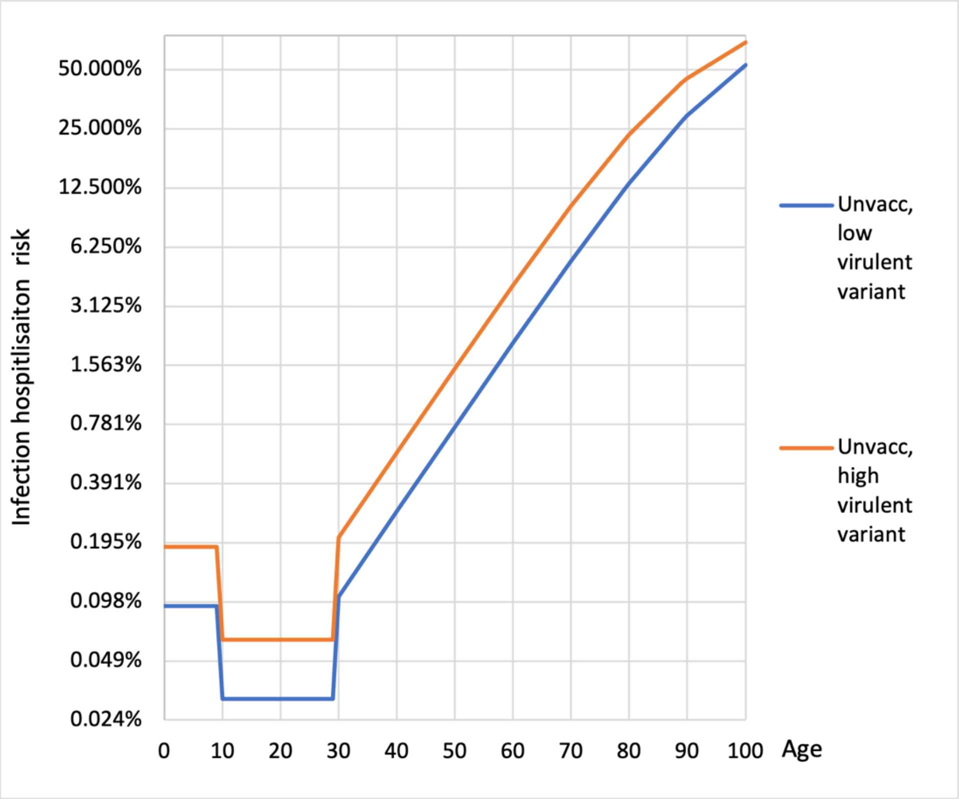

### Symptomatic infection risk

Estimates regarding the probability of being symptomatic given infection with SARS-CoV-2 were based on those from a meta-analysis using data through to April 2021.[21] These were then multiplied $2^{0.25}$ and $0.5^{0.25}$ to obtain estimates of the probably of symptomatic disease given infection on an odds scale for a more or less virulence variant respectively, to give the following (converted back to a percentage scale; uncertainty +/-10% SD on log odds scale for all values):

- Current Omicron and low-virulence new variant:
  - ≥60 years: Pr(symptomatic | inf) = 0.774
  - 20-59 years: Pr(symptomatic | inf) = 0.640
  - <20 years: Pr(symptomatic | inf) = 0.490
- High-virulence new variant:
  - ≥60 years: Pr(symptomatic | inf) = 0.829
  - 20-59 years: Pr(symptomatic | inf) = 0.716
  - <20 years: Pr(symptomatic | inf) = 0.576

### Length of hospital and ICU stay

Variant virulence impacted length of hospital and ICU stay as shown in Appendix Table 7, with estimates based on those for New South Wales, Australia, during periods of Delta and Omicron predominance.[22]

#### Appendix Table 7: Mean (and 95% confidence interval) length of stay associated with low and high virulence SARS-CoV-2 variants

| | Age (years) | Omicron/low virulence variant (LOS, days) | High virulence variant (LOS, days) |
|---|---|---|---|
| Hospital LOS | ≤39 | 2.05 (1.80 to 2.30) | 3.60 (3.48 to 3.81) |
| | 40-69 | 2.92 (2.50 to 3.67) | 5.78 (5.59 to 5.99) |
| | ≥70 | 6.02 (4.91 to 7.01) | 12.31 (11.75 to 12.95) |
| ICU LOS | ≤39 | 3.93 (2.58 to 5.68) | 7.50 (6.99 to 8.33) |
| | 40-69 | 4.30 (3.29 to 5.72) | 9.44 (8.81 to 10.07) |
| | ≥70 | 4.36 (3.40 to 5.57) | 8.94 (7.80 to 9.91) |

*LOS: length of stay. Uncertainty around these values parameterised in the model using a normal draw with a standard deviation based on the width of the reported 95% confidence interval divided by 3.92*

### Immune escape

SARS-CoV-2 is under strong evolutionary pressure to develop mutations that afford it the ability to escape pre-existing immunity, either obtained through vaccination or previous infection.[18] We reflected this in our model via ORs applied to agent immune protection at any given point in time (either vaccine- or natural infection-derived; see below for details regarding calculation of protection).

To understand our immune escape system, it helps to think in terms of:

1. the Omicron BA.2 variant that is dominant in our modelling on 1 April 2022
2. the Omicron BA.4/5 variant that is introduced to our model on 1 May 2022
3. and a variant introduced on 1 November 2022.

1. As the model is initiated and calibrated for BA.2, there is no immune escape. But an Omicron-targeted vaccine (e.g., mRNA.1273.214) will double on the odds ratio scale the vaccine effectiveness, and likewise (yet to be brought to market) next-generation multivalent vaccines. This is represented in the first row of Appendix Table 8.
2. Now consider the arrival of Omicron BA.4/5 (on 1 May 2022 in our model). Vaccine effectiveness of all vaccines (i.e., all columns in Appendix Table 8) is reduced by an 'immune escape odds ratio' of 0.5.
3. A novel variant (i.e., not yet seen) arrives on 1 November 2022. To have a survival advantage, it needs either low (defined in our model as an OR = 0.707) or high (defined in our model as OR = 0.5) immune escape on top of that already possessed by Omicron BA.4/5. Thus, the combined immune escape ORs are those in the second row multiplied by 0.707 and 0.5. The one exception to this is for the next-generation Omicron-targeted vaccine against an antigenically novel (i.e., non-Omicron-like) variant; we assume here that an Omicron-targeted vaccine has no better protection against this novel variant than current generation vaccines.

### Appendix Table 8: Immune escape odds ratios for vaccines cross-classified by variants in the model

| | | First generation double dose | First generation 3+ doses | Next-generation Omicron-targeted 1+ doses | Next-generation multivalent 1+ doses |
|---|---|---|---|---|---|
| | **Omicron BA.2** | 1 (ref)† | 1 (ref)‡ | *2* | *2* |
| | **Omicron BA.4/5** | *0.5* | *0.5* | *1* | *1* |
| **New variants** | **Omicron-like** | *0.354, 0.25* | *0.354, 0.25* | *0.707, 0.5* | *0.707, 0.5* |
| | **Novel** | *0.354, 0.25* | *0.354, 0.25* | *0.5, 0.354* | *0.707, 0.5* |

*†Reference for the current generation double dose column.*
*‡Reference for the current generation triple dose, next-generation omicron-targeted and next-generation multivalent columns.*
*These ORs apply at any point in time, with waning post last dose also in effect. All italicized ORs are drawn from a range of 0.841 to 1.189 of their stated values, with 100% correlation (including the OR of 2 for next-generation vaccines against current Omicron) uniformly on the log OR scale. For each combination of vaccine and new variant, 2 possible levels (low, high) of immune escape are modelled as indicated by the two ORs in each cell.*

### Appendix Table 9: Immune escape odds ratios for natural protection following primary infection cross-classified by variants in the model

| | | Previous infection with Omicron BA.2 | Previous infection with Omicron BA.4/5 | Previous infection with new variant |
|---|---|---|---|---|
| | **Omicron BA.2** | 1 (ref) | *1* | *1* |
| | **Omicron BA.4/5** | *0.5* | *1* | *1* |
| **New variants** | **Omicron-like** | *0.354, 0.25* | *0.707, 0.5* | *1* |
| | **Novel** | *0.25, 0.177* | *0.5, 0.354* | *1* |

*These ORs apply at any point in time, with waning post previous infection also in effect. All italicized ORs are drawn uniformly from a range of 0.841 to 1.189 of their stated values, with 100% correlation, uniformly on the log OR scale. For new variants, two levels of possible immune escape are modelled.*

#### Vaccine effectiveness and protection against reinfection

##### *Vaccine effectiveness*

The effectiveness of COVID-19 vaccines peaks soon after receipt of a full vaccination course and wanes in a non-linear fashion thereafter.[23-25] To dynamically model waning of vaccine-derived immunity for the outcomes of symptomatic infection and hospitalisation from COVID-19, we developed a log-odds system of equations that has been published elsewhere[5] building on VE data published by the UK Health Security Agency (UKHSA).[23,26] In summary, a random effects logistic regression model with a separate class for each combination of vaccine course and outcome was fitted to this data, excluding observations within 2 weeks since last vaccine dose (given peak immunity would still have been developing) and weighted by the inverse variance as follows:

$$logit(VE) = \alpha + \beta_1 Hosp + \beta_2 Triple + \beta_3 Month + \beta_4 Hosp * Month + \beta_5 Triple * Month$$

where:

VE is vaccine effectiveness;

$\alpha$ is the intercept, or the logit of the VE for the reference case (VE against symptomatic illness two weeks after double dose vaccine);

*Hosp* is a dummy variable for hospitalisation versus symptomatic illness;

*Triple* is a dummy variable for triple versus double vaccine course;

*Month* represents months (continuous) since last vaccine dose minus 0.5 (i.e., 'centered' on two weeks post the last vaccine dose to aid interpretation of model coefficients);

*Hosp*Month* is an interaction term of hospitalisation and month (i.e., how VE wanes differently on the ln odds scale for protection against hospitalisation as compared to protection against symptomatic illness); and

*Triple*Month* is an interaction term of triple and month.

For models including interaction terms of *hosp*triple*month* and *triple*hosp* these coefficients had non-significant p values, and the differences in deviance statistics between these models and that without these interaction terms were trivial and non-significant. As such they were not retained in the final model.

The UKHSA data used to develop the random effects logistic regression model described above did not include VE estimates stratified by age. However, there is evidence that VE varies by age.[24] To alter the regression equation to reflect this, we focused on estimates at 10-14 weeks post-vaccination. Using data from a study reporting VE against the Delta variant stratified by age group (≥16 years, 16-39 years, 40-64 years, ≥65 years) and vaccine type (ChAdOx1-S or BNT162b2)[24] we specified the following:

- The VE OR for ≥65-year-olds compared to <65-year-olds for BNT162b2 VE against symptomatic disease was 0.77 (at 10-14 weeks after receipt of a second dose).
- The VE OR for ≥65-year-olds compared to <65-year-olds for BNT162b2 VE against hospitalization was 0.36 (at 10-14 weeks after receipt of a second dose).
- The proportion of symptomatic cases in the <65-year-old group was 94%, and the proportion of hospitalisations in the <65-year-old group was 70.2%.

To fit these data, the above logistic regression model was re-estimated to preserve VE at 3 months post-vaccination, by solving for a new intercept. Further adjustments were then made to generate coefficients split by 3 age strata; <60 years, 60-69 years, and ≥70 years (to fit the 10-year age strata specified in our ABM).

Next, we extended the above logistic regression model to account for the outcomes of any infection (including asymptomatic infection), ICU admission and death. The UKHSA reports VE (all vaccines combined) for mortality against the Omicron variant for those aged ≥50 years to be 95% at ≥2 weeks following receipt of a third dose.[27] By combining this estimate with their reported OR for symptomatic disease ≥2 weeks after 3 doses for the same age group (0.41) and the associated 95% confidence intervals we drew 5,000 estimates of the odds VE of mortality v. symptomatic disease, the median of which was used to estimate a coefficient for the outcome of death in the overall VE logistic regression equation. Whilst these input data were for ≥50-year-olds who were triple-dose vaccinated, given we do not allow for interactions of age with severity and triple dose with severity we could use estimates at any age to estimate the ‘independent’ or ‘main’ effect for VE protection against death.

Limited data is available to inform estimates of VE against any infection with the Omicron variant (i.e., asymptomatic and symptomatic infection combined). As such, data from the UK Vaccine Effectiveness Expert Panel[27] were used to estimate the odds ratio for VE for all infections compared to the VE for symptomatic infection at 3 months (for BNT162b2 and mRNA-1273) and obtain the coefficient for any infection. Specifically, for each of the four combinations of BNT162b2 and mRNA-1273, for two and three doses:

- the values reported in Table 3 of the UKHSA Week 13 COVID-19 Vaccine Surveillance Report[28] (VE as a percentage with a range both for any infection and symptomatic infection) were converted to logit(VE) with mean = ln odds of central VE, and SD = range/3.92 (approximating the 95% uncertainty interval).
- 5,000 draws on the ln odds scale were taken of each of ‘any infection’ and ‘symptomatic infection’, with 0.5 correlation between the two values.
- The median and 2.5$^{th}$ and 97.5$^{th}$ percentiles of the ln odds VE were determined.

The final coefficient for any infection was the average of the four ln odds VE values (-0.78), and the final SD (on the ln odds scale) was the average of the SD (assumed as 97.5$^{th}$ percentile – 2.5$^{th}$ percentile, divided by 3.92; 0.27). For the outcome of ICU admission, the coefficient was estimated as the mid-point between the coefficients for hospitalization and death, with the same standard deviation as that for death. The final VE equation utilized in the ABM was therefore defined as follows:

$$logit(VE) = \alpha + \beta_0 Age + \beta_1 Severity_j + \beta_2 Triple + \beta_3 Month + \beta_4 Severity * Month + \beta_5 Triple * Month$$

where:

$\alpha$ is the intercept or the logit of the VE for the reference case (<60 years old, two doses of vaccine, against symptomatic infection, 2 weeks after the last dose);
*Age* is a categorical variable of ≥70 years, 60-69 years, or <60 years (ref);
*Severity* is categorical value of any infection, symptomatic infection (ref), hospitalisation, ICU admission or death;
*Triple* is a dummy variable for triple versus double vaccine course;

*Month* represents months (continuous) since last vaccine dose minus 0.5 (i.e., 'centered' on two weeks post the last vaccine dose to aid interpretation of model coefficients);

*Severity*Month* is an interaction term of severity and month; and

*Triple*Month* is an interaction term of triple and month.

Coefficients for this VE system are shown in Appendix Table 10. Footnotes to the table describe how values were sampled for each draw (then taken into the 500 iterations of each scenario in the ABM). Note we assumed that waning (on the log odds scale) was the same for any and symptomatic infection, and the same for hospitalisation, ICU and death. We also assumed that peak effectiveness and waning for a third dose vaccine applied also to any subsequent vaccine doses.[29,30] As of 1 April 2022, Omicron BA.2 was assumed to be responsible for all SARS-CoV-2 infections in Victoria – estimates of VE against BA.1 appear to continue to apply for BA.2 and as such VE was not adjusted in response to BA.2 emergence.[27,31,32] See Appendix Figure 4 for an example of VE estimates against the Omicron variant for <60-year-olds by month following second and third vaccine doses, and Appendix Figure 5 for an example of VE estimates against hospitalisation due to the Omicron variant by month since vaccination for double and triple doses in three age strata.

**Appendix Table 10: Vaccine effectiveness log odds equation coefficients for the Omicron variant**

| **Coefficients** | **Central** | **SD** | **Odds (%)** | **95% UI odds** |
|---|---|---|---|---|
| Intercept[†] | 0.75 | 0.12 | 2.27 (68.0%) | 1.67 to 2.70 |
| | | | **OR** | **95% UI** |
| Age ≥70 years[£] | -0.26 | 0.12 | 0.77 | 0.61 to 0.98 |
| Age 60-69 years[‡] | -0.13 | 0.061 | 0.88 | 0.78 to 0.99 |
| Age <60 years (ref) | 0 | N/A | N/A | N/A |
| Severity_Any[‡] | -0.78 | 0.27 | 0.46 | 0.27 to 0.77 |
| Severity_Sympt (ref) | 0 | N/A | N/A | N/A |
| Severity_Hosp[†] | 1.13 | 0.26 | 3.11 | 1.87 to 5.16 |
| Severity_ICU[‡] | 1.87 | 0.37 | 6.47 | 13.14 to 13.40 |
| Severity_Death[‡] | 2.60 | 0.37 | 13.46 | 6.52 to 27.80 |
| ≥3Doses[†] | 0.092 | 0.14 | 1.10 | 0.83 to 1.45 |
| 2 Doses (ref) | 0 | N/A | N/A | N/A |
| Month (continuous, centered on 0.5 months after last dose)[†] | -0.64 | 0.029 | 0.53 | 0.50 to 0.56 |
| Months*Severity_Any | 0 | N/A | N/A | N/A |
| Months*Severity_Sympt (ref) | 0 | N/A | N/A | N/A |
| Months*Severity_Hosp[†] | 0.22 | 0.095 | 1.24 | 1.03 to 1.50 |
| Months*Severity_ICU | 0.22 | 0.095 | 1.24 | 1.03 to 1.50 |
| Months*Severity_Death | 0.22 | 0.095 | 1.24 | 1.03 to 1.50 |
| Months*≥3Doses[†] | 0.21 | 0.033 | 1.23 | 1.16 to 1.32 |

*[†]Drawn from covariance matrix accompanying regression model on UKHSA data.[5]*
*[‡]Correlated 1.0 with the value in this domain drawn from covariance matrix accompanying regression model on UKHSA data.[5] E.g., if the value drawn for Severity_Hosp was at the 63%ile of its distribution, then all other values for Severity_Any, Severity_ICU and Severity_Death are allocated the 63%ile of their distribution.*
*[£]The value for age was drawn independently but correlated 1.0 for ≥ 70 years and 60-69 years.*

#### Appendix Figure 4: Estimated vaccine effectiveness against the Omicron variant by month following a second and third vaccine dose, age <60 years

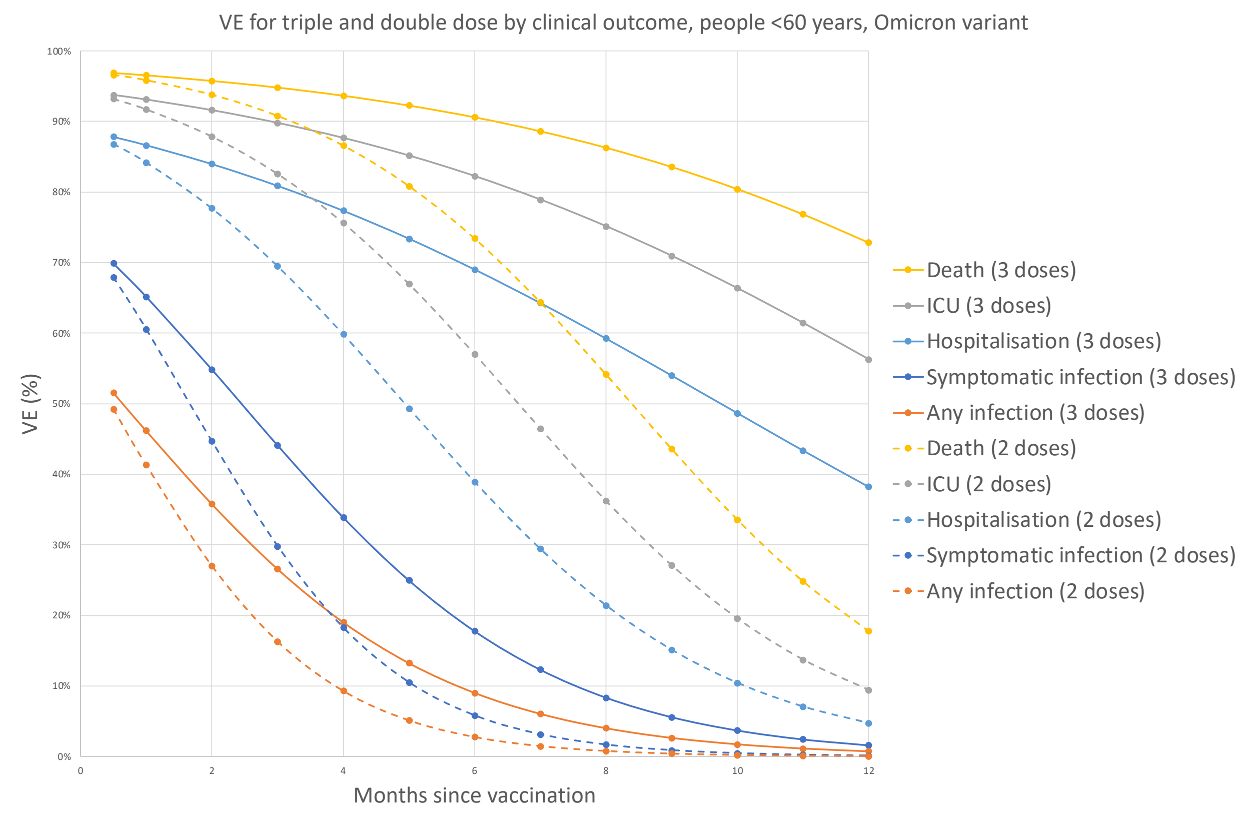

**Appendix Figure 5: Estimated vaccine effectiveness against hospitalisation due to the Omicron variant by month since vaccination for double and triple dose in three age strata**

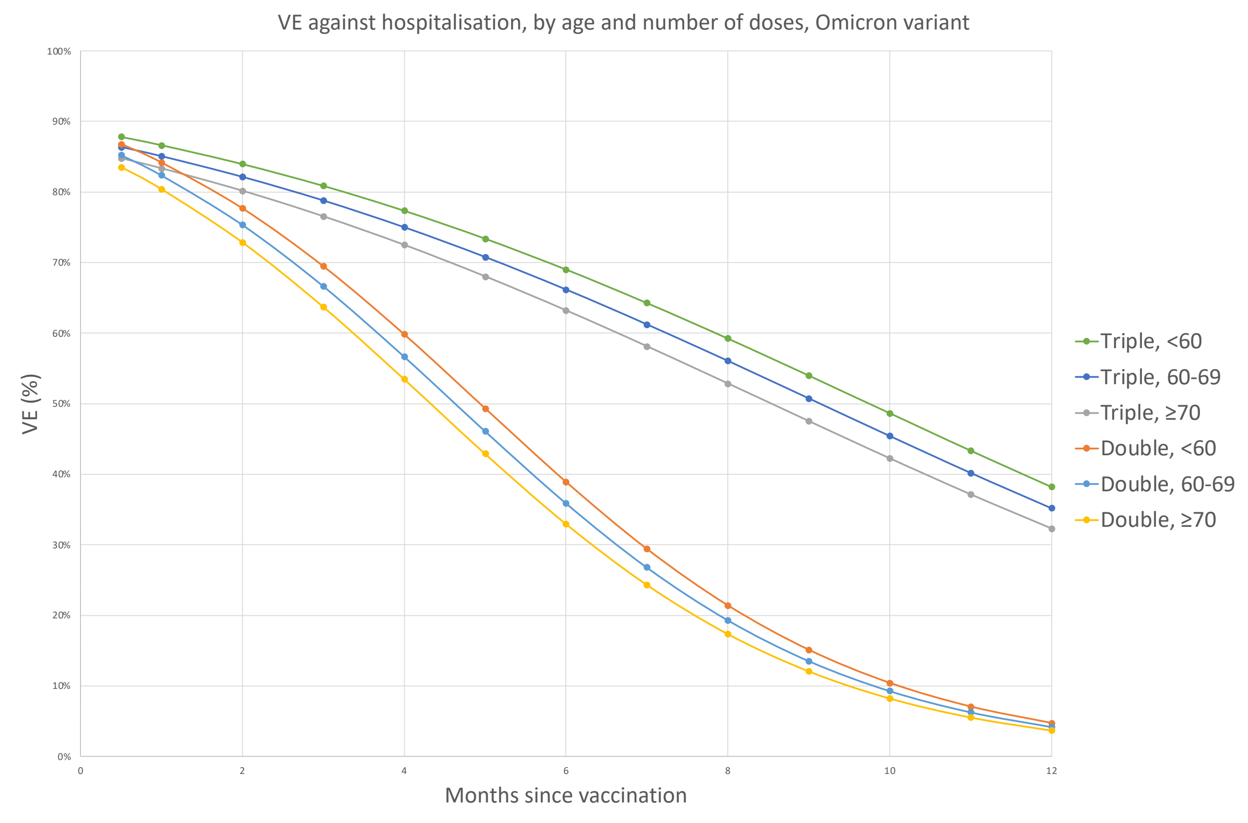

#### *Protection from reinfection*

Evidence indicates that previous natural infection provides considerable protection against reinfection (and subsequent disease if infected) with SARS-CoV-2. To model agent-level protection over time following primary SARS-CoV-2 infection in our model, we extracted data from studies reporting protection against any infection and symptomatic infection following infection and re-exposure both from pre-Omicron variants from Denmark[33] and protection against infection following infection and re-exposure both from Omicron variants from Canada.[34] We fit these data to a logistic regression model with percentage protection as the outcome and months since primary infection, variant responsible for primary infection and re-exposure (either pre-Omicron or Omicron) and clinical outcome (any infection v. symptomatic infection) as explanatory variables. Tests for interaction between these terms were non-significant and as such no interaction terms were retained in the final model.

We compared the level of protection against hospitalization following primary infection to that of any infection as reported by Michlmayr et al. to estimate the increase in protection for this more severe outcome and added a coefficient to the regression model accordingly. Finally, given the paucity of comparable data reporting protection against ICU admission and mortality following primary infection, we added coefficients for these clinical outcomes using the relative increases in protection against severe disease in our VE regression model described above. SDs for the intercept, time since primary infection, Omicron/pre-Omicron infection and re-exposure, and symptomatic v. any infection were derived from the regression output. For the additional clinical outcomes of hospitalization, ICU admission and death, SDs were derived from the absolute value of 20% of the regression coefficient given inherent uncertainty in these estimates.

Immune responses to SARS-CoV-2 vary by age, with reduced protection following infection observed in older age groups. As such, we used age-stratified estimates of protection following infection reported by Michlmayr et al.[33] to add an additional regression coefficient for age (<65/≥65). This was done by splitting the all-ages estimate into

<65/≥65 estimates, based on contributions of person-time per age group and age-specific estimates of protection from Michlmayr et al., converting these to an OR and taking the natural logarithm to generate a regression coefficient. The coefficient for the intercept of the model was also adjusted accordingly.

Draws to parameterise the model were then randomly generated using the covariance matrix from the regression output for those terms in the base model (intercept, time in months, Omicron/pre-Omicron, symptomatic v. any infection) and randomly generated independently for other terms based on their central estimate and SD.

The final natural protection equation utilized in the ABM was therefore defined as follows:

$$logit(natural\ protection) = \alpha + \beta_0 Age + \beta_1 Severity_j + \beta_2 Variant + \beta_3 Month$$

where:

$\alpha$ is the intercept or the logit of natural protection for the reference case (<65 years old, against any infection, with a pre-Omicron variant responsible for primary infection and re-exposure);
*Age* is a categorical variable of ≥65 or <65 years (ref);
*Severity* is categorical value of any infection (ref), symptomatic infection, hospitalisation, ICU admission or death;
*Variant* is a dummy variable representing whether a pre-Omicron (ref) or Omicron variant was responsible for the primary infection and re-exposure; and

*Month* represents months (continuous) since primary infection

Note these apply to unvaccinated individuals and are further adjusted as described below for vaccinated individuals.

See Appendix Figure 6 for an illustration of reinfection protection estimates following primary infection with the Omicron BA.1/2 variant (upon re-exposure to the same variant).

**Appendix Table 11: Natural protection log odds equation coefficients for reinfection with the same variant as the primary infection**

| Coefficients | Central | SD |
|---|---|---|
| Intercept | 2.88 | 0.25 |
| Age ≥65 years | -0.77 | 0.15 |
| Age <65 years (ref) | 0 | N/A |
| Severity_Any (ref) | 0 | N/A |
| Severity_Sympt | 0.27 | 0.15 |
| Severity_Hosp | 0.48 | 0.10 |
| Severity_ICU | 0.78 | 0.16 |
| Severity_Death | 0.99 | 0.20 |
| Month (continuous) | -0.16 | 0.03 |
| Pre-omicron variant (ref) | 0 | N/A |
| Omicron variant | -1.26 | 0.39 |

**Appendix Figure 6: Estimated protection against any reinfection by month since primary infection, unvaccinated (Omicron BA.1/2 primary infection and re-exposure)**

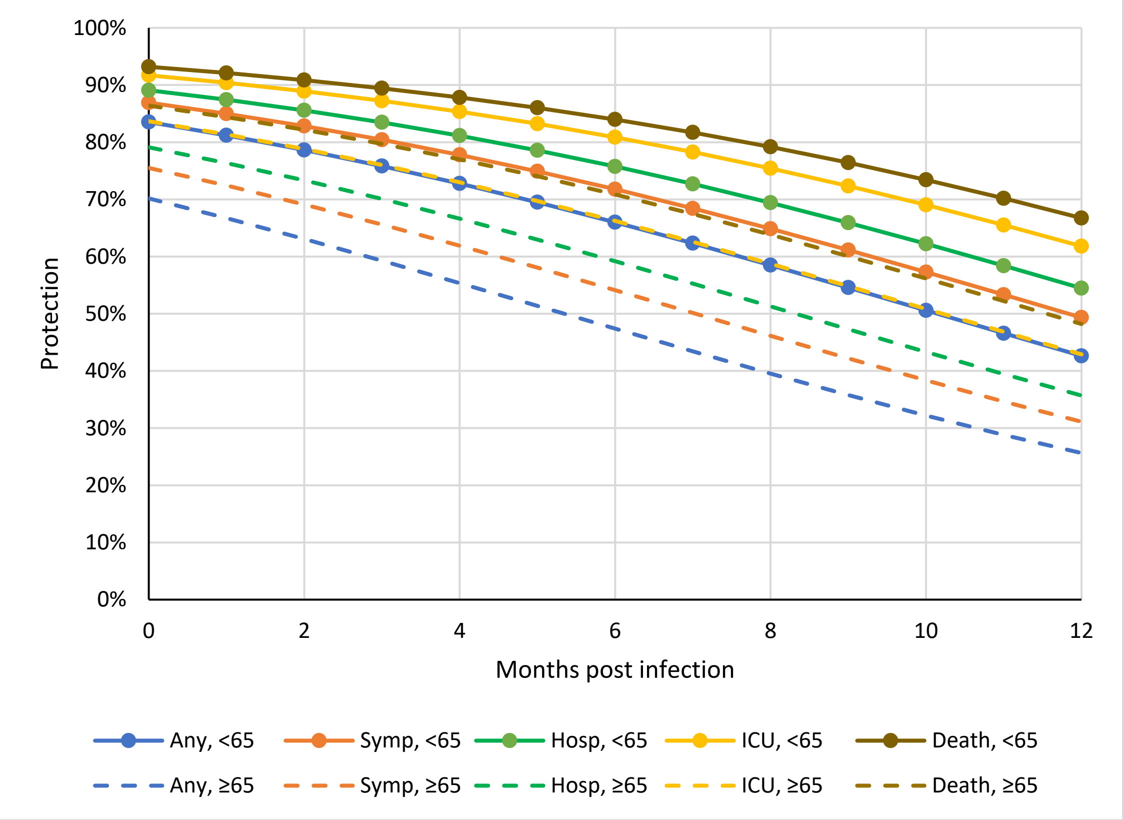

#### *Hybrid immunity*

Immunity provided by the combination of vaccination and natural infection (hybrid immunity) has been quantified by, for example, studies conducted in Israel[35,36] a large population-level study in Sweden,[37] and a study in Qatar.[32] In both Israeli studies, protection against reinfection was assessed among previously infected individuals either receiving a single dose of the BNT162b2 mRNA vaccine or remaining unvaccinated, and the hazard ratio for reinfection among individuals both infected and subsequently vaccinated was 0.18 compared to those previously infected and unvaccinated. In the study from Sweden, hybrid immunity was found to provide additional protection above natural immunity for both reinfection and hospitalisation risk,[37] while the study from Qatar also demonstrated that hybrid immunity provides greater protection than vaccination or natural infection alone.[32] The kinetics of hybrid immunity are complex however, owing to variability in terms of timing of infection and vaccination (and the time elapsed between these two sensitizing events) in addition to consideration of dominant circulating variants, for example, and several potential sources of bias in estimates.[37] Current evidence suggests that the two types of immunity may act independently to confer an overall combined level of protection;[32] accordingly we calculated protection for an individual agent at any given point in time as $1 - (1 - VE) \times (1 - \text{protection from prior infection})$.

#### *The effect of immunity on onward transmission*

Data quantifying the effect of immunity (either from vaccination or previous infection) on ongoing transmission if one becomes infected is scarce. Guided by evidence of reduced transmission for Omicron following vaccination in Danish households[38] and evidence that this reduced transmission does wane over time[39] we assumed that the reduction in ability to transmit the virus if infected following vaccination or previous infection follows the same function as VE and/or natural protection against any infection (and wanes accordingly). Thus if the VE at time t against any infection is 20%, then if infected that agent also has 20% less chance of transmitting the virus on compared to if they had been infected with no previous vaccination.

### Morbidity and mortality estimates

#### *Acute COVID-19 morbidity*

We estimated the number of asymptomatic, non-hospitalised symptomatic, hospitalised symptomatic, and ICU-admitted cases for the quantification of acute COVID-19 morbidity, and deaths for the quantification of HALYs lost due to death from COVID-19. We measured morbidity impacts using disability rates (DR), according to the severity of acute infection (cases managed in the community, being either mild or moderate, and cases requiring hospitalisation, either managed in ward only or additionally including ICU admission) using estimates from the Global Burden of Disease (GBD) study.[40] For ICU admissions, the DR was based on the GBD disability weight (DW) for 'severe chronic obstructive pulmonary disease (COPD) and other respiratory infections', to reflect acute respiratory distress syndrome (ARDS). The GBD does not have an estimate for ARDS directly – the estimate used here is the GBD's most severe rating for respiratory infections.[40] We assumed that survivors return to their baseline health status (DR = 0) following a specified recovery period as described below. This does not apply to those who go on to develop symptoms of long COVID – morbidity associated with long COVID was quantified separately.

Estimates are presented separately for high virulence variant infections (using data relating to the Delta variant), and low virulence variant infections (using data relating to the Omicron variant). The duration of symptoms for the purpose of morbidity calculations changes for low virulence variant infections compared to a high virulence variant, however the DRs applied are the same for both. The duration of symptoms for low virulence variants and high virulence variants was obtained from a large ongoing UK-based study, comparing acute illness parameters for Omicron vs. Delta infections.[41] In this study, the duration of symptomatic infection was not stratified by hospitalisation status – as such, estimates relating to cases managed in the community are for those with infection of duration 21 days or less.[41] For duration of symptoms for hospitalised patient sub-groups, the length of stay defined in Appendix Table 7 are applied (including either hospital stay only, or hospital stay and ICU stay).

Morbidity loss for the four categories of symptomatic SARS-CoV-2 infection with a high virulence variant (Appendix Table 12) was calculated as follows:

- Morbidity for symptomatic, but not hospitalised, infections were split evenly as 50% moderate infection, with a DR of 0.051 (95% CI 0.032-0.074), and 50% mild infection, with a DR of 0.006 (95% CI 0.002-0.012), both with a mean duration of 8.89 days (95% CI 8.61-9.17).[41]
- Morbidity for hospitalised patients not requiring ICU admission assumes a period of 1 week prior to hospitalisation with moderate acute infection, with a DR of 0.051 (95% CI 0.032-0.074), followed by a duration of hospital stay varying by age (as per Appendix Table 7), all with a DR of 0.133 (95% CI 0.088-0.190) for severe acute infection. A duration of 1-week post-hospitalisation for return to baseline health was applied, with a DR reducing linearly to zero from the severe acute infection DR.
- Morbidity for hospitalised patients requiring ICU admission assumes a period of 1 week prior to hospitalisation with moderate acute infection, with a DR of 0.051 (95% CI 0.032-0.074), followed by a duration of stay in hospital (on a general ward) varying by age (as per Appendix Table 7), all with a DR of 0.133 (95% CI 0.088-0.190) for severe acute infection. This is followed by duration of stay in ICU as per Appendix Table , all with a DR of 0.408 (95% CI 0.273-0.556) to approximate ARDS. A duration of 2 weeks for return to baseline health was applied, with a DR reducing linearly to zero from the ARDS DR.

Morbidity loss for the four categories of symptomatic SARS-CoV-2 infection by a low-virulence variant (Appendix Table 13) was calculated as follows:

- The proportion of mild/moderate symptomatic, but not-hospitalised, infections were altered compared to high-virulence variant infections: 28% have moderate infection, with a DR of 0.051 (95% CI 0.032-0.074), and 72% have mild infection, with a DR of 0.006 (95% CI 0.002-0.012), both with duration of 6.87 days (95% CI 6.58-7.16). The proportion with mild vs. moderate severity of infection was estimated from the aforementioned UK-based study, which found the odds of 'classic' acute COVID-19 symptoms were reduced by 44% for Omicron compared to Delta infections.[41]
- Morbidity for hospitalised patients not requiring ICU assumes a period of 1 week prior to hospitalisation with moderate acute infection, applying a DR of 0.051 (95% CI 0.032-0.074), followed by a duration of stay varying by age as per Appendix Table 7, all with DR of 0.133 (95% CI 0.088-0.190) for severe acute infection. A duration of 1-week post-hospitalisation for return to baseline health was applied, with a DR reducing linearly to zero from the severe acute infection DW.

- Morbidity for hospitalised patients requiring ICU admission assumes a period of 1 week prior to hospitalisation with moderate acute infection, with a DR of 0.051 (95% CI 0.032-0.074), followed by a duration of stay in hospital (on a general ward) varying by age as per Appendix Table 7, all with DR of 0.133 (95% CI 0.088-0.190) for severe acute infection. This is followed by duration of stay in ICU as per Appendix Table 7, all with DR of 0.408 (95% CI 0.273-0.556). A duration of 2 weeks for a post-hospitalisation for return to baseline health was applied, with the DW reducing linearly to zero from the ARDS DR.

The overall morbidity for each clinical infection category was calculated as the sum of the symptom severity-specific DR multiplied by the symptom duration (in years). Note these are not additive.

#### Appendix Table 12: Disability rate distribution and duration by SARS-CoV-2 category, high virulence variant

| Category | Symptom severity (health state) | Duration in days, mean (95% CI) | Disability Rate†, mean (95% CI) |
|---|---|---|---|
| Admitted to ICU | Moderate acute infection | 7 | 0.051 (0.032-0.074) |
| | Severe acute infection (ward) | Appendix Table 7 | 0.133 (0.088–0.190) |
| | In-ICU (COPD for ARDS) | Appendix Table 7 | 0.408 (0.273-0.556) |
| | Return to baseline health | 14 | Linearly to 0 from ARDS DR |
| Admitted to hospital, no ICU | Moderate acute infection | 7 | 0.051 (0.032-0.074) |
| | Severe acute infection (ward) | Appendix Table 7 | 0.133 (0.088–0.190) |
| | Return to baseline health | 7 | Linearly to 0 from severe acute infection DWR |
| Symptomatic, not admitted | 50% Moderate acute infection | 8.89 (8.61-9.17) | 0.051 (0.032-0.074) |
| | 50% Mild acute infection | 8.89 (8.61-9.17) | 0.006 (0.002–0.012) |

*†Disability rate applied based on DWs for equivalent health states from [40]*

#### Appendix Table 13: Disability rate distribution and duration by SARS-CoV-2 category, low virulence variant

| Category | Symptom severity (health state) | Duration in days, mean (95% CI) | Disability Rate†, mean (95% CI) |
|---|---|---|---|
| Admitted to ICU | Moderate acute infection | 7 | 0.051 (0.032-0.074) |
| | Severe acute infection (ward) | Appendix Table 7 | 0.133 (0.088–0.190) |
| | In-ICU (COPD for ARDS) | Appendix Table 7 | 0.408 (0.273-0.556) |
| | Return to baseline health | 14 | Linearly to 0 from ARDS DR |
| Admitted to hospital, no ICU | Moderate acute infection | 7 | 0.051 (0.032-0.074) |
| | Severe acute infection (ward) | Appendix Table 7 | 0.133 (0.088–0.190) |
| | Return to baseline health | 7 | Linearly to 0 from severe acute infection DR |
| Symptomatic, not admitted | 28% Moderate acute infection | 6.87 (6.58-7.16) | 0.051 (0.032-0.074) |
| | 72% Mild acute infection | 6.87 (6.58-7.16) | 0.006 (0.002–0.012) |

*†Disability rate applied based on DWs for equivalent health states from [40]*

Given the above, the following HALY loss was applied to each agent based on their clinical outcome category (given the SDs of the inputs approximate 20% of their means, we included uncertainty here by applying a multiplier of 1 with SD 0.2 to acute HALYs generated by the model):

- Current Omicron and low-virulence new variant:
  - For an infected, asymptomatic agent: 0
  - For an infected, symptomatic agent: 0.000350087671
  - For a hospitalised agent: 0.000978082 + ((length of hospital stay in days/365)*0.133) + 0.001275342
  - For an ICU admitted agent: 0.000978082 + ((length of hospital stay in days/365)*0.133) + ((length of ICU stay in days/365)*0.408) + 0.007824658
- High-virulence new variant:

- For an infected, asymptomatic agent: 0
- For an infected, symptomatic agent: 0.000694150685
- For a hospitalised agent: 0.000978082 + ((length of hospital stay in days/365)*0.133) + 0.001275342
- For an ICU admitted agent: 0.000978082 + ((length of hospital stay in days/365)*0.133) + ((length of ICU stay in days/365)*0.408) + 0.007824658

#### *Long COVID morbidity*

Ongoing symptoms following acute SARS-CoV-2 infection can be grouped into two phases: the 'acute post-COVID symptoms' or 'ongoing symptomatic' phase (up to 12 weeks post-infection) and 'long COVID' phase (≥12 weeks post-infection).[42,43] Long COVID, formally termed 'Post-Acute Sequelae of SARS-CoV-2' and defined in ICD-10 classification as 'Post-COVID-19 Condition', describes the persistence and/or emergence of a heterogeneous group of physical and cognitive symptoms following acute SARS-CoV-2 infection.[43,44] The exact mechanisms and symptom profile of long COVID are yet to be fully understood. Comparisons have been made with chronic fatigue syndrome, as fatigue, muscle weakness, and concentration difficulty (or 'brain fog') have been some of the most reported symptoms.[45,46] However, other notable symptoms unique to long COVID include an impaired sense of taste (dysgeusia)/smell (dysosmia) and shortness of breath (dyspnoea).[47]

For the purposes of this model a bottom-up approach was utilized to estimate long COVID morbidity, calculating a morbidity estimate individually for each major long COVID symptom as the product of its DR duration and prevalence among acute COVID-19 survivors. The DR for each symptom has been estimated from the GBD study DWs.[40] Some long COVID symptoms do not have direct health states for which DWs exist – in these cases best estimates have been determined from related health states. For example, the DWs of other sensory conditions (mild hearing loss and mild visual impairment) have been used as proxies for the DWs of dysosmia and dysgeusia (the full list of DWs applied are presented in Appendix Table 15).

Symptom prevalence estimates were drawn from risk differences between COVID-19 cases and COVID-negative controls from large international studies. Four variables were identified from the available literature with which to stratify long COVID estimates: age (adult vs. child), severity of acute infection (approximated for this model as hospitalised vs. community-managed cases), vaccination (at least 2 doses vs. less than 2, with waning not considered), and SARS-CoV-2 variant (high vs. low virulence variant). The 'base cases' for which prevalence estimates were available in the literature are adults/children infected with a pre-Omicron variant of SARS-CoV-2 (i.e. 'high virulence' group), who are not vaccinated at the time of infection.

Symptom prevalence was drawn from four studies:

- A case-control study by Vedel Sørensen et al. conducted in Denmark: used to obtain prevalence of physical long COVID symptoms, for adult patients diagnosed with acute COVID-19 6-12 months previously, compared to PCR-negative/seronegative controls, separately for individuals hospitalised vs. non-hospitalised during the acute disease period.[48]
- A case-control study by Caspersen et al. based in Norway: used to obtain prevalence of cognitive long COVID symptoms for adult patients either 1 to 6 or 11 to 12 months post-infection. Patients were stratified into 'mild' vs. 'moderate/severe' cases, with moderate cases including those not hospitalised – weighting was therefore applied to the prevalence estimates for these groups to achieve the risk ratio (RR) for any long COVID symptoms between hospitalised and non-hospitalised patients found by Vedel Sørensen et al. (RR=2) and approximate the prevalence of symptoms for previously hospitalised vs. non-hospitalised groups.[48,49]
- A multi-country case-control study by Magnúsdóttir et al.: used to obtain the prevalence of psychological long COVID symptoms (insomnia, anxiety and depression), stratified by hospitalised vs. community adult cases. Insomnia occurred at a greater prevalence for cases compared to controls for hospitalised and non-hospitalised groups, however, anxiety and depression only occurred more frequently in hospitalised cases compared to controls (not for community cases vs. controls).[50]
- A systematic review by Behnood et al.: used to obtain prevalence estimates for children (age 0-17), for whom there is estimated to be a much lower risk of ongoing symptoms.[51] The meta-analysis from this review combined data from 5 large, controlled studies undertaken in children. Sub-groups by

hospitalisation/severity are not considered for children, given the lower risk of hospitalisation compared to adults.

Base case prevalence estimates were then adjusted to account for vaccination, and low virulence variants of SARS-CoV-2. The prevalence of each symptom is multiplied by an OR of 0.55 for those who were vaccinated (both adults and children), given findings from large population-based studies by Antonelli et. al. and the Office for National Statistics in the UK, indicating reduced odds of long COVID symptoms for those vaccinated, compared to those unvaccinated.[52,53] Separately, the prevalence of each symptom was multiplied by an OR of 0.25, based on findings from a recent UK-based case-control study showing a reduced odds of any ongoing symptoms at least 4 weeks post-infection for Omicron variant infections compared to Delta variant infections.[54] It is assumed that this reduced symptom prevalence will continue into the long COVID period at least 12 weeks post infection.

A multivariate method was applied to account for the joint probability of having two long COVID symptoms, by multiplying the prevalence of each combination of two symptoms, then using multiplicative adjustment to combine DWs for such comorbidity.[55] The probability of having three or more symptoms was assumed to be low, and therefore only the joint probability of two symptoms was included. The following formulae were used for multiplicative adjustment:

$$Combined\ prevalence = Frequency_{Symptom\ A} \times Frequency_{Symptom\ B}$$

$$Combined\ DW = 1 - (1 - DW_{Symptom\ A}) \times (1 - DW_{Symptom\ B})$$

The duration of each symptom was estimated based on a recent analysis by Wulf Hanson et. al., which found a median duration of long COVID symptoms across 10 cohort studies of 3.99 months (interquartile range [IQR] 3.84-4.20) for community cases, and 8.84 months (IQR 8.10-9.78) for previously hospitalised cases.[56] For community cases, a linear decline in the DW applied at onset to full health at 8 months (DR = 0) was applied. For previously hospitalised cases, the given DW was applied to 6 months post infection, followed by a linear decline to 12 months (DR = 0). Exceptions to this were psychological symptoms, for which a shorter duration has been documented – for these symptoms, a linear decline in the DW from onset to 6 months post-infection was applied for both sub-groups.[50] Additionally, as symptoms in children resolve in a much shorter time frame compared to adults, a time frame of 3 months post-infection was applied for long COVID symptoms in children, followed by immediate return to full health (DR = 0).[57,58] Each time frame applied excludes the duration of acute infection, but includes the ongoing symptomatic phase in addition to the long COVID period.[42] No difference in duration is applied for low vs. high virulence variants, given the lack of data available to inform this. Duration and prevalence estimates for each symptom, across relevant patient sub-groups, are shown in Appendix Table 14.

#### Appendix Table 14: Long COVID symptom prevalence and duration estimates

| **Symptom** | **Prevalence**† | | **Duration** |
|---|---|---|---|
| *Adult community cases* | | | |
| | *High virulence* | *Low virulence* | |
| Dysosmia | 10.4% | 2.6% | Linear decline from onset (1-week post-infection) to 8 months |
| Dysgeusia | 8.2% | 2.1% | Linear decline from onset (1-week post-infection) to 8 months |
| Fatigue | 7.8% | 2.0% | Linear decline from onset (1-week post-infection) to 8 months |
| Dyspnoea | 4.2% | 1.1% | Linear decline from onset (1-week post-infection) to 8 months |
| Chest pain | 1.8% | 0.5% | Linear decline from onset (1-week post-infection) to 8 months |
| Muscle weakness | 4.1% | 1.0% | Linear decline from onset (1-week post-infection) to 8 months |
| Dizziness | 2.3% | 0.6% | Linear decline from onset (1-week post-infection) to 8 months |
| Muscle/joint pain | 3.2% | 0.8% | Linear decline from onset (1-week post-infection) to 8 months |
| Headache | 3.0% | 0.8% | Linear decline from onset (1-week post-infection) to 8 months |
| Numb limbs | 3.2% | 0.8% | Linear decline from onset (1-week post-infection) to 8 months |
| Concentration difficulty* | 7.6% | 1.9% | Linear decline from onset (1-week post-infection) to 8 months |
| Memory impairment* | 5.5% | 1.4% | Linear decline from onset (1-week post-infection) to 8 months |
| Insomnia | 5.3% | 1.3% | Linear decline from onset (1-week post-infection) to 8 months |
| *Adult hospitalised cases*# | | | |

| | | | |
|---|---|---|---|
| Dysosmia | 7.0% | | 6 months from onset (4 weeks post-infection), followed by linear decline to 12 months |
| Dysgeusia | 6.9% | | 6 months from onset (4 weeks post-infection), followed by linear decline to 12 months |
| Fatigue | 13.1% | | 6 months from onset (4 weeks post-infection), followed by linear decline to 12 months |
| Dyspnoea | 10.3% | | 6 months from onset (4 weeks post-infection), followed by linear decline to 12 months |
| Chest pain | 4.1% | | 6 months from onset (4 weeks post-infection), followed by linear decline to 12 months |
| Muscle weakness | 11.3% | | 6 months from onset (4 weeks post-infection), followed by linear decline to 12 months |
| Dizziness | 4.8% | | 6 months from onset (4 weeks post-infection), followed by linear decline to 12 months |
| Muscle/joint pain | 6.3% | | 6 months from onset (4 weeks post-infection), followed by linear decline to 12 months |
| Headache | 4.3% | | 6 months from onset (4 weeks post-infection), followed by linear decline to 12 months |
| Numb limbs | 7.0% | | 6 months from onset (4 weeks post-infection), followed by linear decline to 12 months |
| Concentration difficulty* | 10.9% | | 6 months from onset (4 weeks post-infection), followed by linear decline to 12 months |
| Memory impairment* | 14.3% | | 6 months from onset (4 weeks post-infection), followed by linear decline to 12 months |
| Insomnia | 19.4% | | Linear decline from onset (4-weeks post-infection) to 6 months |
| Anxiety | 11.8% | | Linear decline from onset (4-weeks post-infection) to 6 months |
| Depression | 23.2% | | Linear decline from onset (4-weeks post-infection) to 6 months |
| *Children* | | | |
| | *High virulence* | *Low virulence* | |
| Dysosmia | 8.0% | 2.0% | 3 months from onset (1-week post-infection) |
| Headache | 5.0% | 1.3% | 3 months from onset (1-week post-infection) |
| Eye soreness | 2.0% | 0.5% | 3 months from onset (1-week post-infection) |
| Sore throat | 2.0% | 0.5% | 3 months from onset (1-week post-infection) |
| Cognitive difficulties | 3.0% | 0.8% | 3 months from onset (1-week post-infection) |

*Prevalence estimates presented are for unvaccinated sub-groups – each estimate is multiplied by OR = 0.55 to achieve prevalence for vaccinated groups.*

†*Prevalence measured as a risk difference between cases and controls.*

**Community and hospitalised sub-group prevalence estimates achieved through weighting of estimates for mild and severe sub-groups from* [49]

#*No difference in prevalence applied for low vs. high virulence variant infections, for the hospitalised patient sub-group.*

### Appendix Table 15: Long COVID symptom disability weight estimates

| Symptom | Disability Rate† (95% CI) | Health state and justification |
|---|---|---|
| *Adults* | | |
| Dysosmia | 0.01 (0.004-0.020) | Health state: hearing loss, mild & presbyopia.<br>Assumed to be equivalent to mild impairment of other senses. |
| Dysgeusia | 0.01 (0.004-0.020) | Health state: hearing loss, mild & presbyopia<br>Assumed to be equivalent to mild impairment of other senses. |
| Fatigue | 0.051 (0.036-0.062)* | Health state: infectious disease, post-acute consequences (adjusted down for depression and pain).<br>No reasonable estimate exists for fatigue in the 2019 GBD study, however reasonable estimates exist for muscle/joint pain, and depression. Therefore, the DW applied in the present study for depression and muscle/joint pain were subtracted from 'infectious disease, post-acute consequences' DW (DW = 0.219 [95% CI 0.148-0.308]), which is characterised by weakness/tiredness, depression and body pain. |
| Dyspnoea | 0.019 (0.011-0.033) | Health state: chronic obstructive pulmonary disease (COPD) and other chronic respiratory problems, mild. |
| Chest pain | 0.011 (0.005-0.021) | Health state: abdominopelvic problem, mild.<br>Assumed to be equivalent to gastroesophageal reflux disease (GERD). |

| | | |
|---|---|---|
| Muscle weakness | 0.004 (0.001-0.008) | Health state: anaemia, mild.<br>Mild anaemia is characterised by feeling slightly weak/tired. |
| Dizziness | 0.032 (0.021-0.046)* | Health state: vertigo (adjusted to estimate 'mild' vertigo health state).<br>Assumed that dizziness is mild form of vertigo – as there is no 'mild' vertigo DW, 'vertigo' DW (DW=0.113 (95% CI: 0.074-0.158)) was multiplied by 0.29 (average ratio of mild/moderate DWs across all outcomes with mild, moderate, and severe ratings in the GBD study 2019). |
| Muscle/joint pain | 0.023 (0.013-0.037) | Health state: musculoskeletal problems, lower limbs, mild. |
| Headache | 0.037 (0.022-0.057) | Health state: headache, tension-type. |
| Numb/tingling limbs | 0.023 (0.013-0.037) | Health state: musculoskeletal problems, lower limbs, mild. |
| Concentration difficulty | 0.069 (0.046-0.099) | Health state: dementia, mild. |
| Memory impairment | 0.069 (0.046-0.099) | Health state: dementia, mild. |
| Insomnia | 0.03 (0.018-0.046) | Health state: anxiety disorder, mild.<br>Anxiety disorder is characterised by difficulty sleeping and concentrating. |
| Anxiety | 0.03 (0.018-0.046) | Health state: anxiety disorder, mild. |
| Depression | 0.145 (0.099-0.209) | Health state: major depressive disorder, mild. |
| *Children* | | |
| Dysosmia | 0.01 (0.004-0.020) | Health state: hearing loss, mild & presbyopia.<br>Assumed to be equivalent to mild impairment of other senses. |
| Headache | 0.037 (0.022-0.057) | Health state: headache, tension-type. |
| Eye soreness | 0.011 (0.005-0.02) | Health state: presbyopia.<br>No health state for eye pain exists the 2019 GBD study, therefore estimated from presbyopia (characterised by mild near vision loss). |
| Sore throat | 0.006 (0.002-0.012) | Health state: infectious disease, acute episode, mild.<br>Assumed to be equivalent to mild upper respiratory infection. |
| Cognitive difficulty | 0.045 (0.028-0.066) | Health state: attention deficit hyperactivity disorder (ADHD).<br>ADHD is characterised by difficulty with concentration and memory. |

†*Disability Rate applied based on DWs for health states from* [40]
**DW not directly taken from GBD study. Weight and 95% CI estimated by adjusting existing weights.*

Overall long COVID morbidity estimates were calculated for each sub-group, by adding the morbidity contributions for each individual symptom and joint symptom combinations. These are presented in Appendix Table 16. They are applied to the ABM output for the relevant group of symptomatic acute COVID-19 survivors – asymptomatic infections are excluded. Uncertainty is assumed to be high; therefore, a SD of +/- 30% is applied to each estimate.

#### Appendix Table 16: Long COVID morbidity estimates

| **Population group** | | **Unvaccinated (95% UI)** | **Vaccinated (95% UI)** |
|---|---|---|---|
| *High virulence variant, symptomatic* | | | |
| Adults | Non-hospitalised | 0.0075 (0.0031-0.0119) | 0.0038 (0.0016-0.0060) |
| | Hospitalised | 0.0442 (0.0182-0.0701) | 0.0186 (0.0077-0.0296) |
| Children* | | 0.0010 (0.0004-0.0016) | 0.0006 (0.0002-0.0009) |
| *Low virulence variant, symptomatic* | | | |
| Adults | Non-hospitalised | 0.0017 (0.0007-0.0026) | 0.0009 (0.0004-0.0014) |
| | Hospitalised | 0.0442 (0.0182-0.0701) | 0.0186 (0.0077-0.0296) |
| Children* | | 0.0003 (0.0001-0.0004) | 0.0001 (0.00006-0.0002) |

*95% UI produced using +/-30% standard deviation.*
*OR of 0.25 applied to prevalence estimates from high-virulence variant cases to approximate prevalence of long COVID symptoms among low virulence variant cases. OR of 0.55 applied to prevalence estimates from unvaccinated (<2 doses) cases to approximate prevalence of long COVID symptoms among vaccinated cases.*
**Parameterised as ≤18 years in the model.*

#### *Longer term consequences of SARS-CoV-2 infection*

In addition to the long COVID symptoms described above, SARS-CoV-2 infection has been associated with an increased risk of cardiovascular outcomes including myocarditis, stroke, arrhythmias, and acute coronary disease, as well as other sequelae including Type 2 Diabetes Mellitus and acute kidney injury. This risk has been identified particularly in those hospitalised during their acute infection.[59] It is currently unknown whether the risk of chronic outcomes will reduce over time as is assumed for the above discussed long COVID symptoms, or whether such damage confers a lifetime increase in risk. Therefore, this risk has not been included in this model.

#### *Mortality*

For each COVID-19 death during the 18-month intervention duration, we estimate their future HALY loss (3% discount rate) as follows:

- All-cause mortality rates by sex and age for Australia for 2019 were sourced from the Global Health Data Exchange (GHDx). An ongoing 2% per annum fall in mortality rates (uniform by sex and age) was assumed. On average, people dying of COVID-19 have more comorbidities and are frailer than people not dying of COVID-19. Therefore, we assumed the counterfactual future all-cause mortality rates for people dying of COVID-19 (had they not died of COVID-19) would have been twice that of the general population. Given this is uncertain, we assumed a 10% SD about the 2.0 rate ratio (i.e., 95% UI 1.64 to 2.43). A sensitivity analysis for a 1.5-fold rate ratio was also conducted.
- All-cause morbidity rates by sex and age were calculated as the GHDx 2019 YLDs for each sex by age group, divided by the population size in 2019, to generate a proportion of quality of life lost (also called prevalent YLDs, or pYLDs). These were assumed constant into the future by sex and age as all-cause morbidity rates by sex and age have been stable over time in the GBD.[40] Just as people who die of COVID-19 have higher counterfactual mortality rates into the future if they had not died (above), the same applies for morbidity. We used our experience with the New Zealand Burden of Disease study[60] that separately estimates Maori and non-Maori morbidity, where for an all-cause mortality rate ratio of two the all-cause morbidity ratio is approximately 1.5. Accordingly, we assumed the pYLD rate for those dying of COVID-19 into the future (had they counterfactually not died) would have been 1.5 times higher (95% UI 1.23 to 1.82). A sensitivity analysis for a 1.25-fold rate ratio was also conducted.

The ABM has agents stratified by ten-year age group. Accordingly, we estimated the future HALY loss for decedents aged 5, 15, …, 95 and 105 years old. Estimates are shown in Appendix Table 17, with standard deviations across Monte Carlo simulated values using the above uncertainties. The model does not distinguish sexes, so we generated an average HALY loss by age, each with uncertainty expressed as SD as a percentage of the HALY loss estimate. Total HALY loss for each COVID-19 policy scenario was the sum of acute COVID-19 morbidity, long COVID-19 morbidity, and future HALY losses from COVID-19 deaths.

Appendix Table 18 shows HALYs by age (with same % SD for uncertainty as in Appendix Table 17) for HALYs alternatively discounted at 1.5% (following UK Treasury recommendations[61] of health gains discounted at 1.5% and costs at 3.5%, used as part of sensitivity analyses).

Appendix Table 17 also shows the HALY losses for sensitivity analyses using 1.5-fold higher mortality and 1.25 higher morbidity, counterfactually, among COVID-19 deaths.

### Appendix Table 17: HALY losses (3% discount rate) for each COVID death, by sex and age

| | Females | | | Males | | | Combined (average males & females) | |
|---|---|---|---|---|---|---|---|---|
| *Main analysis (2-fold increase in counterfactual ACMR and 1.5-fold increase in counterfactual morbidity, compared with general population, for COVID-19 deaths* | | | | | | | | |
| *Age group* | *Average HALY loss* | *SD HALY loss* | *SD as % of Average* | *Average HALY loss* | *SD HALY loss* | *SD as % of Average* | *Average HALY loss* | *Average SD as % of HALY loss* |
| 0-9 yrs | 23.94 | 0.55 | 2.40% | 24.28 | 0.48 | 2.00% | 24.11 | 2.20% |
| 10-19 yrs | 21.89 | 0.65 | 3.00% | 22.23 | 0.56 | 2.50% | 22.06 | 2.75% |
| 20-29 yrs | 19.86 | 0.7 | 3.60% | 20.06 | 0.6 | 3.00% | 19.96 | 3.30% |
| 30-39 yrs | 17.7 | 0.7 | 4.00% | 17.62 | 0.61 | 3.50% | 17.66 | 3.75% |
| 40-49 yrs | 15.04 | 0.68 | 4.60% | 14.6 | 0.61 | 4.10% | 14.82 | 4.35% |
| 50-59 yrs | 11.82 | 0.64 | 5.50% | 11 | 0.57 | 5.20% | 11.41 | 5.35% |
| 60-69 yrs | 8.17 | 0.56 | 6.80% | 7.2 | 0.5 | 6.90% | 7.685 | 6.85% |
| 70-79 yrs | 4.54 | 0.41 | 9.10% | 3.82 | 0.36 | 9.40% | 4.18 | 9.25% |
| 80-89 yrs | 1.78 | 0.23 | 13.10% | 1.52 | 0.2 | 13.00% | 1.65 | 13.05% |
| 90-99 yrs | 0.48 | 0.1 | 20.90% | 0.5 | 0.09 | 18.20% | 0.49 | 19.55% |
| *Sensitivity analysis (1.5-fold increase in counterfactual ACMR and 1.25-fold increase in counterfactual morbidity), compared with general population, for COVID-19 deaths (only expected values re-estimated; SD HALY loss as above)* | | | | | | | | |
| *Age group* | *Average HALY loss* | | | *Average HALY loss* | | | *Average HALY loss* | |
| 0-9 yrs | 25.00 | | | 24.24 | | | 24.62 | |
| 10-19 yrs | 23.16 | | | 22.19 | | | 22.68 | |
| 20-29 yrs | 21.29 | | | 20.01 | | | 20.65 | |
| 30-39 yrs | 19.23 | | | 17.58 | | | 18.40 | |
| 40-49 yrs | 16.64 | | | 14.55 | | | 15.60 | |
| 50-59 yrs | 13.43 | | | 10.96 | | | 12.20 | |
| 60-69 yrs | 9.68 | | | 7.17 | | | 8.42 | |
| 70-79 yrs | 5.77 | | | 3.80 | | | 4.78 | |
| 80-89 yrs | 2.54 | | | 1.51 | | | 2.02 | |
| 90-99 yrs | 0.83 | | | 0.49 | | | 0.66 | |

*100% correlation on uncertainty between age groups*

### Appendix Table 18: HALY losses (1.5% discount rate) for each COVID death, by sex and age

| | Females | Males | Combined (average males & females) |
|---|---|---|---|
| *Age group* | *Average HALY loss* | *Average HALY loss* | *Average HALY loss* |
| 0-9 yrs | 36.40 | 36.66 | 36.53 |
| 10-19 yrs | 32.38 | 32.49 | 32.43 |
| 20-29 yrs | 28.28 | 28.12 | 28.20 |

| | | | |
|---|---|---|---|
| 30-39 yrs | 24.06 | 23.56 | 23.81 |
| 40-49 yrs | 19.43 | 18.55 | 18.99 |
| 50-59 yrs | 14.49 | 13.27 | 13.88 |
| 60-69 yrs | 9.48 | 8.27 | 8.88 |
| 70-79 yrs | 5.01 | 4.19 | 4.60 |
| 80-89 yrs | 1.87 | 1.60 | 1.74 |
| 90-99 yrs | 0.48 | 0.51 | 0.50 |

### Economic analyses

#### *Testing costs*

In Victoria, approximately one third of cases are detected with PCR and two thirds detected with rapid antigen testing (RAT).[62] We included costs for PCR tests based on a reported PCR test positivity rate of approximately 20%.[63] Given we assume 50% case ascertainment (i.e., the number of infections output in our model is double the number of cases that would likely be notified in Victoria) we applied a cost of 0.83 PCR tests (0.5*0.33*5) to each infection. The current RAT positivity rate in Victoria is unknown, given only positive RAT results are reported. As such we assumed a 10% RAT positivity rate (given extensive use of RATs following, for example, exposure to a confirmed case in Victoria) and applied the cost of 3.33 RAT kits (0.5*0.66*10) to each infection. Uncertainty of +/- 20% was applied to the number of PCR/RAT tests. Unit costs for PCR and RAT testing were estimated at $42.50 (MBS item 69479)[64] with +/- 10% uncertainty applied and $7.60 (median wholesale price as reported by the Australian Competition and Consumer Commission[65], with +/- 10% uncertainty applied) respectively.

#### *Acute COVID-19 morbidity expenditure*

We estimated acute COVID-19-related health expenditure using an ingredients approach. For each clinical patient subgroup, we estimated the expected patient pathway through the health system and calculated total health expenditure by first estimating resource use required for a typical patient (e.g., hospital or outpatient visits, or drugs) and then multiplying by unit costs for each of the specified resources based on model outputs. For ICU-admitted patients, we added the cost of their ICU stay onto the cost of a hospital stay.

Appendix Table 19 shows resource use assumptions. The SARS-CoV-2 pandemic is expected to add additional costs to hospital operations, adjusting for complexity of patients and added infection control requirements including the need for isolation of patients, fitting of personal protective equipment, and enhanced cleaning regimens. As a result, inpatient and ICU hospital costs have been scaled up by 20% to account for these additional costs.

**Appendix Table 19: Resource use assumptions by SARS-CoV-2 category**

| Patient Subgroup | Resource | Explanation |
|---|---|---|
| Treated in ICU (note for these patients, resources from a non-ICU hospitalisation stay are also added) | GP visit | Assume all patients will have 2 GP consultations (in addition to those applied for non-ICU hospitalisation), for example as part of post-discharge follow-up |
| | ER visit | Captured in non-ICU hospitalisation, no additional cost applied |
| | ICU | Daily cost of ICU admission multiplied by length of ICU stay (see Appendix Table 7) |
| | Inpatient (non-ICU) | Added as per costs for non-ICU hospitalisation below |
| | Pandemic loading | All hospital costs have 20% loading for pandemic |
| Hospitalised | GP visit | Assume all patients will have 2 GP consultations prior to hospital admission |
| | ER visit | Base fee charged for presenting to an ER |
| | Inpatient | Cost of inpatient day multiplied by length of stay (see Appendix Table 7) |
| | Pandemic loading | All hospital costs have 20% pandemic loading |
| | Readmission | Approximately 10% of hospitalised patients are readmitted.[66] As such, hospitalisation costs are multiplied by 1.1. |
| Symptomatic | GP Visit | Assume half of all patients attend a GP appointment during their illness |
| | Paracetamol | Assume all patients will purchase 1 packet of paracetamol for symptomatic treatment |
| Asymptomatic | No resources | N/A |

*ER: emergency room, ICU: intensive care unit, LOS: length of stay, GP: general practitioner*

Unit costs for each resource are shown in Appendix Table 20.

**Appendix Table 20: Unit costs for healthcare resource use**

| Resource | Cost (2022 AUD) | Source | Detail |
|---|---|---|---|
| Paracetamol | $6.80 | PBS item 10582Y (100 units, 500mg)[67] | For symptomatic treatment of mild illness |

| | | | |
|---|---|---|---|
| GP visit | $39.10 | MBS item 23 (level B general consultation)[68] | |
| ER visit for dyspnoea | $907.68 | Data from NHCDC Report Round 2014 (financial year 2019-20)[69] | Adjusted to 2022 cost |
| Inpatient day | $1644.06 | Data from H1N1 outbreak using AR-DRG code[70] | Total cost of inpatient day post ICU. Also assumed for all non-ICU patients. Adjusted to 2022 cost. |
| ICU day | $6710.86 | Micro-costed from H1N1 outbreak[70] | Total cost per day admitted to ICU. Allied health and overheads included. Adjusted to 2022 cost. |

*PBS: pharmaceutical benefits scheme, MBS: Medicare benefits schedule, AR-DRG: Australian-refined diagnostic related groups, ICU: intensive care unit. All costs have +/- 10% uncertainty on log normal distribution applied.*
*All costs in Australian dollars*

Given the above, the following costs were applied for acute COVID-19-related health expenditure (note these costs are additive, unlike the morbidity calculations above):

- For an infected, asymptomatic agent: no additional costs
- For an infected, symptomatic agent: add 0.5*[GP visit cost] and 1*[paracetamol cost]
- If an agent is hospitalised (*in addition* to symptomatic agent costs):
    - Add 1.5*[GP visit cost] and 1.2*[ER visit cost]
    - Add length of hospital stay (days)*[cost of inpatient hospital bed per day]*1.1*1.2
- If an agent is admitted to ICU (*in addition* to hospitalised agent costs):
    - Add 2*[GP visit cost]
    - Add length of ICU stay (days)*[cost of ICU bed per day]*1.2

#### *Long COVID morbidity expenditure*

Few observational studies of long COVID occurrence, in Australia or internationally, have included thorough data collection regarding healthcare utilisation following acute COVID-19 infection. Additionally, clinical data on healthcare use relating to long COVID is not currently collected in Australia – the ICD-10 code assigned for long COVID is yet to have associated published hospital data. Therefore, in order to estimate the healthcare costs associated with long COVID, we applied expert estimates on the likely healthcare use for those experiencing long COVID symptoms, based on available international data and formal guidelines for healthcare professionals in Australia provided by the Royal Australian College for General Practitioners (RACGP) and the National COVID-19 Clinical Evidence Taskforce.[71,72] An additional informal expert knowledge elicitation was also conducted with General Practitioners (GPs) to confer information obtained from the former sources. Appendix Table 21 shows the resources applied to each sub-group; unit costs are indicated in Appendix Table 22. For estimates specific to particular long COVID symptoms, costs are assigned to proportions of symptomatic acute COVID-19 survivors with the relevant long-COVID-19 symptom (see 'long COVID morbidity' section above for methodology).

The following treatment items are considered:

- GP attendance: current guidelines indicate that long COVID is predominantly managed in the primary care setting, though few evidence-based management options currently exist.[71] In a survey of COVID-19 survivors 6-months post-acute illness (pre-Omicron, unvaccinated cohort) in Zurich, Switzerland, Menges et al. found that 63% of previously hospitalised adults, and 29% adult community cases, had at least 1 GP consult related to COVID-19 following the acute illness period.[73] The estimate for hospitalised adults is multiplied by 1.5 to include GP use in the second 6 months (reflecting the estimated duration of long COVID for this sub-group).
- Specialist attendance: a survey of symptomatic COVID-19 survivors (pre-Omicron, unvaccinated cohort) attending a post-COVID assessment clinic in London found that 18% were referred on to specialist management, most commonly to a cardiologist, neurologist or ENT specialist.[74] Therefore, 18% of those who attend a GP at least once are assumed to go on to see a specialist (single consult).
- ER attendance: ER attendance is calculated specifically for those experiencing dyspnoea or chest pain – these groups are assumed to present to ER once throughout the disease course.
- Medication: medication use is estimated based on RACGP guidelines and informal expert elicitation with GPs. Short-acting beta-agonists (salbutamol), or corticosteroids (budesonide), are applied for patients with ongoing dyspnoea.[72] Paracetamol is applied for those with headache and muscle/joint pain.[72]

- Diagnostics: routine testing is advised by RACGP to rule out other causes of symptoms, e.g., anaemia. Therefore, for those with fatigue, routine blood testing including iron studies, a full blood examination, electrolytes and thyroid function tests are costed. It is assumed that any diagnostic tests required for those with ongoing dyspnoea/chest pain (e.g., chest x-ray or ECG as per RACGP guidelines) occur at ER presentation, so are not separately included to avoid double costing. Other symptoms are judged as being too infrequent to consider any other specific testing.

Magnusson et al. conducted a large study in Norway on healthcare use in children in the period 5-12 weeks post-COVID infection.[75] No significant difference was found in healthcare utilisation between cases and COVID-negative controls during this period. This is in keeping with the relatively low morbidity impact of long COVID estimated for children compared to adults.  Therefore, costs for children are not considered.

The proportions indicated in Appendix Table 21 are calculated and applied for each sub-group of initially symptomatic acute COVID-19 survivors. For symptom-specific costs, proportions reflect long COVID symptom frequencies calculated for each patient sub-group (see long COVID morbidity section above for methodology). GP and specialist attendance proportions applied from the aforementioned literature are multiplied by an OR of 0.55 and separately by an OR of 0.25 to reflect use in vaccinated sub-groups and for low virulence variant infections (e.g., Omicron), respectively (in line with long COVID morbidity estimates).[52-54]

**Appendix Table 21*: Resource use assumptions for COVID-19 survivors by SARS-CoV-2 category***

| Patient subgroup | Treatment item | Source |
|---|---|---|
| Community (acute infection) adult cases | GP visit | 20% acute COVID-19 community cases have 1-2 visits, 7% 3-5 visits, 1% 6 visits.[73] For vaccinated groups, proportions are multiplied by an OR of 0.55. For low virulence variant groups, proportions are multiplied by an OR of 0.25. |
| | Specialist visit | 18% who see GP (above) are referred to specialist.[74] Assume single visit. For vaccinated groups, proportions are multiplied by an OR of 0.55. For low virulence variant groups, proportions are multiplied by an OR of 0.25. |
| | ER visit | Assume those with dyspnoea and those with chest pain present to ER once. Standard ER presentation cost for asthma (to represent dyspnoea) and chest pain, are applied. |
| | Medication (budesonide, salbutamol) | Assume patients with dyspnoea are prescribed either salbutamol or budesonide (50/50 split) (single prescription with 5 repeats, equating to approximately 6 months use). |
| | Medication (paracetamol) | Assume patients with muscle/joint pain and headache purchase 1 packet of paracetamol. |
| | Diagnostics | Iron studies for those with fatigue to rule out anaemia, per RACGP guidelines.[72] Routine testing of FBC, electrolytes and TFTs also included for this group. |
| Hospitalised (acute infection) adult cases | GP visit | During first 6 months, 44% 1-2 visits, 12% 3-5 visits, 5% 6 visits.[73] Proportions halved in the second 6 months, to reflect longer duration of symptoms compared to non-hospitalised patients. For vaccinated groups, proportions are multiplied by an OR of 0.55. |
| | Specialist visit | 18% who see GP are referred to specialist.[74] Assume single visit. |
| | ER visit | Assume those with dyspnoea and those with chest pain present to ER once. Standard ER presentation cost for asthma (to represent dyspnoea) and chest pain, are applied. |
| | Diagnostics | Iron studies for those with fatigue to rule out anaemia, per RACGP guidelines. Also include routine FBC, electrolytes and TFT testing. |
| | Medication (budesonide, salbutamol) | Assume patients with dyspnoea are prescribed either salbutamol or budesonide (50/50 split) (single prescription with 5 repeats, equating to approximately 6 months use). Proportions halved in the second 6 months, to reflect longer duration of symptoms compared to non-hospitalised patients. |

| | | |
|---|---|---|
| | Medication (paracetamol) | Assume patients with muscle/joint pain and headache purchase 1 packet of paracetamol. |

GP: general practitioner; ER: emergency room; FBC: full blood count; TFT: thyroid function tests.

#### Appendix Table 22: Unit costs for long COVID healthcare resource use by COVID-19 survivors

| Resource and cost (2022 AUD) | Source | Detail |
|---|---|---|
| ER visits<br><br>ER visit for chest pain<br>$998.97 (plus $199.79 for 20% pandemic loading)<br><br>ER visit for dyspnoea<br>$907.68 (plus $181.54 for 20% pandemic loading) | Data from NHCDC Report Round 2014 (financial year 2019-20)[69] | Average cost per ER presentation, using AECC codes for chest pain and asthma (complexity level B for each). Adjusted to 2022 value. |
| Medication<br><br>Budesonide (dyspnoea)<br>$40.13<br><br>Salbutamol (dyspnoea)<br>$26.30<br><br>Paracetamol (headache/muscle/joint pain)<br>$6.80 | PBS item 2065Q (500µg/2mL inhalation solution, 30 x 2mL ampoules)[76]<br><br>PBS item 8288F (100µg/actuation inhalation, 200 actuations)[77]<br><br>PBS item 10582Y (500mg tablets, 100 units)[67] | PBS fee for budesonide with 5 repeats<br><br>PBS fee for salbutamol with 5 repeats<br><br>PBS fee for paracetamol with 1 repeat |
| GP consultation<br>$39.10 | MBS item 23. Level B General consultation.[68] | MBS fee for standard attendance by general practitioners. |
| Specialist consultation<br>$76.80 | MBS item 104. Initial referred consultation.[78] | MBS fee for standard initial attendance by specialist practitioner. |
| Pathology testing<br><br>Iron studies<br>$27.70<br><br>TFT<br>$29.60<br><br>FBE (including FBC)<br>$14.45<br><br>Electrolytes<br>$13.35 | MBS item 66596[79]<br><br>MBS item 66719[80]<br><br>MBS item 65070[81]<br><br>MBS item 66509[82] | MBS fees for pathology testing items. |

*ER: emergency room; NHCDC: national hospital cost data collection; AECC: Australian emergency care classification; PBS: pharmaceutical benefits scheme; GP: general practitioner; MBS: Medicare benefits schedule; TFT: thyroid function tests; FBE: full blood examination; FBC: full blood count*
*All costs in Australian dollars*

The total cost for each patient sub-category is presented in Appendix Table 23. Each overall cost is applied to the relevant sub-group of all previously symptomatic acute COVID-19 survivors.

#### Appendix Table 23: Cost per patient category

| Population group | | Unvaccinated (95% UI) | Vaccinated (95% UI) |
|---|---|---|---|
| *High virulence variant symptomatic* | | | |
| Adults | Non-hospitalised | $95.29 (39.26-151.32) | $52.98 (21.83-84.14) |
| | Hospitalised | $269.76 (111.14-428.37) | $148.37 (61.13-235.60) |
| *Low virulence variant symptomatic* | | | |
| Adults | Non-hospitalised | $24.08 (9.92-38.24) | $13.25 (5.46-21.03) |
| | Hospitalised | $269.76 (111.14-428.37) | $148.37 (61.13-235.60) |

*UI applied using +/- 30%, in line with uncertainty applied for long COVID morbidity estimates. Costs apply to adults (≥18 years). All costs in Australian dollars.*

### *Intervention costs*

#### Appendix Table 24: Vaccine and respirator costs

| | Item | Price (2022 AUD) | SD | Source |
|---|---|---|---|---|
| Vaccination costs | Current mRNA vaccine (per dose) | $35 | 10% | Reported in [83] |
| | Omicron-specific vaccine (per dose)* | $35 | 10% | Estimated as equivalent in cost to current mRNA vaccines |
| | Multivalent vaccine (per dose)* | $52.50 | 10% | Estimated 1.5 times the cost of current generation mRNA vaccine |
| | Vaccine transport (per dose) | $1 | 10% | Estimate |
| | Personnel, consumables, waste management (per dose) | $30 | 10% | Relevant MBS items for GP administration of vaccines and rebate for pharmacy administration[84-87] |
| | Overheads (per dose) | $1 | 10% | Estimate |
| | Promotion and advertising (per month) | $615,000 | 20% | Australian Government Department of Finance report on COVID-19 vaccination campaign advertising spending 2020-2021 financial year, scaled to Victorian population and one month, updated to 2022 cost and rounded to nearest $5,000[88] |
| Respirator policy costs | Respirator costs (per person, per round, i.e., 10 masks) | $14.70 | 10% | Commercial price for N95 masks in Australia (quoted from supplier, for bulk order) |
| | Respirator distribution cost (per person, per round) | $2.73 | 10% | Assumed 20% of the cost of vaccine administration cost and 2.2 persons aged ≥10 years in each household collecting respirators |
| | Overheads (per person, per round of respirator distribution) | $0.09 | 10% | Assumed 20% of that for vaccination program and 2.2 persons aged ≥10 years in each household collecting respirators |
| | Promotion and advertising (per day) | $15,164 | 20% | Estimated from Australian Government Department of Finance report on general COVID-19 health campaign advertising spending 2020-2021 financial year, scaled to Victorian population and one day, updated to 2022 cost and rounded to nearest $5,000[88] |
| | Mask storage over 18 months (full stockpile) | $295,000 | 20% | 6,400 respirators per pallet, commercial price for storage $4.50 per pallet per week (quoted from supplier), 10 respirators per person per round, |

| | | | | |
|---|---|---|---|---|
| | | | | approximately 586,250 ≥10-year-olds in Victoria. Rounded to nearest $5,000 |
| Increased mask wearing policy cost | Promotion and advertising (per day) | $15,164 | 20% | Estimated from Australian Government Department of Finance report on general COVID-19 health campaign advertising spending 2020-2021 financial year, scaled to Victorian population and one day, updated to 2022 cost and rounded to nearest $5,000[88] |

*[*]Uncertainty on vaccine dose costs 100% correlated.*
*All costs in Australian dollars*

Regarding fixed costs for vaccines, three months of promotion and advertising is conducted per vaccination rollout event (i.e., with each subsequent dose that becomes available). The respirator policy incurs an initial cost for mask storage (on 1 April 2022) and additional costs depending time spent in stage 3 or greater. The number of restock rounds is $\lceil(\text{time in stage} \geq 3)/28\rceil$, and the restock costs are applied to everyone over 10 years old. The days spent on promotion and advertising is just the time spent in stage 3 or greater. This advertising cost is the only cost associated with the increased mask usage policy, and when both policies are active the cost of promotion and advertising is doubled. Differences in costs between high and low vaccine coverage scenarios are driven by the differences in the number of vaccine doses administered (i.e., sunk costs in a low coverage scenario whereby vaccines are purchased and then not administered are not included).

#### *GDP costs of stages of PHSMs*

We updated previously published estimates of GDP loss due to government-imposed pandemic control interventions[89] to estimate the current economic impact of time spent in various stages of PHSMs. Due to societal changes since the beginning of the pandemic (such as adaptations to facilitate remote working) we assumed no GDP impacts in stages 1 and 2. GDP loss in stages 3 – 5 in 2022 and 2023 are difficult to estimate but given societal change we assumed wide uncertainty of 10% to 50% (uniform distribution) of the GDP losses per week estimated in 2020. GDP losses per stage were thus estimated by the following (Appendix Table 25):

**Appendix Table 25: GDP loss by stage of PHSMs (per week)**

| Stage | GDP loss per week in stage (AUD billions, Victoria only) |
|---|---|
| 1 | $0 |
| 2 | $0 |
| 3 | 10% to 50% (uniform distribution) of $0.725 |
| 4 | 10% to 50% (uniform distribution) of $1.275 |
| 5 | 10% to 50% (uniform distribution) of $2.61 |

#### *Unrelated disease costs*

Consistent with recommended practice in cost effectiveness analyses,[90-93] we included ‘unrelated disease costs’ in the economic evaluation. This means that in addition to including the health expenditure on SARS-CoV-2 cases as above, future changes in health system expenditure are also included. This means that if someone dies due to SARS-CoV-2 infection, their reduced health expenditure in the future is included (leading to a potentially net negative expenditure depending on the balance of costs, age and discount rate).

We estimated these future health expenditure savings for each COVID-19 death in the 18-month intervention duration, using an approach similar to that above for future HALY losses (Appendix Table 17). Specifically, we used lifetables with sex and age-specific all-cause mortality rates (double that of the general population due to people dying of COVID-19 having more co-morbidities and frailty) to estimate remaining life years. For each expected remaining life-year, 1.5 times (due to greater comorbidities) the sex and age-specific expected annual health expenditure according to AIHW estimates[94] was allocated.

We assumed variable health expenditure was 90%, allowing 10% for fixed costs in running services. AIHW estimates capture 62.7% of total health expenditure, therefore we multiplied all age by sex empirical estimates by a factor of 90/62.7 to generate the estimated predicted Australian variable health expenditure per capita, by age and sex. Next, we adjusted these expenditures for inflation to 2022 Australian dollars using Australian consumer price index adjustment factors. Finally, we also generated US dollar values using OECD purchasing power parity (https://data.oecd.org/price/inflation-cpi.htm; 2021 PPP used as 2022 not available at the time of writing).

Appendix Table 26 shows future averted health expenditure per COVID-19 death (discount rate 3%). Males and females are not distinguished separately in the ABM. Therefore, also shown in Appendix Table 26 are averages by sex, which are used in the modelling. Noting the SD about the sex-specific estimates, we set the SD as 15% of the age-specific estimates (normal distribution).

Appendix Table 27 shows future averted health expenditure at the discount rate of 3.5% as recommended by the UK Treasury[61] (with the same % SD for uncertainty as in Appendix Table 26).

#### Appendix Table 26: Future averted health expenditure for each COVID-19 death (3% discount rate), by sex and age

| | **Females** | | | **Males** | | | **Combined (average of males and females)** | |
|---|---|---|---|---|---|---|---|---|
| *Age group* | *Expected value, AUD, CPI adjusted to 2022* | *Expected value, USD, OECD PPP adjusted* | *SD in Monte Carlo uncertainty analyses as % of expected* | *Expected value, AUD, CPI adjusted to 2022* | *Expected value, USD, OECD PPP adjusted* | *SD in Monte Carlo uncertainty analyses as % of expected* | *Expected value, AUD, CPI adjusted to 2022* | *Expected value, USD, OECD PPP adjusted* |
| *Main analysis (2-fold increase in counterfactual ACMR and 1.5 fold increase in counterfactual morbidity, compared with general population, for COVID-19 deaths* | | | | | | | | |
| 0-9 yrs | $79,825 | $54,526 | 15.90% | $97,190 | $66,387 | 12.40% | $88,508 | $60,457 |
| 10-19 yrs | $90,913 | $62,099 | 15.70% | $77,596 | $53,003 | 12.10% | $84,255 | $57,551 |
| 20-29 yrs | $158,112 | $108,000 | 15.70% | $79,192 | $54,093 | 12.30% | $118,652 | $81,047 |
| 30-39 yrs | $215,990 | $147,534 | 15.00% | $99,970 | $68,285 | 11.70% | $157,980 | $107,910 |
| 40-49 yrs | $175,489 | $119,869 | 14.00% | $128,337 | $87,662 | 11.10% | $151,913 | $103,766 |
| 50-59 yrs | $180,501 | $123,293 | 13.60% | $160,764 | $109,811 | 11.00% | $170,633 | $116,552 |
| 60-69 yrs | $193,058 | $131,870 | 13.90% | $191,507 | $130,811 | 11.20% | $192,283 | $131,341 |
| 70-79 yrs | $188,429 | $128,708 | 14.40% | $188,763 | $128,936 | 11.50% | $188,596 | $128,822 |
| 80-89 yrs | $123,066 | $84,061 | 14.70% | $130,017 | $88,810 | 11.70% | $126,542 | $86,436 |
| 90-99 yrs | $56,881 | $38,853 | 14.30% | $67,573 | $46,157 | 11.60% | $62,227 | $42,505 |
| *Sensitivity analysis (1.5-fold increase in counterfactual ACMR and 1.25-fold increase in counterfactual morbidity), compared with general population, for COVID-19 deaths (only expected values re-estimated; SD HALY loss as above)* | | | | | | | | |
| 0-9 yrs | $75,952 | $51,880 | 15.90% | $92,474 | $63,166 | 12.40% | $84,213 | $57,523 |
| 10-19 yrs | $86,502 | $59,086 | 15.70% | $73,831 | $50,431 | 12.10% | $80,166 | $54,759 |
| 20-29 yrs | $150,440 | $102,759 | 15.70% | $75,350 | $51,468 | 12.30% | $112,895 | $77,114 |
| 30-39 yrs | $205,509 | $140,375 | 15.00% | $95,119 | $64,971 | 11.70% | $150,314 | $102,673 |
| 40-49 yrs | $166,973 | $114,052 | 14.00% | $122,110 | $83,408 | 11.10% | $144,541 | $98,730 |
| 50-59 yrs | $171,743 | $117,310 | 13.60% | $152,963 | $104,482 | 11.00% | $162,353 | $110,896 |
| 60-69 yrs | $183,690 | $125,471 | 13.90% | $182,214 | $124,463 | 11.20% | $182,952 | $124,967 |
| 70-79 yrs | $179,285 | $122,462 | 14.40% | $179,603 | $122,679 | 11.50% | $179,444 | $122,571 |
| 80-89 yrs | $117,094 | $79,982 | 14.70% | $123,708 | $84,501 | 11.70% | $120,401 | $82,241 |
| 90-99 yrs | $54,120 | $36,967 | 14.30% | $64,294 | $43,917 | 11.60% | $59,207 | $40,442 |

*AUD: Australian dollars, USD: US dollars, CPI: consumer price index, OECD: Organisation for Economic Co-operation and Development, PPP: purchasing power parity, SD: standard deviation*
*100% correlation on uncertainty between age groups*

#### Appendix Table 27: Future averted health expenditure for each COVID-19 death (3.5% discount rate), by sex and age

| | Females | Males | Combined (average males & females) |
|---|---|---|---|
| *Age group* | *Expected value, AUD, CPI adjusted to 2022* | *Expected value, AUD, CPI adjusted to 2022* | *Expected value, AUD, CPI adjusted to 2022* |
| 0-9 yrs | $69,103 | $84,459 | $76,781 |
| 10-19 yrs | $79,331 | $67,810 | $73,570 |
| 20-29 yrs | $139,561 | $70,328 | $104,944 |
| 30-39 yrs | $192,419 | $89,641 | $141,030 |
| 40-49 yrs | $158,200 | $116,688 | $137,444 |
| 50-59 yrs | $165,886 | $149,004 | $157,445 |
| 60-69 yrs | $181,354 | $181,151 | $181,252 |
| 70-79 yrs | $180,690 | $181,910 | $181,300 |
| 80-89 yrs | $120,000 | $127,094 | $123,547 |
| 90-99 yrs | $56,044 | $66,613 | $61,328 |
| 100-109 yrs | $42,507 | $52,934 | $47,720 |
